# Analytic Interoperability Framework for High-Fidelity Representation of Perioperative Electronic Health Record Data in the OMOP Common Data Model Using Clinical Representation and Analytic Concept Reconstruction

**DOI:** 10.64898/2026.10.30.26366954

**Authors:** Jacqueline C. Stocking, Orion Stewart, Garth H. Utter, Jason Y. Adams, Nicholas J. Kenyon, Christian E. Sandrock, Timothy E. Albertson, Joe G.N. Garcia, Ivo L. Abraham

## Abstract

**Objective:** To evaluate whether complex perioperative electronic health record (EHR) data can be represented within the Observational Medical Outcomes Partnership (OMOP) Common Data Model (CDM) while preserving clinically meaningful analytic concepts.

**Materials and Methods:** Data from 31,158 consecutive adult elective surgical encounters (October 2015– September 2025) were extracted from an academic medical center’s Epic Clarity database. Source concepts required for a postoperative respiratory failure dataset were manually mapped to OMOP using the OHDSI Athena vocabulary and classified by Health Level 7 equivalence categories under strict and lenient criteria. SQL-based workflows reconstructed clinical representations and analytic concepts, which were validated against the native Epic implementation.

**Results:** The reference dataset comprised 28 analytic variables derived from 538 source concepts. Under strict criteria, 40.7% of mappings were equal; under lenient criteria permitting clinical equivalence despite terminology or granularity differences, 97.4% were classified as equal. OMOP achieved >99% cohort concordance. Among 31,083 shared encounters, reconstructed analytic variables showed high agreement, with meaningful differences primarily involving vasopressor infusion identification, body mass index selection, and payer assignment; admission source accounted for most discordant cohort assignments.

**Discussion:** Concept equivalence alone incompletely characterized analytic interoperability. Reconstruction of clinical representations and analytic concepts from standardized OMOP concepts preserved analytic fidelity despite incomplete exact concept matching.

**Conclusions:** Complex perioperative EHR data can be represented within OMOP with high implementation fidelity despite imperfect concept equivalence. Evaluating preservation of clinically meaningful analytic concepts, rather than vocabulary equivalence alone, provides a practical framework for interoperable perioperative research.

## MAIN TEXT

### OBJECTIVE

To evaluate whether complex perioperative electronic health record data can be represented within the Observational Medical Outcomes Partnership (OMOP) Common Data Model while preserving clinically meaningful analytic concepts. Using a postoperative respiratory failure dataset as a representative use case, we assessed concept coverage, standardized extract-transform-load implementation, and fidelity of the OMOP-derived analytic dataset relative to the native Epic implementation.

### BACKGROUND AND SIGNIFICANCE

Electronic health records (EHRs) contain detailed perioperative data that support clinical decision-making, predictive modeling, and observational research. Increasingly, structured perioperative EHR data are also being used for electronic quality measurement, including recently developed national electronic clinical quality measures for postoperative respiratory failure.[1] However, differences in local data structures, clinical terminology, and extract-transform-load (ETL) processes make these data difficult to harmonize across institutions. Common data models (CDMs) address this challenge by standardizing the representation of clinical information, enabling interoperable analytics, reusable implementation workflows, and multicenter federated research across heterogeneous observational databases.[2 3] Among these, the Observational Medical Outcomes Partnership (OMOP) Common Data Model (CDM), developed by the Observational Health Data Sciences and Informatics (OHDSI) collaborative, has emerged as one of the most widely adopted frameworks for transforming heterogeneous clinical data into a unified structure using standardized vocabularies,[4 5] enabling consistent analyses across healthcare systems and supporting multicenter federated research.[2 6 7] Comparative evaluations have further demonstrated that OMOP provides broader clinical content coverage and terminology support than several alternative research CDMs, making it well suited for heterogeneous EHR data.[8]

Database interoperability requires systematic harmonization of disparate data structures and terminologies through *crosswalking*, in which local concepts are mapped to standardized vocabularies while preserving clinical meaning.[5 9] This is particularly challenging in perioperative and critical care data, where clinically meaningful variables frequently depend on temporal relationships, derived calculations, and workflow-specific context rather than individual concepts alone.[9–11] Prior transformations of EHR and anesthesia information systems into OMOP have therefore required local vocabulary adaptation, implementation-specific ETL, and, in some cases, additional structures for computed features and perioperative episodes.[10 12] Similar challenges have been reported across critical care, biobank, primary care, pulmonary hypertension, and diabetes implementations.[13–17] Consistent with these findings, our prior ICU evaluation found that only 49.6% of EHR concepts mapped directly to equivalent standard OMOP concepts.[9] These findings demonstrate that successful standardization of complex clinical domains requires preservation of analytic relationships and context beyond one-to-one vocabulary mapping.

Previous OMOP evaluations across ophthalmology, anesthesia, critical care, nursing, and multi-institutional settings have largely emphasized vocabulary mapping, semantic harmonization, structural implementation, and data quality.[2 3 5 8-10 12-14 18-22]

However, concept equivalence does not establish whether transformation preserves the analysis-ready dataset produced by the native EHR implementation. We define *implementation fidelity* as the degree to which a standardized implementation reproduces native cohort construction, clinical representations, derived analytic variables, and resulting analysis-ready data despite differences in underlying representation. To our knowledge, implementation fidelity has not been systematically evaluated for complex perioperative EHR data.

Although implementation fidelity is necessary for interoperable research, it is not sufficient. Ultimately, the objective of transforming perioperative EHR data into a common data model is to enable clinically meaningful analyses, including prediction modeling, quality measurement, and multicenter observational research.[1] Demonstrating that standardized implementations faithfully reproduce analysis-ready datasets therefore represents a critical intermediate step toward transportable analytic frameworks that can be implemented consistently across institutions.

## MATERIALS AND METHODS

### Study Design

We conducted a retrospective single-center informatics evaluation using Epic Clarity data from 31,158 consecutive adult elective surgical encounters performed between October 1, 2015, and September 30, 2025, at an academic medical center. This study evaluated the technical feasibility of representing perioperative electronic health record data within the Observational Medical Outcomes Partnership (OMOP) Common Data Model (CDM) and assessed whether clinically meaningful analytic concepts could be reconstructed with high implementation fidelity relative to the native Epic implementation. This study was approved by IRB #822917 with waiver of consent and waiver of HIPAA authorization.

### Reference Perioperative Dataset

The data elements required for a previously developed electronic health record–based prediction model of postoperative respiratory failure (PRF)[23] served as the reference analytic dataset for OMOP implementation and validation. The native Epic cohort included 265 surgical encounters with PRF and 30,893 without PRF. The reference dataset comprised perioperative timestamps, demographic characteristics, physiologic measurements, medications, fluid administration, operative variables, outcomes, and hospital utilization measures required to construct the analytic dataset.

### Epic-to-OMOP Concept Mapping and Analytic Concept Development

To support reproducible analytic interoperability, we developed a hierarchical framework that distinguished between source concepts, standardized OMOP concepts, clinical representations, and analytic concepts (Figure 1). The framework comprised five sequential levels: (1) native Epic EHR source concepts, (2) standardized OMOP concepts, (3) ETL-based clinical representations, (4) derived analytic concepts, and (5) an analysis-ready analytic dataset. Rather than assuming a one-to-one correspondence between source and standardized concepts, the framework explicitly separated vocabulary standardization, clinical representation, and analytic concept derivation, allowing clinically meaningful variables to be reconstructed from standardized concepts using reproducible ETL logic. A detailed description of this conceptual framework and guiding principles is provided in Supplementary Method 1.

**Figure 1:**
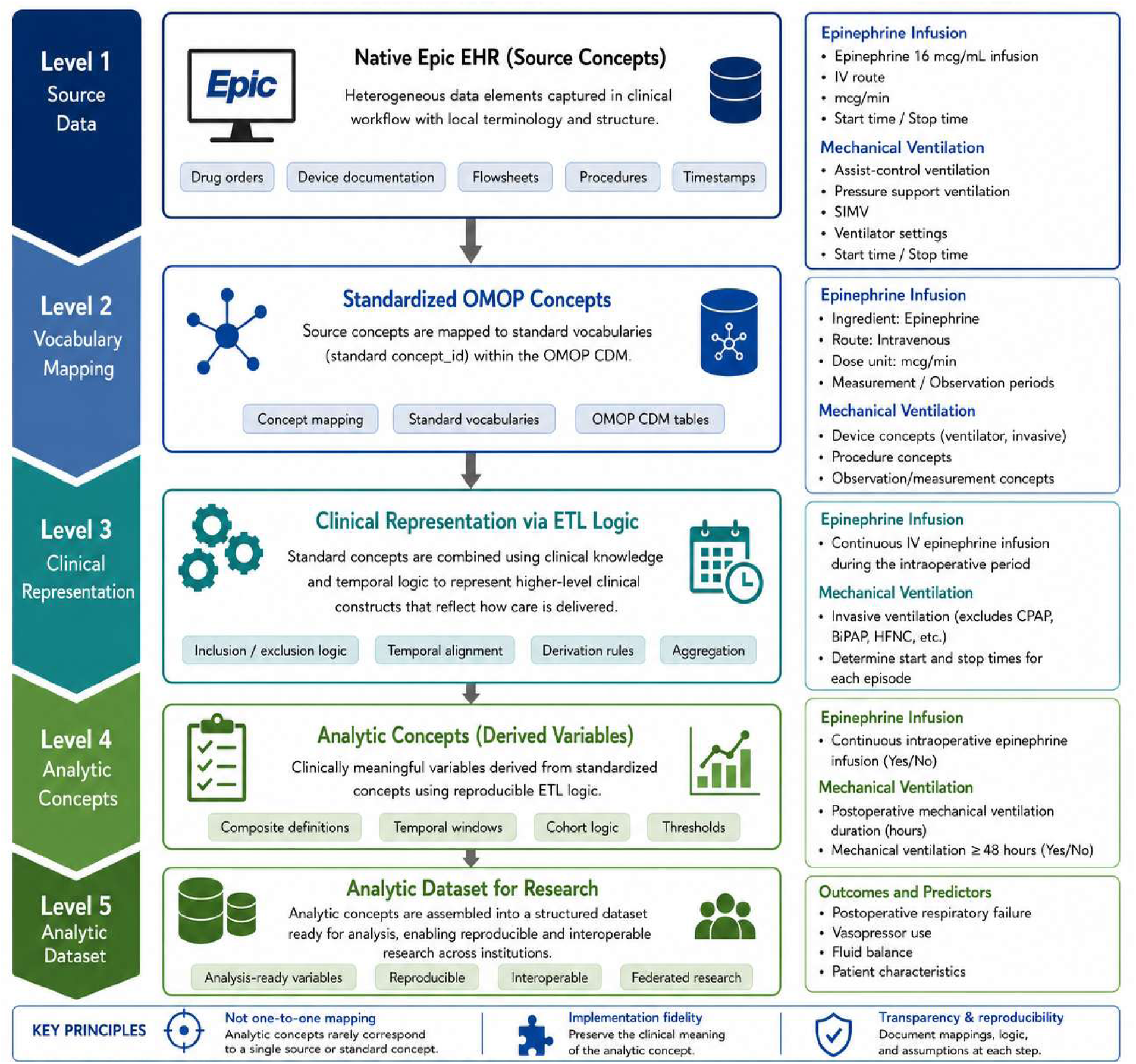
Conceptual Framework. Conceptual framework for analytic interoperability of perioperative electronic health record (EHR) data within the Observational Medical Outcomes Partnership (OMOP) Common Data Model (CDM). The framework distinguishes sequential levels of representation - from native source concepts to standardized OMOP concepts, clinical representations, derived analytic concepts, and an analysis-ready dataset. Clinically meaningful analytic variables are reconstructed from standardized concepts through reproducible clinical representation and ETL logic incorporating temporal relationships and clinical knowledge, enabling high implementation fidelity despite incomplete one-to-one vocabulary equivalence. ALT TEXT: Five-level framework showing transformation of native Epic perioperative EHR data through OMOP vocabulary mapping, clinical representation, and derived analytic concepts into a reproducible, interoperable, analysis-ready research dataset.

Data elements from the reference dataset were first identified as source concepts within the native Epic EHR and manually mapped to standard OMOP CDM concepts using the OHDSI Athena vocabulary browser (Version 1.15.6.58.260417.1529, OMOP Vocabulary version: v20260227).[24] Mapping analysis followed the Health Level 7 (HL7) Fast Healthcare Interoperability Resources (FHIR) ConceptMap equivalence framework[25] and prior EHR-to-OMOP mapping methodologies[18 19]. A detailed description of the mapping methodology, adjudication process, and governance procedures is provided in Supplementary Method 2. Source concepts were linked to standard OMOP concepts using standard concept identifiers. We analyzed mapping concordance using two concept map equivalence approaches. For our “strict” mapping analysis, defined as exact terminology and granularity, we used four categories: (1) equal – the definitions of the Epic and OMOP concepts were exactly the same; (2) wider – the OMOP mapping was wider in meaning than the source Epic concept and some granularity could be lost; (3) narrower – the OMOP mapping was more specific in meaning than the source Epic concept; and (4) unmatched – there was no match for the Epic concept in the OMOP code system. For the lenient analysis, mappings classified as wider or narrower under strict terminology and granularity criteria were re-evaluated according to whether the Epic and OMOP concepts preserved the same clinically relevant meaning for the intended analytic use. Mappings judged clinically equivalent despite terminology or granularity differences were classified as equal under the lenient criterion; mappings that did not preserve clinically relevant meaning retained their wider or narrower classification.

Throughout this study, source concepts refer to individual data elements extracted from the native Epic EHR, standard OMOP concepts refer to standardized vocabulary concepts representing those data elements, and analytic concepts refer to clinically meaningful variables derived from one or more standardized concepts for analysis.

Because many perioperative variables cannot be represented by a single standardized concept, mapped OMOP concepts served as building blocks for higher-level clinical representations. This approach reflects the distinction between vocabulary standardization and analytic concept reconstruction, recognizing that clinically meaningful variables frequently emerge from combinations of standardized concepts rather than individual source concepts. Representative clinical representation rules for each major analytic concept are summarized in Supplementary Table S1 and described in Supplementary Method 3. Reproducible ETL logic incorporating temporal relationships, inclusion and exclusion criteria, aggregation rules, and clinical knowledge was used to reconstruct clinically meaningful events, such as continuous vasopressor infusions and episodes of invasive mechanical ventilation. These clinical representations were subsequently transformed into analytic concepts, including binary exposure indicators, treatment durations, and threshold-based variables.

When an exact standard OMOP concept corresponding to a source medication formulation or concentration was unavailable, medications were mapped to the appropriate standard ingredient concept. Analytic variables were then reconstructed using administration route, dose units, timing, duration, and other contextual information. This approach prioritized preservation of the clinical meaning required for analysis rather than exact product-level equivalence.

Initial concept mappings and derivation logic were independently reviewed by two clinical informaticists (BP, OS) and a critical care nurse (JS). Discrepancies were adjudicated through multidisciplinary consensus involving two critical care physicians (NK, JA), a surgeon (GU), a critical care nurse (JS), and three clinical informaticists (BP, OS, AL) until a final mapping and derivation specification was achieved.

### ETL Implementation

Following vocabulary standardization, SQL-based ETL pipelines reconstructed standardized OMOP concepts into clinically meaningful clinical representations and analytic concepts within a project-specific OMOP database.[22] The workflow sequentially (1) extracted source data from Epic Clarity, (2) mapped source concepts to standard OMOP concepts, (3) integrated standardized concepts across OMOP tables, (4) applied temporal logic and clinical rules to reconstruct higher-level clinical representations, (5) derived analytic concepts using predefined cohort definitions, temporal windows, aggregation rules, and thresholds, and (6) assembled the final analysis-ready dataset. (Supplementary Figure S1).

Separate ETL modules were developed for cohort construction, perioperative timeline generation, clinical representation of exposures and outcomes, derivation of analytic variables, demographic characteristics, and predictor variables to facilitate validation, maintenance, and reuse. This modular architecture allowed vocabulary standardization, clinical representation, and analytic concept derivation to be validated independently before assembly of the final analysis-ready dataset. Additional details regarding the modular ETL architecture are provided in Supplementary Method 4.

ETL outputs were validated at multiple stages by comparing standardized concepts, reconstructed clinical representations, and derived analytic variables with the corresponding native Epic implementation to ensure fidelity to the original clinical workflow, consistent with prior anesthesia OMOP implementations.[10]

### Evaluation of Representation Fidelity

For this evaluation, concept equivalence refers to the relationship between Epic source concepts and standardized OMOP concepts according to the HL7 equivalence categories; cohort concordance refers to agreement in encounter inclusion between the native Epic and OMOP implementations; variable-level agreement refers to agreement in reconstructed analytic-variable values among encounters represented in both implementations; and implementation fidelity refers to the overall degree to which the OMOP implementation reproduced the native Epic analytic dataset.

To test fidelity, three complementary evaluations were performed. First, concept coverage was assessed by calculating the proportion of Epic concepts classified as equal, wider, narrower, or unmatched under both strict and lenient HL7 equivalence criteria. Mapping performance was additionally summarized across major clinical domains, including oxygen delivery, fluid administration, surgical specialty, perioperative timestamps, demographics, outcomes, and physiologic measurements. Second, implementation fidelity was evaluated by assessing cohort concordance between the native Epic and OMOP implementations and variable-level agreement for all derived analytic variables. Third, mapping discrepancies were systematically reviewed to determine whether differences between the Epic and OMOP implementations resulted from limitations of the OMOP vocabulary, structural differences between the Epic and OMOP data model, or implementation-specific ETL logic. Among encounters represented in both implementations, variable-level discrepancies were quantified overall and stratified by postoperative respiratory failure case status, and each discrepancy was evaluated according to its source, magnitude, and potential effect on analytic representation.

## RESULTS

### Reference Perioperative Dataset

The native Epic PRF reference dataset comprised 28 analytic variables (Supplementary Table S2) derived from 538 individual source concepts spanning perioperative timestamps, demographic characteristics, physiologic measurements, medications, fluid administration, operative characteristics, outcomes, and hospital utilization measures.

Representation of these variables within the OMOP CDM required standardization of source concepts followed by reconstruction of higher-level clinical representations and analytic concepts using reproducible derivation logic.

### Epic-to-OMOP Concept Mapping

Under strict equivalence criteria, 40.7% (n=219) of Epic-to-OMOP concept mappings were equal, 57.1% (n=306) were wider, 2.4% (n=13) were narrower, and 0% (n=0) were unmatched. Under lenient criteria, which permitted mappings with terminology or granularity differences to be classified as equal when clinically relevant meaning for the intended analytic use was preserved, 97.4% (n=524) were classified as equal, 2.0% (n=11) as wider, 0.6% (n=3) as narrower, and 0% (n=0) as unmatched. The complete source-to-concept mapping specification is provided in Supplementary Table S3.

Mapping performance varied across clinical domains. Among clinical domains with greater than 10 variables, using strict mapping criteria, flowsheet variables had the highest proportion of equal mappings (92.3%, n=12) and fluid intake and output had the lowest (20.5%, n=38), reflecting greater dependence on local implementation logic or approximation within the current OMOP vocabulary. (Table 1; Supplementary Table S4)

**Table 1.** Epic-to-OMOP Concept Mapping Performance by Clinical Domain.

| Clinical Domain | Source Concepts (n) | Equal % (Strict) | Equal % (Lenient) | Primary Reason for Strict Non-equivalence |
|---|---|---|---|---|
| Admission Source | 20 | 30.0% | 70.0% | OMOP CDM does not reliably differentiate transfers from external versus internal emergency departments |
| Admission Type | 6 | 100.0% | 100.0% | None |
| Anesthesia Class | 7 | 100.0% | 100.0% | None |
| Anesthesia Type | 1 | 100.0% | 100.0% | None |
| Drug Frequency | 2 | 0.0% | 0.0% | OMOP CDM does not represent medication dose frequency; continuous infusion identified using dose-unit time component |
| Drug Route | 3 | 100.0% | 100.0% | None |
| Ethnicity | 5 | 80.0% | 100.0% | OMOP CDM does not allow option of decline to state |
| Financial Class | 45 | 71.1% | 100.0% | Local financial classes mapped to standardized payer concepts |
| Fluids I/O | 185 | 20.5% | 97.8% | Fluid intake and output represented using multiple OMOP concepts and derived calculations; intake/output inferred from concept identifiers because OMOP has no dedicated intake/output field |
| Flowsheet | 13 | 92.3% | 100.0% | Institution-specific flowsheet values mapped to standard concepts |
| Medication | 66 | 54.5% | 100.0% | Local medication records standardized to ingredient-level OMOP concepts |
| Other (drip v push) | 3 | 66.7% | 100.0% | Administration type inferred from route and dose-unit information |
| Oxygen Device | 100 | 49.0% | 98.0% | Multiple standard concepts required to represent oxygen delivery and mechanical ventilation |
| Patient Type | 1 | 100.0% | 100.0% | None |
| Race | 26 | 23.1% | 100.0% | Local race values mapped to standard OMOP race concepts |
| Surgery Type | 1 | 100.0% | 100.0% | None |
| Surgical Specialty | 53 | 26.4% | 100.0% | Local OR service values mapped to standardized surgical specialty concepts |
| Tracheostomy | 1 | 100.0% | 100.0% | None |
| **TOTAL** | **538** | **40.7%** | **97.4%** | **Local implementation differences, concept granularity, and OMOP structural limitations** |

Note: Lenient equivalence permitted mappings classified as wider or narrower under strict terminology/granularity criteria to be classified as equal when clinically relevant meaning for the intended analytic use was preserved.

Because many perioperative variables did not correspond to individual standardized OMOP CDM concepts, reconstruction of higher-level clinical representations was required before analytic concepts could be derived. For example, continuous intraoperative vasopressor administration was reconstructed by integrating standard ingredient concepts with administration route, dose units, and temporal relationships rather than relying on product-specific drug concepts. Similarly, postoperative mechanical ventilation duration was derived by combining multiple standardized procedure and device concepts using predefined temporal logic. These implementations preserved the intended clinical meaning despite incomplete product-level vocabulary equivalence, demonstrating that clinically meaningful analytic variables could be reconstructed from standardized OMOP concepts.

### Analytic Concept Reconstruction

Application of the analytic interoperability framework produced a complete analysis- ready OMOP dataset containing cohort definitions, perioperative timelines, medication exposures, physiologic variables, operative characteristics, outcomes, and derived predictor variables. Reconstruction of these variables was achieved through standardized concepts combined with reproducible clinical representation and derivation logic. Details are provided in Supplementary Appendix A, and representation limitations are presented in Supplementary Appendix B.

Application of the study inclusion and exclusion criteria produced a final OMOP cohort of 31,169 surgical encounters compared with 31,158 encounters in the native Epic implementation. Overall cohort concordance exceeded 99%, with 31,083 surgical encounters represented in both datasets. None of the 161 encounters with discordant cohort membership were PRF casesAmong the 75 Epic-only encounters, 38 reflected differences in admission source mapping, with most remaining differences attributable to institutional ETL requirements for valid CPT codes. Among the 86 OMOP-only encounters, 78 reflected limitations in distinguishing external from internal emergency department transfers within the OMOP CDM (Figure 2; Supplementary Figure S2).

**Figure 2:**
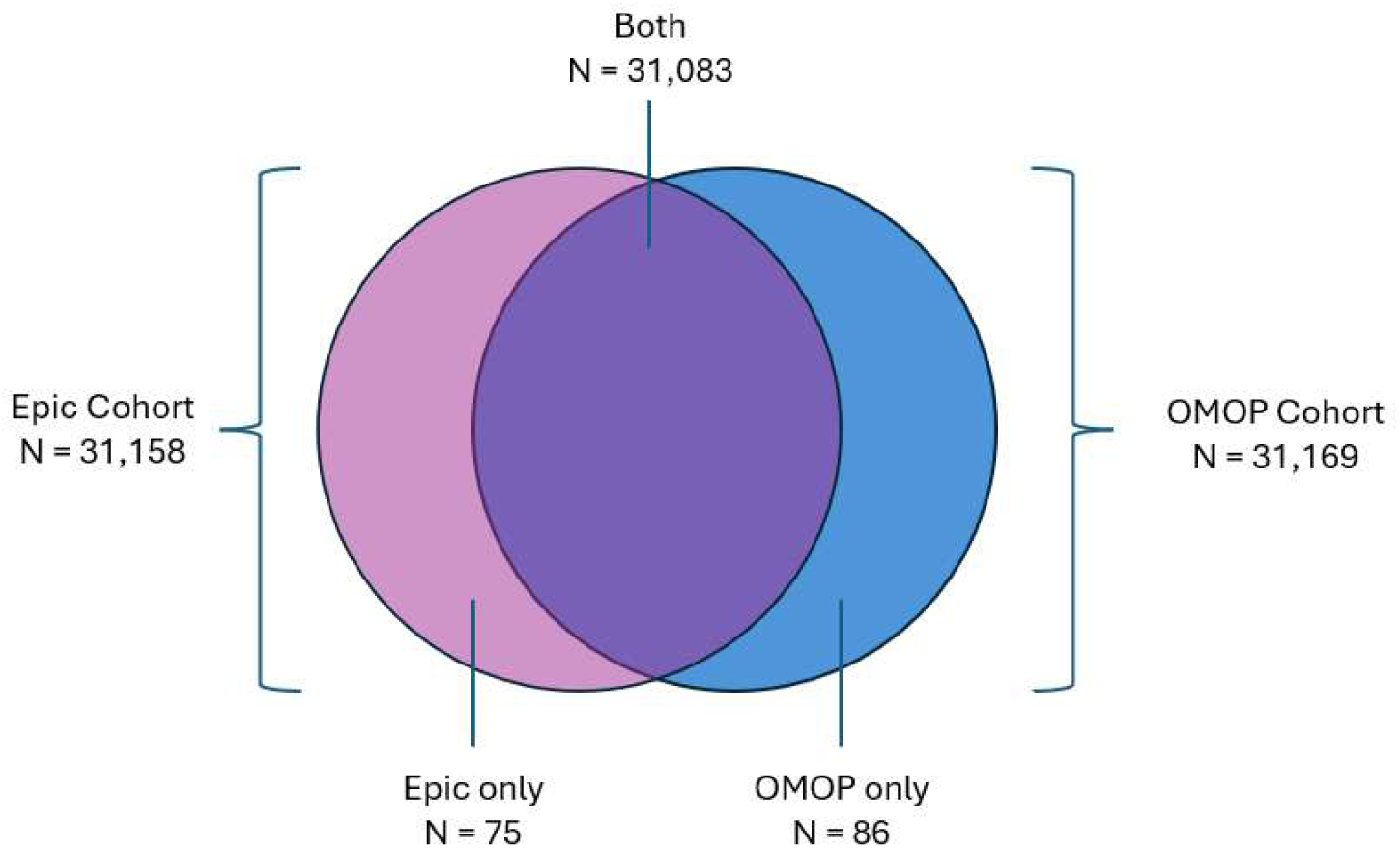
Cohort concordance between the native Epic and reconstructed OMOP postoperative respiratory failure (PRF) cohort. Application of identical cohort eligibility criteria produced 31,158 encounters in the native Epic implementation and 31,169 encounters in the OMOP implementation, with 31,083 encounters represented in both cohorts (>99% concordance). Epic-only encounters (n = 75) primarily reflected surgical encounters absent from the institutional OMOP instance because of existing ETL requirements for valid CPT codes (n = 36) or differences in admission source mapping (n = 38); one encounter lacked evidence of inpatient status (n = 1). OMOP-only encounters (n = 86) primarily reflected differences in representation of admission source, including encounters classified as external transfers in the OMOP implementation (n = 78), with the remaining discrepancies attributable to unidentified implementation differences (n = 8). ALT TEXT: Venn diagram showing >99% cohort concordance between native Epic and reconstructed OMOP implementations: 31,083 encounters overlap, with 75 Epic-only and 86 OMOP-only encounters.

### Representation Fidelity

Variable-level agreement was evaluated among the 31,083 surgical encounters represented in both datasets, including 264 postoperative respiratory failure cases and 30,819 controls. Agreement was high across nearly all reconstructed analytic concepts. Encounter-level discrepancies, including their frequency overall and by case status, magnitude, source, and potential effect on analytic representation, are detailed in Supplementary Table S5. These findings demonstrate that preservation of clinically meaningful analytic variables was achievable despite incomplete one-to-one vocabulary equivalence. All perioperative timestamps - including hospital admission and discharge, operating room entry and exit, anesthesia start and stop times, and derived operative durations - demonstrated complete or near-complete agreement between the native Epic and OMOP implementations. Likewise, postoperative respiratory failure status, mechanical ventilation duration, ASA physical status, physiologic measurements, operative characteristics, and hospital outcomes demonstrated either complete agreement or only minor numeric differences attributable primarily to SQL date-time calculations or floating-point precision that did not meaningfully affect the resulting analytic variables.

### Sources of Representation Differences

Among the 31,083 encounters represented in both implementations, meaningful analytic-variable differences were limited to three areas: continuous vasopressor infusion identification, BMI selection, and Medicare payer assignment. A fourth area, admission source, contributed to discordant cohort membership.

Continuous vasopressor infusion differed because the native Epic implementation classified continuous administration using medication frequency metadata, whereas the OMOP implementation inferred continuous administration from standardized dose units and administration records. These disparate EHR source data elements contained conflicting information for four (0.01%) encounters.

Body mass index differences resulted from selection of different qualifying height and weight observations when multiple measurements were available. Differences in categorical BMI values were observed for 81 (0.26%) encounters.

Medicare payer discrepancies arose when multiple temporally overlapping payer records existed during a hospitalization. Inability to apply exact payer selection rules led to differences among 142 (0.46%) encounters.

Admission source differences reflected limitations in representation of admission source within the OMOP CDM, which prevented reliable differentiation of transfers from external versus internal emergency departments and accounted for most discordant cohort assignments – 38 encounters included in Epic but excluded in OMOP and 78 encounters included in OMOP but excluded in Epic.

Meaningful analytic-variable differences among encounters represented in both implementations affected fewer than 1% of encounters and primarily reflected limitations of the current OMOP CDM vocabulary and structure, such as inability to explicitly represent medication administration frequency or associate a payer record directly to a hospital encounter, rather than errors in concept mapping or analytic concept reconstruction. Encounter-level analytic-variable discrepancies are summarized in Supplementary Table S5, additional details regarding sources of representation differences are provided in Supplementary Appendix B, and admission source mapping is illustrated in Supplementary Figure S3.

## DISCUSSION

This study demonstrates that complex perioperative electronic health record (EHR) data can be represented within the Observational Medical Outcomes Partnership (OMOP) Common Data Model (CDM) while preserving the clinically meaningful analytic concepts required for perioperative research. Although only 40.7% of Epic source concepts mapped directly to equal OMOP concepts under strict equivalence criteria, reconstruction of higher-level clinical representations and analytic concepts using standardized, reproducible clinical representation and ETL logic achieved greater than 99% cohort concordance with the native Epic implementation and preserved nearly all analytic variables required for postoperative respiratory failure research. These findings demonstrate that analytic interoperability extends beyond vocabulary mapping, requiring standardized concepts to be combined through transparent derivation logic that faithfully represents the underlying clinical workflow.

The conceptual framework presented in this study distinguishes four related but distinct layers of interoperability: source concepts, standardized OMOP concepts, clinical representations, and analytic concepts. Previous evaluations of OMOP implementation have emphasized vocabulary mapping, semantic harmonization, structural implementation, and data quality across heterogeneous databases.[3 9 18 19 21 22] Our findings demonstrate that these approaches, while essential, incompletely capture the ability of a common data model to support clinically meaningful perioperative research because many perioperative variables do not correspond to individual standardized concepts. Instead, standardized concepts function as reusable building blocks from which higher-level clinical representations and ultimately analytic concepts are reconstructed.

Continuous vasopressor administration and postoperative mechanical ventilation illustrate this distinction. In both cases, the clinically meaningful analytic variable did not correspond to any single standardized OMOP concept. Rather, ingredient concepts, device concepts, temporal relationships, administration characteristics, and other standardized data elements were integrated through reproducible derivation logic to reconstruct the intended clinical representation. Representative reconstruction of continuous vasopressor infusion episodes is illustrated in Supplementary Figure S4, while reconstruction of invasive mechanical ventilation episodes from standardized OMOP concepts is illustrated in Supplementary Figure S5. The complete standardized concept set used for mechanical ventilation reconstruction is provided in Supplementary Appendix C. Consequently, preservation of clinically meaningful analytic variables was achieved despite incomplete product- or device-level vocabulary equivalence. These examples demonstrate that successful interoperability depends less on exact one-to-one concept mapping than on transparent reconstruction of clinically meaningful analytic concepts.

Our findings suggest that implementation fidelity is a more informative measure of interoperability than vocabulary equivalence alone. Comparative evaluations of common data models have emphasized content coverage, terminology support, and structural implementation,[8] while other authors have cautioned that transformation of heterogeneous EHR data into standardized models may result in loss of clinically meaningful information when local concepts or relationships cannot be adequately represented.[26] Our findings extend these observations by demonstrating that, for complex perioperative research, preservation of analytic meaning can frequently be achieved through reconstruction of clinical representations and analytic concepts from standardized OMOP concepts using transparent, reproducible derivation logic.

Consequently, interoperability should be evaluated according to preservation of analytically equivalent clinical variables rather than vocabulary equivalence alone.

These findings extend prior OMOP implementation studies. Garza et al.[8] demonstrated broad OMOP content coverage while noting tradeoffs between standardization and preservation of source information. Lamer et al.[10 14] demonstrated transformation of French EHR and anesthesia data, including additional structures for computed perioperative concepts, while our prior critical care work identified limitations in one-to- one OMOP concept coverage.[9] The present study extends this literature by showing that preservation of clinically meaningful analytic variables depends on standardized vocabularies plus explicit, reproducible reconstruction of clinical representations and analytic concepts.

Meaningful representation differences were uncommon and primarily reflected structural limitations of the OMOP CDM - such as inability to distinguish external from internal emergency department transfers or explicitly represent medication infusion frequency - rather than failures of the mapping methodology or analytic reconstruction framework.

Similar implementation-specific differences have been reported in other large OMOP transformation studies, reflecting the practical challenges of harmonizing heterogeneous EHR systems rather than failures of the underlying common data model.[12 14-16]

Similar to previous single-network and institution-specific OMOP implementation studies,[10 13 15] our evaluation reflects a single institutional Epic configuration and a project-specific OMOP implementation. Because the OMOP implementation and validation were conducted within the same institution and with overlapping investigator expertise, the high observed implementation fidelity may partly reflect shared knowledge of the source data, clinical workflow, and reference analytic definitions. Evaluation across additional institutions, EHR implementations, and existing OMOP networks will therefore be necessary to determine the transportability of the proposed analytic interoperability framework. This study evaluated fidelity of cohort construction and analytic concept reconstruction rather than equivalence of downstream statistical analyses. Whether the remaining implementation differences affect model coefficients, calibration, discrimination, or other inferential results requires separate evaluation and is the subject of ongoing work. A major strength of this work is the comprehensive validation of the complete analytic implementation rather than evaluation of vocabulary mappings alone. By comparing cohort construction, reconstructed clinical representations, derived analytic concepts, and sources of discordance against the native Epic implementation, we demonstrate that high implementation fidelity can be achieved despite incomplete one-to-one concept equivalence. This distinction is particularly important for investigators implementing the OMOP CDM for secondary use research because it emphasizes preservation of analytic meaning rather than vocabulary completeness as the principal objective.

Perioperative OMOP implementation remains resource intensive. Concept mapping, analytic definitions, ETL implementation, validation, and manuscript development required considerable effort across study investigators over 18 months. Furthermore, maintenance will also be needed as local EHR implementations and OMOP vocabularies evolve. Clinical and informatics expertise were both essential to preserve semantic meaning and translate it into standardized concepts and reproducible derivation logic. This investment produced reusable definitions and workflows for future perioperative studies. Future research into methods that improve the efficiency of this work, such as development of AI-assisted mapping and analytical concept derivation, may help to lower resource and support barriers that currently limit broader use.

More broadly, the fundamental unit of interoperability for complex perioperative research is the analytic concept, not the individual standardized vocabulary concept. Standard vocabularies provide a common semantic foundation, but interoperable research depends on transparent, reproducible transformation of standardized concepts into clinically meaningful analytic variables. Figure 1 formalizes this hierarchy, which may extend to other complex domains where analytic variables emerge from combinations of standardized concepts. This approach complements the OHDSI vision of distributed observational research by extending interoperability from standardized vocabularies to reproducible analytic concept definitions.[4]

Analytic interoperability also has implications beyond observational research. A recent electronic quality measure for postoperative respiratory failure showed that structured perioperative EHR data can support scalable quality measurement across hospitals and EHR vendors.[1] Our findings show that the underlying analytic concepts can also be represented reproducibly in OMOP, supporting interoperable prediction, quality measurement, and other secondary uses.

Analytic interoperability for perioperative research requires reconstruction of clinically meaningful analytic concepts from standardized OMOP concepts through reproducible representation and implementation logic, not one-to-one vocabulary mapping alone.

Future work should test transportability across distributed OMOP networks and develop reusable perioperative analytic concept libraries for multicenter federated analyses.[2 4 6] Better representation of medication administration, perioperative workflows, and admission source would reduce custom implementation. Reusable analytic concept definitions and implementation logic may ultimately be as important for multicenter perioperative research as further vocabulary harmonization.

## CONCLUSION

This study demonstrates that high-fidelity representation of complex perioperative EHR data within the OMOP Common Data Model is achievable through reconstruction of clinically meaningful analytic concepts rather than one-to-one vocabulary mapping. By emphasizing preservation of analytic meaning, the proposed analytic interoperability framework provides a practical methodology for evaluating standardized data models and supporting scalable multicenter research across clinically complex domains.

## Supporting information

Supplemental Matter

## Data Availability

All data produced in the present study are available upon reasonable request to the authors

## Author Contributions

Stocking: conceptualization, data curation, formal analysis, funding acquisition, investigation, methodology, validation, visualization, writing – original draft, writing – review and editing

Stewart - conceptualization, data curation, formal analysis, methodology, validation, visualization, writing – original draft, writing – review and editing

Utter: conceptualization, formal analysis, investigation, methodology, validation, visualization, writing – original draft, writing – review and editing

Adams: conceptualization, methodology, supervision, writing – review and editing Kenyon: conceptualization, methodology, supervision, writing – review and editing

Sandrock: conceptualization, methodology, writing – review and editing Albertson: conceptualization, methodology, writing – review and editing Garcia: conceptualization, methodology, writing – review and editing

Abraham: conceptualization, formal analysis, methodology, supervision, writing – original draft, writing – review and editing

## Acknowledgments

Anna Liu and Brian Paciotti PhD, Enterprise Analytics and Data Services (EADS), UC Davis Health, Information Technology for their assistance with data curation and initial Epic to OMOP mappings.

Cindy X. Cai, MD, Hopkins Medicine and Sally Baxter, MD UC San Diego Health for their assistance with OMOP CDM familiarization and mapping.

## Competing Interests

None

## Funding

JCS National Institutes of Health, National Heart, Lung, and Blood Institute K01HL168222

**Tables (1 – within text) and Figures (2 - submitted as separate documents)**

**Figures:** (listed here – Figure Legend uploaded as a separate document)

**Total Main Text Figures**: 2

**Supplementary Materials: (submitted as separate documents)**

## Supplementary Methods

1. Supplementary Method 1. Conceptual framework and guiding principles for analytic interoperability
2. Supplementary Method 2. Epic-to-OMOP mapping methodology, adjudication process, and governance procedures
3. Supplementary Method 3. Clinical representation and analytic concept derivation methodology
4. Supplementary Method 4. Modular ETL architecture

## Supplementary Tables

1. Supplementary Table S1. Representative clinical representation rules for major analytic concepts
2. Supplementary Table S2. Reference perioperative analytic dataset
3. Supplementary Table S3. Complete Epic source-to-OMOP concept mapping specification
4. Supplementary Table S4. Epic-to-OMOP mapping performance by clinical domain
5. Supplementary Table S5. Validation of analytic concept reconstruction between native Epic and OMOP implementations

## Supplementary Figures

1. Supplementary Figure S1. ETL workflow for analytic concept reconstruction
2. Supplementary Figure S2. Comparison of cohort inclusion between native Epic and OMOP implementations
3. Supplementary Figure S3. Admission source mapping and representation differences
4. Supplementary Figure S4. Reconstruction of continuous vasopressor infusion episodes
5. Supplementary Figure S5. Reconstruction of invasive mechanical ventilation episodes from standardized OMOP concepts

## Supplementary Appendices

1. Supplementary Appendix A. Analytic concept reconstruction details
2. Supplementary Appendix B. Representation limitations and sources of representation differences
3. Supplementary Appendix C. Standardized OMOP concept set for mechanical ventilation reconstruction

## Notes

### Competing Interest Statement

The authors have declared no competing interest.

### Author Declarations

IRB of UC Davis gave approval (IRB #822917) with waiver of consent and waiver of HIPAA authorization.

