## Supplemental Matter for "Analytic Interoperability Framework for High-Fidelity Representation of Perioperative Electronic Health Record Data in the OMOP Common Data Model Using Clinical Representation and Analytic Concept Reconstruction"

**Supplementary Materials**

**Supplementary Methods**

1. Supplementary Method 1. Conceptual framework and guiding principles for analytic interoperability
2. Supplementary Method 2. Epic-to-OMOP mapping methodology, adjudication process, and governance procedures
3. Supplementary Method 3. Clinical representation and analytic concept derivation methodology
4. Supplementary Method 4. Modular ETL architecture

**Supplementary Tables**

1. Supplementary Table S1. Representative clinical representation rules for major analytic concepts
2. Supplementary Table S2. Reference perioperative analytic dataset
3. Supplementary Table S3. Complete Epic source-to-OMOP concept mapping specification
4. Supplementary Table S4. Epic-to-OMOP mapping performance by clinical domain
5. Supplementary Table S5. Validation of analytic concept reconstruction between native Epic and OMOP implementations

**Supplementary Figures**

1. Supplementary Figure S1. ETL workflow for analytic concept reconstruction
2. Supplementary Figure S2. Comparison of cohort inclusion between native Epic and OMOP implementations
3. Supplementary Figure S3. Admission source mapping and representation differences
4. Supplementary Figure S4. Reconstruction of continuous vasopressor infusion episodes
5. Supplementary Figure S5. Reconstruction of invasive mechanical ventilation episodes from standardized OMOP concepts

**Supplementary Appendices**

1. Supplementary Appendix A. Analytic concept reconstruction details
2. Supplementary Appendix B. Representation limitations and sources of representation differences
3. Supplementary Appendix C. Standardized OMOP concept set for mechanical ventilation reconstruction

**Total supplementary material: 4 methods, 5 tables, 5 figures, and 3 appendices (17 items).**

### Supplementary Method 1. Analytic Concept Development Framework

The objective of this study was not simply to map native EHR data to standardized OMOP CDM vocabulary concepts, but to preserve the clinically meaningful variables required for perioperative research. To achieve this, we developed a hierarchical framework for analytic concept development that distinguishes the sequential transformation of native EHR data into analysis-ready variables (Figure 1, main manuscript). The framework consists of five conceptual levels: source concepts, standardized OMOP concepts, clinical representations, analytic concepts, and the analysis-ready dataset. Each level serves a distinct purpose in preserving clinical meaning while enabling standardized, reproducible analyses.

#### Level 1: Source Concepts

Source concepts are the original data elements recorded within the native EHR during routine clinical care. These concepts reflect local clinical workflows and institutional documentation practices and may include medications, device documentation, procedure records, physiologic measurements, timestamps, flowsheet entries, and other structured EHR data. Source concepts often use local terminology, are highly granular, and may and frequently lack direct semantic equivalence with standardized vocabularies. Because source concepts originate from operational clinical systems, they should be regarded as representations of clinical documentation rather than analytically meaningful variables. Consequently, multiple source concepts often contribute to a single clinical construct.

#### Level 2: Standardized OMOP Concepts

Source concepts are translated into standard OMOP vocabulary concepts using established terminology mappings. Standardization provides a common semantic representation across institutions and permits data from heterogeneous EHR systems to be represented within a common data model. Importantly, standardization alone does not guarantee preservation of analytic meaning. Individual source concepts may map directly to equivalent OMOP CDM concepts, to broader or narrower concepts, require approximation when no exact standard concept exists, or be unmappable. Furthermore, many clinically meaningful perioperative variables cannot be represented by a single standardized concept. Instead, standardized concepts serve as reusable semantic building blocks for subsequent analytic reconstruction.

#### Level 3: Clinical Representations

Clinical representations combine one or more standardized OMOP CDM concepts using reproducible ETL and clinical logic to reconstruct higher-order clinical events that reflect patient care. This process incorporates temporal relationships, inclusion and exclusion criteria, aggregation rules, and domain-specific clinical knowledge. For example, continuous intraoperative vasopressor administration cannot be represented solely by an individual drug concept. Instead, it is reconstructed by integrating standard ingredient concepts with administration route, dose-unit information, and timing relative to the operative period. Likewise, episodes of invasive mechanical ventilation are reconstructed by combining standardized procedure and device concepts with temporal overlap rules to identify the onset and duration of mechanical ventilation. Clinical representations therefore bridge the gap between standardized terminology and clinically meaningful events.

#### Level 4: Analytic Concepts

Analytic concepts are the derived variables used for statistical analysis, predictive modeling, and observational research. These variables are generated from one or more clinical representations using predefined derivation rules, temporal windows, threshold definitions, or aggregation methods. Examples include binary indicators of intraoperative vasopressor exposure, postoperative mechanical ventilation duration, fluid balance during specified perioperative intervals, and binary postoperative respiratory failure outcomes. Unlike standardized concepts, analytic concepts correspond directly to the variables included in statistical models and therefore represent the primary unit of analytic interoperability. Because analytic concepts are reconstructed from standardized concepts rather than mapped directly from source concepts, multiple institutions can generate analytically equivalent variables despite differences in local documentation practices or vocabulary mappings.

**Level 5: Analysis-Ready Dataset**

The final level consists of an integrated analysis-ready dataset assembled from all derived analytic concepts. At this stage, cohort definitions, demographic characteristics, exposures, outcomes, and predictor variables have been reconstructed using standardized, reproducible implementation logic and are suitable for downstream statistical analysis or machine learning. Separation of analytic concept derivation from dataset assembly promotes modularity, allowing individual analytic concepts to be validated, reused, or updated without requiring redevelopment of the complete analytic dataset. This modular approach also facilitates transparent reporting of derivation logic and supports reproducible implementation across institutions.

**Guiding Principles**

Three principles underlie the analytic concept development framework. First, analytic concepts rarely correspond to a single source or standardized concept. Rather, clinically meaningful variables typically emerge from combinations of standardized concepts linked through temporal relationships and clinical knowledge. Second, implementation fidelity is the primary objective of interoperability. The success of an OMOP implementation should therefore be evaluated according to its ability to preserve clinically meaningful analytic concepts rather than by vocabulary equivalence alone. Third, transparency and reproducibility are essential throughout the transformation process. Documentation of concept mappings, derivation rules, temporal logic, and implementation assumptions allows analytic concepts to be reproduced consistently across institutions while preserving local data governance.

Together, these five conceptual levels and guiding principles provide a general framework for transforming heterogeneous EHR data into interoperable analytic datasets and emphasize that successful interoperability is achieved through reproducible analytic concept reconstruction rather than one-to-one vocabulary mapping alone. Implementation of this conceptual framework is described in Supplementary Methods 2–4. The complete Epic-to-OMOP mapping specification is provided in Supplementary Table S1, representative clinical representation rules are summarized in Supplementary Table S3, and the operational ETL workflow is illustrated in Supplementary Figure S1.

### Supplementary Method 2. Epic-to-OMOP Concept Mapping Methodology

#### Overview

Epic-to-OMOP concept mapping was performed to establish a standardized semantic foundation for subsequent reconstruction of clinically meaningful analytic concepts. Consistent with the conceptual framework described in Supplementary Method 1 and depicted in Figure 1, concept mapping was regarded as the process of translating native Epic source concepts into standardized OMOP vocabulary concepts rather than the final representation of analytic variables. Consequently, standardized concepts served as reusable semantic building blocks for downstream clinical representation and analytic concept derivation. The mapping process consisted of five sequential stages: (1) source concept identification, (2) vocabulary mapping, (3) assessment of concept equivalence, (4) multidisciplinary review and adjudication, and (5) governance and documentation of final mappings.

#### Source Concept Identification

All source concepts required to recreate the native postoperative respiratory failure (PRF) analytic dataset were first identified within the Epic Clarity database. Source concepts included structured EHR elements spanning perioperative timestamps, medication orders and administrations, physiologic measurements, device documentation, procedures, demographic characteristics, outcomes, and hospital utilization variables. Rather than mapping only the final analytic variables, each underlying source concept contributing to an analytic variable was identified independently. This approach allowed analytic variables composed of multiple source concepts to be reconstructed using standardized OMOP concepts while preserving the original clinical intent.

#### Standard Vocabulary Mapping

Each source concept was manually mapped to a standard OMOP concept using the OHDSI Athena vocabulary browser (Version 1.15.6.58.260417.1529; OMOP Vocabulary version v20260227). Standard concepts were preferentially selected whenever available, following OMOP conventions regarding valid concepts, standard concept identifiers, and domain assignment. Where multiple candidate concepts existed, concept selection prioritized preservation of the intended clinical meaning rather than lexical similarity. Mapping decisions considered concept definitions, hierarchical relationships, domain assignments, and expected use within the OMOP CDM. When an exact standard concept corresponding to a local Epic concept was unavailable, the closest clinically appropriate standard concept was selected for use in subsequent analytic reconstruction. For medications, this frequently involved mapping locally defined formulations or concentrations to standard ingredient concepts while preserving administration characteristics, including route, dose units, timing, and duration, through additional clinical representation logic.

#### Assessment of Concept Equivalence

Concept equivalence was evaluated using the Health Level 7 (HL7) Fast Healthcare Interoperability Resources (FHIR) ConceptMap relationship framework.

Each Epic-to-OMOP mapping was classified as one of four relationship types:

- **Equal:** the Epic source concept and standard OMOP concept represented identical clinical meaning.
- **Wider:** the selected OMOP concept represented a broader clinical concept than the Epic source concept.
- **Narrower:** the selected OMOP concept represented a more specific clinical concept than the Epic source concept.

- **Unmatched:** no clinically appropriate standard OMOP concept was available.

To distinguish semantic equivalence from analytic equivalence, mappings were evaluated using both **strict** and **lenient** criteria. Under the **strict** definition, only mappings demonstrating exact semantic correspondence between the Epic source concept and the standard OMOP concept were classified as equivalent. Any reduction or increase in clinical specificity resulted in classification as narrower or wider. Under the **lenient** definition, concepts were considered equivalent when they adequately represented the intended clinical meaning required for the analytic variable, even when differences existed in terminology, formulation, or granularity. This distinction recognizes that many clinically meaningful analytic variables do not require exact product-level or workflow-level equivalence to preserve analytic validity. For example, a locally defined epinephrine infusion concentration may not possess an exact standard OMOP equivalent. However, mapping the source concept to the standard epinephrine ingredient concept while preserving administration route, dose units, and timing permits accurate reconstruction of the intended analytic concept (continuous intraoperative epinephrine infusion).

**Multidisciplinary Review and Adjudication**

Initial mappings were independently reviewed by two clinical informaticists (BP, OS) and a critical care nurse (JS) with expertise in perioperative clinical documentation and OMOP implementation. Mappings requiring interpretation of clinical workflow, local documentation practices, or vocabulary ambiguity were subsequently reviewed through multidisciplinary consensus involving two critical care physicians (NK, JA), a surgeon (GU), a critical care nurse (JS), and three clinical informaticists (BP, OS, AL). Review focused on preservation of clinical intent rather than vocabulary similarity alone. For each mapping, reviewers considered whether the selected standardized concept would support accurate reconstruction of the intended clinical representation and downstream analytic concept. When disagreement occurred, consensus was achieved through iterative review of the source documentation, OMOP vocabulary definitions, and anticipated analytic use until a final mapping decision was reached.

**Mapping Governance**

All mapping decisions were documented in a master mapping specification document that served as the authoritative reference throughout implementation. Each record included the source concept, corresponding standard OMOP CDM concept identifier, relationship classification, rationale for concept selection, and any implementation notes required for analytic concept reconstruction. Because standardized concept mapping represented only one component of the overall implementation framework, governance extended beyond vocabulary selection to include documentation of assumptions, derivation rules, temporal logic, and known limitations affecting analytic concept development. Maintaining this comprehensive mapping specification ensured consistency across ETL development, validation, and future refinement of analytic concepts while providing a transparent record of implementation decisions.

**Relationship to Analytic Concept Development**

The mapping process described above established the semantic foundation for analytic interoperability but did not itself generate the final analytic variables. As described in Supplementary Method 1, standardized OMOP concepts were subsequently combined using reproducible clinical representation logic and predefined derivation rules to reconstruct clinically meaningful analytic concepts. Consequently, the success of the implementation depended not only on vocabulary mapping but also on transparent documentation of how standardized concepts were transformed into analysis-ready variables. The complete mapping specification is provided in Supplementary Table S1, while domain-specific mapping performance is summarized in Supplementary Table S2. Transformation of standardized concepts into clinical representations is described in Supplementary Method 3.

**Supplementary Method 3. Clinical Representation Rules**

**Overview**

Many clinically meaningful perioperative variables cannot be represented by individual standardized OMOP concepts. Instead, they emerge from the integration of multiple standardized concepts using temporal relationships, aggregation rules, inclusion and exclusion criteria, and clinically informed derivation logic. We refer to these intermediate constructs as clinical representations, which bridge standardized vocabulary concepts and analysis-ready variables.

The clinical representation rules described below document how standardized OMOP concepts were combined to reconstruct the analytic concepts used in the postoperative respiratory failure (PRF) dataset. Unlike source concept mappings, these rules are independent of any particular EHR implementation and therefore represent reusable implementation specifications for analytic interoperability.

**Supplementary Method 3, Table 1. Clinical representation rules for major analytic concepts.**

| Analytic Concept | Primary Source Concepts | Standard OMOP Concepts | Clinical Representation | Derived Analytic Variable |
|---|---|---|---|---|
| Continuous vasopressor infusion | Medication orders and medication administration records | Standard drug ingredient concepts, administration route concepts, dose-unit concepts | Standard drug concepts were integrated with administration route, dose units containing a time component (e.g., mcg/min), and temporal overlap with the intraoperative period to identify continuous intravenous vasopressor administration. | Binary indicator (Yes/No) |
| Invasive mechanical ventilation episode | Device documentation, ventilator settings, respiratory procedures | Standard device concepts and procedure concepts representing invasive mechanical ventilation episode duration derived from recorded start and end times | Consecutive qualifying invasive ventilation concepts were combined into ventilation episodes using temporal overlap. Non-invasive ventilation concepts (CPAP, BiPAP, HFNC, etc.) were excluded. | Ventilation episode duration (hours) |

| | | | | |
|---|---|---|---|---|
| Invasive mechanical ventilation duration | Mechanical ventilation episode | Clinical representation above | Duration calculated from episode start and stop times after anesthesia end. Multiple episodes were summed when appropriate. | Cumulative ventilation episode duration (hours)Duration (hours) |
| Postoperative respiratory failure | Mechanical ventilation duration | Clinical representation above | Patients with cumulative postoperative invasive mechanical ventilation ≥48 hours were classified as PRF cases. | Binary outcome (Case/Control) |
| Perioperative fluid intake | Intake documentation | Standard measurement and observation concepts representing administered fluids (measurement and observation domains were required to capture volume values) | Fluid administration records were aggregated during predefined perioperative time windows. | Volume (L) |
| Perioperative fluid output | Output documentation | Standard measurement and observation concepts representing urine output, blood loss, drains, etc. (measurement and observation domains were required to capture volume values) | Output records aggregated within predefined perioperative windows. | Volume (L) |
| Net fluid balance | Fluid intake and output | Clinical representations above | Net fluid balance calculated as intake minus output for each analytic window. | Continuous variable |
| Anesthesia duration | Anesthesia procedure records | General anesthesia procedure concept | Difference between standardized anesthesia start and stop timestamps. | Duration (hours) |
| Surgery duration | Procedure records, OR timestamps | Standard procedure concepts | Earliest qualifying procedure start (incision time) through latest qualifying procedure end (incision closed), with OR timestamps used when | Duration (hours) |

| | | | | |
|---|---|---|---|---|
| | | | procedure timestamps were unavailable. | |
| Tracheostomy status | Condition concepts | Presence of tracheostomy | At hospital admission and OR wheels out, used as an exclusion criteria for ENT specialty surgeries | Binary indicator (Yes/No) |
| Perioperative physiologic measurements | Physiologic observations | Standard measurement concepts | Measurements occurring within predefined intraoperative windows were summarized using median values. | Continuous variables (heart rate, $FiO_2$, tidal volume, end-tidal $CO_2$, etc.) |
| Body mass index | BMI, height, weight measurements | Standard measurement concepts | Existing BMI measurement preferentially selected. If unavailable, BMI calculated from the most recent qualifying height and weight measurements before anesthesia. | Continuous BMI and BMI category |
| Medicare payer | Insurance coverage records | Standard payer concepts | Coverage overlapping the operative hospitalization was evaluated, with predefined rules for selecting the primary payer when multiple records existed. | Binary indicator (Yes/No) |
| Surgical specialty | Operating room documentation | Standard observation concepts | Standardized operating room service concepts mapped to analytic specialty categories. | Categorical variable |
| ICU length of stay | ICU visit records | Visit detail concepts | Consecutive ICU visit intervals aggregated into cumulative ICU stay, accounting for operative periods occurring between ICU encounters. | Duration (days) |

**Temporal Logic**

Temporal relationships were fundamental to clinical representation and were applied consistently across analytic concepts. Rather than relying solely on the presence of standardized concepts, derivation rules incorporated the timing of events relative to perioperative milestones, including hospital admission, operating room entry and exit, anesthesia start and end, and hospital discharge.

Representative temporal windows included:

- Intraoperative: anesthesia start through anesthesia end.
- Immediate postoperative: anesthesia end through 24 hours postoperatively.
- Hospitalization: admission through discharge.
- Mechanical ventilation outcome: cumulative invasive ventilation occurring after completion of anesthesia.

These standardized temporal definitions ensured that analytic concepts reflected clinical workflow rather than isolated observations.

**Aggregation Rules**

Several analytic concepts required aggregation of multiple standardized observations before derivation.

Aggregation methods included:

- summation (e.g., fluid intake and output),
- cumulative duration (e.g., mechanical ventilation),
- median values (e.g., physiologic measurements),
- earliest or latest qualifying timestamp (e.g., operative events),
- binary indicators generated from occurrence of one or more qualifying events (e.g., vasopressor administration).

Aggregation rules were specified before implementation and applied consistently across all encounters.

**Clinical Decision Rules**

Certain analytic concepts required clinically informed decision rules that could not be determined from standardized vocabulary concepts alone.

Examples included:

- distinguishing continuous intravenous vasopressor infusions from intermittent medication administration using dose-unit and administration information;
- classifying invasive versus non-invasive mechanical ventilation despite multiple underlying device concepts;
- identifying the most clinically appropriate body mass index measurement when multiple qualifying observations were available;
- selecting the primary payer when multiple overlapping payer records existed during a hospitalization.

These rules were developed collaboratively by clinicians and clinical informaticists to preserve the intended clinical meaning of each analytic concept.

**Reproducibility**

All clinical representation rules were implemented using deterministic logic and version-controlled ETL workflows. Each analytic concept was developed independently of downstream statistical analyses, permitting modular validation, reuse, and future refinement without requiring redevelopment of the complete analytic dataset.

By separating vocabulary mapping from clinical representation, the framework enables institutions using different EHR implementations to reconstruct analytically equivalent variables from standardized OMOP concepts while preserving local documentation practices. This distinction forms the basis of the analytic interoperability framework proposed in this study. Representative implementations of these clinical representation rules are illustrated in Supplementary Figures S4 and S5. Detailed analytic concept definitions for each reconstructed variable are provided in Supplementary Appendix A, while the complete standardized OMOP CDM concept set used to reconstruct invasive mechanical ventilation is listed in Supplementary Appendix C. Together, these materials constitute a reference implementation supporting the analytic interoperability framework described in the main manuscript.

### Supplementary Method 4. ETL Architecture

#### Overview

The extract-transform-load (ETL) architecture was designed to operationalize the analytic interoperability framework described in Figure 1 by separating vocabulary standardization from clinical representation and analytic concept derivation. Rather than implementing a single monolithic ETL process, the workflow was organized into modular components corresponding to major stages of analytic concept development. This modular architecture facilitated independent validation, maintenance, reuse, and future extension while preserving transparency throughout the implementation process.

**Supplementary Method 4, Figure 1: Summary of the overall ETL workflow**

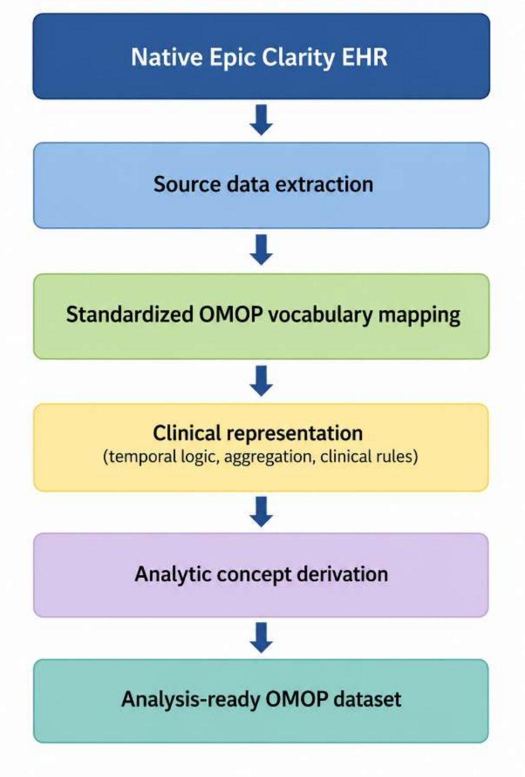


Each stage produces standardized outputs that serve as inputs for the subsequent stage, enabling analytic concepts to be reconstructed independently of the native EHR implementation.

#### Source Extraction Module

The source extraction module identified and extracted all structured Epic Clarity data required to reconstruct the reference perioperative dataset. Source data included demographic information, hospital and surgical encounters, perioperative timestamps, medication administrations, physiologic measurements, device documentation, procedures, outcomes, and hospital utilization variables. Only structured EHR data were included in this implementation. Source concepts retained their original clinical context prior to vocabulary standardization.

**Vocabulary Standardization Module**
Extracted source concepts were translated into standard OMOP concepts using the mapping methodology described in Supplementary Method 2. Standardization established a common semantic representation while preserving the relationship between original source concepts and their corresponding OMOP concepts. This stage intentionally performed only semantic standardization. No analytic variables were derived during vocabulary mapping. Outputs from this module consisted of standardized OMOP concepts distributed across the appropriate OMOP clinical tables according to OMOP domain conventions.

**Clinical Representation Module**

The clinical representation module reconstructed higher-order clinical events from combinations of standardized OMOP concepts. This stage incorporated temporal relationships, aggregation methods, inclusion and exclusion criteria, and clinically informed decision rules to represent how care was delivered rather than how individual observations were documented.

Representative clinical representations included:
- perioperative episodes defined relative to anesthesia and surgery;
- invasive mechanical ventilation episodes reconstructed from multiple device and procedure concepts;
- continuous vasopressor administration reconstructed from drug ingredient, administration route, dose-unit, and temporal information;
- perioperative fluid administration reconstructed through aggregation of standardized intake and output concepts.

These intermediate clinical representations formed the foundation for subsequent analytic variable derivation.

**Analytic Concept Derivation Module**

Derived analytic concepts were generated from clinical representations using predefined derivation rules, temporal windows, aggregation methods, and threshold definitions. Unlike standardized concepts, analytic concepts correspond directly to variables included in statistical analyses.

Representative analytic concepts included:
- binary vasopressor exposure indicators;
- duration of postoperative mechanical ventilation;
- aggregation of perioperative fluid balance;
- operative and anesthesia durations;
- physiologic summary measures;
- categorization of continuous body mass index measurement;
- postoperative respiratory failure classification based on threshold definition of duration of postoperative mechanical ventilation.

Each analytic concept was generated independently using deterministic implementation logic, allowing individual variables to be validated and modified without affecting unrelated portions of the analytic dataset.

**Analysis-Ready Dataset Assembly**

The final module assembled all derived analytic concepts into a single analysis-ready dataset suitable for statistical modeling and observational research. At this stage, cohort definitions, exposures, outcomes, demographic characteristics, and predictor variables had been reconstructed using standardized implementation rules. Dataset assembly represented the final step in the analytic interoperability framework and contained no additional clinical transformation beyond integration of validated analytic concepts.

**Modular Implementation Components**
To improve maintainability and facilitate independent validation, the ETL architecture was organized into six functional modules.

**Cohort Construction Module**
Applied study inclusion and exclusion criteria to standardized OMOP data to identify the eligible surgical cohort. Cohort definitions incorporated demographic criteria, operative characteristics, hospitalization requirements, admission characteristics, and exclusion conditions.

**Perioperative Timeline Module**
Reconstructed clinically meaningful perioperative time intervals from standardized timestamps. This module generated the temporal framework used throughout subsequent analytic concept derivation, including anesthesia periods, operative periods, and postoperative observation windows.

**Exposure Module**
Generated standardized representations of perioperative exposures, including medication administration, fluid administration, physiologic measurements, and operative characteristics. Clinical representation rules were applied to transform standardized concepts into clinically meaningful exposure variables.

**Outcome Module**
Constructed postoperative outcome variables using predefined clinical representation rules. Outcomes included postoperative respiratory failure status, duration of invasive mechanical ventilation, hospital length of stay, and intensive care unit length of stay.

**Demographic Module**
Generated demographic and administrative variables including age, sex, race, ethnicity, insurance status, and body mass index using standardized OMOP concepts and predefined selection rules.

**Predictor Module**
Derived all relevant perioperative predictor variables required for the postoperative respiratory failure analytic dataset. Predictor derivation integrated outputs from the timeline, exposure, demographic, and outcome modules to generate the complete set of variables used for subsequent statistical analyses.

**Validation Strategy**
Validation was performed at each stage of the ETL architecture rather than only after completion of the final analytic dataset. Vocabulary mappings, reconstructed clinical representations, derived analytic concepts, and cohort definitions were independently compared with the corresponding native Epic implementation to ensure preservation of clinical meaning throughout the transformation process. This staged validation approach enabled discrepancies to be

localized to individual implementation modules, facilitating efficient refinement while maintaining overall analytic fidelity.

**Design Principles**

Three architectural principles guided implementation. First, modularity separated vocabulary standardization, clinical representation, and analytic concept derivation into independent components, permitting validation and reuse of each stage. Second, deterministic implementation ensured that identical standardized inputs consistently generated identical analytic outputs, supporting reproducibility across repeated executions and future implementations. Third, transparency was maintained by documenting all transformation logic, assumptions, and derivation rules rather than embedding them within implementation code. Consequently, the architecture can be reproduced using alternative software platforms while preserving equivalent analytic concepts. Validation of the resulting implementation is summarized in Supplementary Table S5, while sequential cohort derivation is illustrated in Supplementary Figure S2.

**Supplementary Table S1. Clinical Representation Rules**

**Purpose:** Documents how standardized concepts become clinically meaningful variables.

| Analytic Variable | Standard OMOP Concepts | Inclusion Rules | Temporal Logic | Aggregation Method | Derived Variable | Threshold / Output |
|---|---|---|---|---|---|---|
| Continuous vasopressor infusion | Drug ingredient + IV route + dose units | IV administration only | Between anesthesia start and end | Any qualifying infusion | Binary | Yes/No |
| Mechanical ventilation duration | Device + procedure concepts | Invasive ventilation only | After anesthesia end | Sum of qualifying episodes | Continuous | Hours |
| OR fluid intake | Measurement + observation concepts | Intake concepts only | During anesthesia | Sum | Continuous | Liters |
| OR fluid output | Measurement + observation concepts | Output concepts only | During anesthesia | Sum | Continuous | Liters |
| OR fluid balance | Fluid intake/output | None | During anesthesia | Intake − Output | Continuous | Liters |
| BMI | BMI OR height + weight | Closest pre-op measurement | Before anesthesia | BMI or calculated | Continuous | kg/m² |
| Medicare payer | Payer records | Overlapping hospitalization | During Admission | Closest qualifying payer | Binary | Yes/No |
| Surgical specialty | Observation concepts | OR service | Surgery | Category assignment | Categorical | Specialty |
| Heart rate | Measurement | HR concepts | During anesthesia | Median | Continuous | bpm |
| $FiO_2$ | Measurement | $FiO_2$ concepts | During anesthesia | Median | Continuous | % |
| Tidal volume | Measurement | TV concepts | During anesthesia | Median | Continuous | mL |
| End-tidal $CO_2$ | Measurement | $ETCO_2$ concepts | During anesthesia | Median | Continuous | mmHg |

**Supplementary Table S2. Analytic Variable Dictionary**

| Column Name | Column Description |
|---|---|
| CASE_CONTROL | OUTCOME VARIABLE 1=case; 0=non-case |
| AGE | Patient's age at the time of admission. |
| SEX_BIN | 0=Male; 1=Female; 2=Other (non-binary, decline to state, other) |
| RACE_BIN | 0=White; 1=Black (African American or Black); 2=Asian (Asian; Native Hawaiian or other Pacific Islander; American Indian or Alaska Native); 3=Other (Other; Multiracial); 4=Unknown (Unable to respond; Unavailable or unknown; Decline to State; Blank). |
| ETHNICITY_BIN | 0=non-Hispanic; 1=Hispanic; 2=Unknown (Declined to state; Unavailable or Unknown; Blank) |
| PRIMARY_PAYOR_BIN | 0= Commercial; 1= M-Care;2= M-Cal; 3=Other (All Other; Other Public, Self Pay, Unknown, Blank) |
| SURG_SPECIALTY_BIN | SURG_SPECIALTY:<br>0=Cardiovascular: ACS, CVT, Cardiac Surgery, Cardiology, Peds CVT, Vascular, Pulmonary<br>1=Ortho (non-spine): Ortho Foot & Ankle, Ortho Hand, Ortho Joint, Ortho Sports, Ortho Trauma, Orthopedics, Peds Orthopedics, Community Orthopedics<br>2=Neurosurgery, Spine: Neuro Spine, Neurosurgery, Ortho spine, Peds Neurosurgery<br>3=Oncology: Gynecologic Oncology, Ortho Oncology, Peds Hem Onc, Surgical Oncology<br>4=EENT: Dental, ENT, Ophthalmology<br>5=Other: Pain management<br>6=General Surgery: GEN SURG, Colo-Rectal, FEMS (Bari/FG), Peds GI, Peds Surgery, Thoracic Surgery, Transplant, Plastics, Burn<br>7=Urology and gynecology: Urogynecology, Urology, Obstetrics/GYN, Family Planning |
| BMI | The calculated body mass index (BMI) using the height and weight reported in this data extract. |
| BMI_CAT | 0 = Underweight: < 18.5; 1 = Normal weight: 18.5 - 24.9; 2 = Overweight:25 - 29.9; 3 = Obese1: 30 - 34.9; 4 = Obese2: 35 - 39.9; 5 = Morbidly obese: >40 |

| | |
|---|---|
| ASA_CLASS | The American Society of Anesthesiologists (ASA) rating category number for the procedure. |
| ASA_CLASS_ABN | 0= ASA class of 1 and 2 and 1=ASA class3, 4, 5 |
| VASO_CONT_INF_YN_OR | Flag to indicate whether any vasopressors are administered via continuous infusion during the surgery: 1=Yes; 0=No |
| ANES_DUR | Duration of time from anesthesia start date/time to anesthesia end date/time in hours. |
| SURG_DUR | Duration of time from incision start time to incision closed time in hours |
| HR_MED_OR | The median heart rate recorded during the surgery (from anesthesia start to anesthesia stop) |
| ETCO2_MED_OR | The median ETCO2 recorded during the surgery (from anesthesia start to anesthesia stop) |
| VTCC_SET_MED_OR | The median VTCC Set recorded during the surgery (from anesthesia stat to anesthesia stop) |
| FIO2_MED_OR | The median FIO2 recorded during the surgery (from anesthesia stat to anesthesia stop) |
| POST_OR_INTAKE_LITER | Total fluid intake, excluding irrigant total, during the surgery (from anesthesia start to anesthesia stop) |
| POST_OR_OUTPUT_LITER | Total fluid output, excluding irrigant total, during the surgery (from anesthesia start to anesthesia stop) |
| POST_OR_NET_LITER | Net fluid balance total, total fluid intake - total fluid output, during the surgery (from anesthesia start to anesthesia stop) |
| HAS_TRACHEOSTOMY_AT_ADMSN | Flag to indicate whether patient has a Tracheostomy airway at the time of hospital admission (retained as an analytic variable and applied as an exclusion criterion only for ENT specialty surgeries). |
| HAS_TRACHEOSTOMY_AT_OR_WHEELS_WO | Flag to indicate whether patient has a Tracheostomy airway at the time of OR wheels out (retained as an analytic variable and applied as an exclusion criterion only for ENT specialty surgeries). |
| MORTALITY | 0=no; 1=yes |
| DISCHARGE_DISP_GROUPER_BIN | 0=Home: HOME 1=Died: DEATH, DEATH - HOSPICE READMIT, HOSPICE, HOSPICE READMIT 2=Other Facility: ACUTE TRANSFER, LONG TERM ACUTE |

| | |
|---|---|
| | CARE, PSYCH, REHAB, SNF 3=Other: AMA, LAW ENF, OTHER |
| LOS_OBS | Hospital length of stay in days |
| ICU_DAYS_OBS_ICUFILE | ICU length of stay in days |
| POD_DX | Post operative day number that patient has total post-operative vent time of 48 hours or greater with surgery date = day 0 |

**Supplemental Table 3: Epic to OMOP Concept Mapping**

| Row | Category Level 1 | Category Level 2 | Category Level 3 | Epic Concept Flowsheet Name | Epic Concept Flowsheet Value | Mapping Granularity STRICT | Mapping Granularity CLINCAL INTENT (LENIENT) | Explanation for STRICT Mapping not "equal" | OMOP Concept ID | OMOP Concept Code | OMOP Concept Name | Standard (S) or Non-Standard (NS) Concept | OMOP Domain ID | OMOP Concept Class ID | OMOP Vocabulary ID |
|---|---|---|---|---|---|---|---|---|---|---|---|---|---|---|---|
| 1 | Admission Type | n/a | n/a | Emergency Admission | n/a | equal | equal | n/a | 32203 | 1 | Emergency - The patient required immediate medical intervention as a result of severe, life threatening, or potentially disabling conditions. Generally, the patient was admitted through the emergency room. | S | Observation | UB04 Pri Typ of Adm | UB04 Pri Typ of Adm |
| 2 | Admission Type | n/a | n/a | Urgent Admission | n/a | equal | equal | n/a | 32204 | 2 | Urgent - The patient required immediate attention for the care and treatment of a physical or mental disorder. Generally, the patient was admitted to the first available and suitable accommodation. | S | Observation | UB04 Pri Typ of Adm | UB04 Pri Typ of Adm |
| 3 | Admission Type | n/a | n/a | Elective Admission | n/a | equal | equal | n/a | 32205 | 3 | Elective - The patient's condition permitted adequate time to schedule the availability of suitable accommodations. | S | Observation | UB04 Pri Typ of Adm | UB04 Pri Typ of Adm |
| 4 | Admission Type | n/a | n/a | Newborn | n/a | equal | equal | n/a | 32206 | 4 | Newborn - Necessitates the use of special source of admission codes. | S | Observation | UB04 Pri Typ of Adm | UB04 Pri Typ of Adm |
| 5 | Admission Type | n/a | n/a | Trauma | n/a | equal | equal | n/a | 32207 | 5 | Trauma Center - visits to a trauma center/hospital as licensed or designated by the State or local government authority authorized to do so, or as verified by the American College of Surgeons and involving a trauma activation. | S | Observation | UB04 Pri Typ of Adm | UB04 Pri Typ of Adm |
| 6 | Admission Type | n/a | n/a | Not Applicable | n/a | equal | equal | n/a | 32208 | 9 | Information not available | S | Observation | UB04 Pri Typ of Adm | UB04 Pri Typ of Adm |
| 7 | Admission Source | n/a | n/a | Non-Health Care Facility | n/a | narrower | narrower | adds type specificity | 581476 | OMOP4822459 | Home Visit | S | Visit | Visit | Visit |
| 8 | Admission Source | n/a | n/a | Physician Referral | n/a | narrower | narrower | adds type specificity | 581476 | OMOP4822459 | Home Visit | S | Visit | Visit | Visit |
| 9 | Admission Source | n/a | n/a | Clinic Referral | n/a | equal | equal | n/a | 38004207 | 261Q00000X | Clinic/Center | S | Visit | Visit | NUCC |
| 10 | Admission Source | n/a | n/a | SITE Clinic Referral | n/a | wider | equal | missing site specificity | 38004207 | 261Q00000X | Clinic/Center | S | Visit | Visit | NUCC |
| 11 | Admission Source | n/a | n/a | Transfer From a Hospital | n/a | wider | equal | missing inpatient specificity | 8717 | 21 | Inpatient Hospital | S | Visit | Visit | CMS Place of Service |
| 12 | Admission Source | n/a | n/a | Transfer from Skilled Nursing (SNF), Intermediate | n/a | wider | equal | missing intermediate specificity | 8863 | 31 | Skilled Nursing Facility | S | Visit | Visit | CMS Place of Service |
| 13 | Admission Source | n/a | n/a | Transfer from Another Health Care Facility | n/a | equal | equal | n/a | 32197 | 6 | Transfer from another Healthcare facility | NS | Visit | UB04 Point of Origin | UB04 Point of Origin |
| 14 | Admission Source | n/a | n/a | Court Law Enforcement | n/a | narrower | narrower | adds type specificity | 38003619 | 9 | Prison / Correctional Facility | S | Visit | Visit | CMS Place of Service |
| 15 | Admission Source | n/a | n/a | Ext Facility OP Procedure | n/a | wider | wider | missing site specificty | 32199 | 9 | Information not available | NS | Visit | UB04 Point of Origin | UB04 Point of Origin |
| 16 | Admission Source | n/a | n/a | Information Not Available | n/a | equal | equal | n/a | 32199 | 9 | Information not available | NS | Visit | UB04 Point of Origin | UB04 Point of Origin |
| 17 | Admission Source | n/a | n/a | Laboratory Outreach | n/a | wider | wider | missing site specificity | 32199 | 9 | Information not available | NS | Visit | UB04 Point of Origin | UB04 Point of Origin |
| 18 | Admission Source | n/a | n/a | Rehab Center | n/a | wider | equal | missing rehab center specificity | 8717 | 21 | Inpatient Hospital | S | Visit | Visit | CMS Place of Service |
| 19 | Admission Source | n/a | n/a | Transfer from SITE Health Hospital | n/a | wider | equal | missing transfer form site specificity | 8717 | 21 | Inpatient Hospital | S | Visit | Visit | CMS Place of Service |
| 20 | Admission Source | n/a | n/a | SITEAcute to SITEInpatient Hospice | n/a | wider | equal | missing transfer, acute, and inpatient specificity | 8717 | 21 | Inpatient Hospital | S | Visit | Visit | CMS Place of Service |
| 21 | Admission Source | n/a | n/a | Transfer from Ambulatory Surgery | n/a | wider | wider | missing transfer, ambulatory surgery specificity | 38004207 | 261Q00000X | Clinic/Center | S | Visit | Visit | NUCC |
| 22 | Admission Source | n/a | n/a | Transfer from a Hospice Facility | n/a | wider | equal | missing transfer specificity | 8546 | 34 | Hospice | S | Visit | Visit | CMS Place of Service |
| 23 | Admission Source | n/a | n/a | Emergency Room | n/a | equal | equal | n/a | 9203 | ER | Emergency Room Visit | S | Visit | Visit | Visit |
| 24 | Admission Source | n/a | n/a | Transfer From an Emergency Room | n/a | wider | equal | missing transfer | 9203 | ER | Emergency Room Visit | S | Visit | Visit | Visit |
| 25 | Admission Source | n/a | n/a | Born In This Hospital | n/a | equal | equal | n/a | 32592 | S | Born inside this hospital | NS | Visit | UB04 Point of Origin | UB04 Point of Origin |
| 26 | Admission Source | n/a | n/a | Born Outside of This Hospital | n/a | equal | equal | n/a | 32593 | T | Born outside this hospital | NS | Visit | UB04 Point of Origin | UB04 Point of Origin |
| 27 | Patient type | n/a | n/a | Inpatient status at anytime during addmission | n/a | equal | equal | n/a | 4205408 | 308540004 | Inpatient stay | S | Observation | Clinical Finding | SNOMED |
| 28 | Surgery type | n/a | n/a | Elective surgery | n/a | equal | equal | n/a | 4314435 | 8715000 | Hospital admission, elective | S | Observation | Procedure | SNOMED |
| 29 | Anesthesia type | n/a | n/a | General anesthesia | n/a | equal | equal | n/a | 4171773 | 274507007 | Operative general anesthesia | S | Procedure | Procedure | SNOMED |
| 30 | Tracheostomy | n/a | n/a | Tracheostomy | n/a | equal | equal | n/a | 4201387 | 302108003 | Tracheostomy present | S | Condition | Clinical Finding | SNOMED |
| 31 | Ethnicity | n/a | n/a | Hispanic or Latino | n/a | equal | equal | n/a | 38003563 | Hispanic | Hispanic or Latino | S | Ethnicity | Ethnicity | Ethnicity |
| 32 | Ethnicity | n/a | n/a | Not Hispanic or Latino | n/a | equal | equal | n/a | 38003564 | Not Hispanic | Not Hispanic or Latino | S | Ethnicity | Ethnicity | Ethnicity |
| 33 | Ethnicity | n/a | n/a | Decline to state | n/a | wider | equal | missing reason is decline | 8552 | UNK | Unknown | S | Race (non-standard) | Race | Race |
| 34 | Ethnicity | n/a | n/a | Null | n/a | equal | equal | n/a | 8552 | UNK | Unknown | S | Race (non-standard) | Race | Race |
| 35 | Ethnicity | n/a | n/a | Unavailable or unknown | n/a | equal | equal | n/a | 8552 | UNK | Unknown | S | Race (non-standard) | Race | Race |
| 36 | Anesthesia Classification System | Observation | n/a | ASA | n/a | equal | equal | n/a | 4159411 | 273270000 | American Society of Anesthesiologists physical status classification | S | Measurement | Staging / Scales | SNOMED |
| 37 | Anesthesia Classification System | n/a | n/a | ASA 1 | n/a | equal | equal | n/a | 4186042 | 413495001 | American Society of Anesthesiologists physical status class 1 | S | Condition | Staging / Scales | SNOMED |
| 38 | Anesthesia Classification System | n/a | n/a | ASA 2 | n/a | equal | equal | n/a | 4184967 | 413496000 | American Society of Anesthesiologists physical status class 2 | S | Condition | Staging / Scales | SNOMED |
| 39 | Anesthesia Classification System | n/a | n/a | ASA 3 | n/a | equal | equal | n/a | 4186043 | 413497009 | American Society of Anesthesiologists physical status class 3 | S | Condition | Staging / Scales | SNOMED |
| 40 | Anesthesia Classification System | n/a | n/a | ASA 4 | n/a | equal | equal | n/a | 4211334 | 413498004 | American Society of Anesthesiologists physical status class 4 | S | Condition | Staging / Scales | SNOMED |
| 41 | Anesthesia Classification System | n/a | n/a | ASA 5 | n/a | equal | equal | n/a | 4186044 | 413499007 | American Society of Anesthesiologists physical status class 5 | S | Condition | Staging / Scales | SNOMED |
| 42 | Anesthesia Classification System | n/a | n/a | ASA 6 | n/a | equal | equal | n/a | 4186045 | 413500003 | American Society of Anesthesiologists physical status class 6 | S | Condition | Staging / Scales | SNOMED |
| 43 | drug_freq | n/a | n/a | CONTINUOUS INFUSION | n/a | wider | wider | missing specificity - continuous v push | 45884925 | LA11258-3 | Infusion, intravenous | S | Meas Value | Answer | LOINC |
| 44 | drug_freq | n/a | n/a | CONTINUOUS INFUSION PRN | n/a | wider | wider | missing specificity - continuous v push | 45884925 | LA11258-3 | Infusion, intravenous | S | Meas Value | Answer | LOINC |
| 45 | drug_route | n/a | n/a | IV | n/a | equal | equal | n/a | 4112421 | 255560000 | Intravenous | S | Observation | Qualifier Value | SNOMED |
| 46 | drug_route | n/a | n/a | IV INFUSION | n/a | equal | equal | n/a | 45884925 | LA11258-3 | Infusion, intravenous | S | Meas Value | Answer | LOINC |
| 47 | drug_route | n/a | n/a | IV (intraop) | n/a | equal | equal | n/a | 45884925 | LA11258-3 | Infusion, intravenous | S | Meas Value | Answer | LOINC |
| 48 | Financial Class | n/a | n/a | Commercial | n/a | equal | equal | n/a | 327 | 5 | (Commercial) Private Health Insurance | S | Payer | Payer | SOPT |
| 49 | Financial Class | n/a | n/a | Blue Shield | n/a | equal | equal | n/a | 327 | 5 | (Commercial) Private Health Insurance | S | Payer | Payer | SOPT |
| 50 | Financial Class | n/a | n/a | Blue Cross | n/a | equal | equal | n/a | 327 | 5 | (Commercial) Private Health Insurance | S | Payer | Payer | SOPT |
| 51 | Financial Class | n/a | n/a | MEDI-CAL/NON-CAPITAT | n/a | equal | equal | n/a | 289 | 2 | Medicaid | S | Payer | Payer | SOPT |
| 52 | Financial Class | n/a | n/a | MCAL (CAPITATED) | n/a | equal | equal | n/a | 289 | 2 | Medicaid | S | Payer | Payer | SOPT |
| 53 | Financial Class | n/a | n/a | CCS/GHPP | n/a | wider | equal | mising program specificity | 408 | 389 | Federal, State, Local not specified - Other | S | Payer | Payer | SOPT |
| 54 | Financial Class | n/a | n/a | CMSP | n/a | wider | equal | mising program specificity | 408 | 389 | Federal, State, Local not specified - Other | S | Payer | Payer | SOPT |
| 55 | Financial Class | n/a | n/a | OTHER COUNTY MIA | n/a | wider | equal | mising program specificity | 404 | 379 | Local Government FFS/Indemnity not otherwise specified (other local, county) | S | Payer | Payer | SOPT |
| 56 | Financial Class | n/a | n/a | OTHER SPONSOR | n/a | wider | equal | mising program specificity | 345 | 92 | Other Non-government | S | Payer | Payer | SOPT |
| 57 | Financial Class | n/a | n/a | RESEARCH GRANTS-DO NOT USE | n/a | equal | equal | n/a | 442 | 85 | Research/Donor | S | Payer | Payer | SOPT |
| 58 | Financial Class | n/a | n/a | SACTO COUNTY | n/a | wider | equal | mising program specificity | 404 | 379 | Local Government FFS/Indemnity not otherwise specified (other local, county) | S | Payer | Payer | SOPT |
| 59 | Financial Class | n/a | n/a | MEDICAREFFS | n/a | equal | equal | n/a | 280 | 1 | Medicare | S | Payer | Payer | SOPT |
| 60 | Financial Class | n/a | n/a | Medigap | n/a | equal | equal | n/a | 327 | 5 | (Commercial) Private Health Insurance | S | Payer | Payer | SOPT |
| 61 | Financial Class | n/a | n/a | CHAMPUS (MILITARY) | n/a | equal | equal | n/a | 296 | 311 | DoD TRICARE, Civilian Health and Medical Program of the Uniformed Services (CHAMPUS) | S | Payer | Payer | SOPT |
| 62 | Financial Class | n/a | n/a | OTHER INSURANCE | n/a | wider | equal | mising program specificity | 345 | 92 | Other Non-government | S | Payer | Payer | SOPT |
| 63 | Financial Class | n/a | n/a | PPO/HMO INS PLANS | n/a | equal | equal | n/a | 327 | 5 | (Commercial) Private Health Insurance | S | Payer | Payer | SOPT |
| 64 | Financial Class | n/a | n/a | MCARE RISK PLNS(HMO) | n/a | equal | equal | n/a | 280 | 1 | Medicare | S | Payer | Payer | SOPT |
| 65 | Financial Class | n/a | n/a | SHRINERS | n/a | wider | equal | mising program specificity | 345 | 92 | Other Non-government | S | Payer | Payer | SOPT |
| 66 | Financial Class | n/a | n/a | NON-SPONSORED-DO NOT USE | n/a | wider | equal | mising program specificity | 345 | 92 | Other Non-government | S | Payer | Payer | SOPT |
| 67 | Financial Class | n/a | n/a | PEND MCAL/CMSP/CMISP | n/a | wider | equal | mising program specificity | 408 | 389 | Federal, State, Local not specified - Other | S | Payer | Payer | SOPT |
| 68 | Financial Class | n/a | n/a | WORKERS COMPENSATION | n/a | equal | equal | n/a | 447 | 95 | Worker's Compensation | S | Payer | Payer | SOPT |
| 69 | Financial Class | n/a | n/a | CTS DISCOUNT | n/a | wider | equal | mising program specificity | 345 | 92 | Other Non-government | S | Payer | Payer | SOPT |
| 70 | Financial Class | n/a | n/a | PATIENT LIABILITY | n/a | equal | equal | n/a | 436 | 81 | Self-Pay | S | Payer | Payer | SOPT |
| 71 | Financial Class | n/a | n/a | Other | n/a | wider | equal | mising program specificity | 345 | 92 | Other Non-government | S | Payer | Payer | SOPT |
| 72 | Financial Class | n/a | n/a | CONVERSION DEFAULT | n/a | wider | equal | mising program specificity | 345 | 92 | Other Non-government | S | Payer | Payer | SOPT |
| 73 | Financial Class | n/a | n/a | PATIENT INSTALLMENTS | n/a | equal | equal | n/a | 436 | 81 | Self-Pay | S | Payer | Payer | SOPT |
| 74 | Financial Class | n/a | n/a | BAD DEBT - CMRE | n/a | equal | equal | n/a | 441 | 83 | Refusal to Pay/Bad Debt | S | Payer | Payer | SOPT |

| | | | | | | | | | | | | | | | |
|---|---|---|---|---|---|---|---|---|---|---|---|---|---|---|---|
| 75 | Financial Class | n/a | n/a | CONVERTED BAD DEBT | n/a | equal | equal | n/a | 441 | 83 | Refusal to Pay/Bad Debt | S | Payer | Payer | SOPT |
| 76 | Financial Class | n/a | n/a | BAD DEBT PRELIST | n/a | equal | equal | n/a | 441 | 83 | Refusal to Pay/Bad Debt | S | Payer | Payer | SOPT |
| 77 | Financial Class | n/a | n/a | BAD DEBT MEDICARE | n/a | equal | equal | n/a | 280 | 1 | Medicare | S | Payer | Payer | SOPT |
| 78 | Financial Class | n/a | n/a | BAD DEBT - USCB | n/a | equal | equal | n/a | 441 | 83 | Refusal to Pay/Bad Debt | S | Payer | Payer | SOPT |
| 79 | Financial Class | n/a | n/a | BAD DEBT-ASSET | n/a | equal | equal | n/a | 441 | 83 | Refusal to Pay/Bad Debt | S | Payer | Payer | SOPT |
| 80 | Financial Class | n/a | n/a | CHARITY CARE-DO NOT USE | n/a | equal | equal | n/a | 438 | 821 | Charity | S | Payer | Payer | SOPT |
| 81 | Financial Class | n/a | n/a | Medicare Replacement | n/a | equal | equal | n/a | 280 | 1 | Medicare | S | Payer | Payer | SOPT |
| 82 | Financial Class | n/a | n/a | DK Regional | n/a | equal | equal | n/a | 327 | 5 | (Commercial) Private Health Insurance | S | Payer | Payer | SOPT |
| 83 | Financial Class | n/a | n/a | Managed Care | n/a | equal | equal | n/a | 327 | 5 | (Commercial) Private Health Insurance | S | Payer | Payer | SOPT |
| 84 | Financial Class | n/a | n/a | Medicaid Replacement | n/a | equal | equal | n/a | 289 | 2 | Medicaid | S | Payer | Payer | SOPT |
| 85 | Financial Class | n/a | n/a | Medicare | n/a | equal | equal | n/a | 280 | 1 | Medicare | S | Payer | Payer | SOPT |
| 86 | Financial Class | n/a | n/a | Medicaid | n/a | equal | equal | n/a | 289 | 2 | Medicaid | S | Payer | Payer | SOPT |
| 87 | Financial Class | n/a | n/a | Self-pay | n/a | equal | equal | n/a | 436 | 81 | Self-Pay | S | Payer | Payer | SOPT |
| 88 | Financial Class | n/a | n/a | Worker's Comp | n/a | equal | equal | n/a | 447 | 95 | Worker's Compensation | S | Payer | Payer | SOPT |
| 89 | Financial Class | n/a | n/a | Tricare | n/a | equal | equal | n/a | 296 | 311 | DoD TRICARE, Civilian Health and Medical Program of the Uniformed Services (CHAMPUS) | S | Payer | Payer | SOPT |
| 90 | Financial Class | n/a | n/a | Champva | n/a | equal | equal | n/a | 296 | 311 | DoD TRICARE, Civilian Health and Medical Program of the Uniformed Services (CHAMPUS) | S | Payer | Payer | SOPT |
| 91 | Financial Class | n/a | n/a | Group Health Plan | n/a | equal | equal | n/a | 327 | 5 | (Commercial) Private Health Insurance | S | Payer | Payer | SOPT |
| 92 | Financial Class | n/a | n/a | FECA Black Lung | n/a | wider | equal | mising program specificity | 323 | 39 | Other Federal Program | S | Payer | Payer | SOPT |
| 93 | Fliuds I/O | INTAKE | INTAKE : Blood | R AN BLOOD ADMINISTRATION VOLUME | n/a | equal | equal | n/a | 4024636 | 116781005 | Blood product volume transfused | S | Observation | Observable Entity | SNOMED |
| 94 | Fliuds I/O | INTAKE | INTAKE : Blood | R PERF AUTOLOGOUS WASHED PRBC | n/a | wider | equal | missing specificity of blood product type, mapped to closest concept representing the clinical concept intent | 4024636 | 116781005 | Blood product volume transfused | S | Observation | Observable Entity | SNOMED |
| 95 | Fliuds I/O | INTAKE | INTAKE : Blood | R PERF AUTOLOGOUS UNWASHED PRBC VOLUME | n/a | wider | equal | missing specificity of blood product type, mapped to closest concept representing the clinical concept intent | 4024636 | 116781005 | Blood product volume transfused | S | Observation | Observable Entity | SNOMED |
| 96 | Fliuds I/O | INTAKE | INTAKE : Blood | R PERF AUTOLOGOUS PLATELET RICH PLASMA (APRP) VOLUME | n/a | wider | equal | missing specificity of blood product type, mapped to closest concept representing the clinical concept intent | 4024636 | 116781005 | Blood product volume transfused | S | Observation | Observable Entity | SNOMED |
| 97 | Fliuds I/O | OUTPUT | OUTPUT : Drains | R SITE ANE OUTPUT NG (DRAIN) | n/a | wider | equal | missing location OR | 4125311 | 235420007 | Intubation of gastrointestinal track via nose | S | Procedure | Procedure | SNOMED |
| 98 | Fliuds I/O | OUTPUT | OUTPUT : Urine Output | R SITE ANE OR URINE | n/a | wider | equal | missing location OR | 3014315 | 9187-6 | Urine output | S | Measurement | Clinical Observation | LOINC |
| 99 | Fliuds I/O | OUTPUT | OUTPUT : Other | (RETIRED SPR19) R PERF RETROGRADE AUTOLOGOUS PRIME (RETIRED) | n/a | wider | equal | missing specificity of blood output type | 4053108 | 16086006 | Blood volume | S | Measurement | Observable Entity | SNOMED |
| 100 | Fliuds I/O | OUTPUT | OUTPUT : Other | R PERF RETROGRADE AUTOLOGOUS PRIME (RAP) | n/a | wider | equal | missing specificity of blood output type | 4053108 | 16086006 | Blood volume | S | Measurement | Observable Entity | SNOMED |
| 101 | Fliuds I/O | OUTPUT | OUTPUT : Blood | R PERF AT CELL SAVER VOLUME PROCESSED | n/a | equal | equal | n/a | 4176156 | 42736004 | Cell saver | S | Observation | Procedure | SNOMED |
| 102 | Fliuds I/O | OUTPUT | OUTPUT : Other | R PERF MODIFIED ULTRAFILTRATE OUTPUT (MUF) | n/a | wider | equal | missing R Perf route | 1988590 | 99741-1 | Ultrafiltrate volume removed | S | Measurement | Clinical Observation | LOINC |
| 103 | Fliuds I/O | OUTPUT | OUTPUT : Other | R PERF ULTRAFILTRATE OUTPUT (CUF) | n/a | wider | equal | missing R Perf route | 1988590 | 99741-1 | Ultrafiltrate volume removed | S | Measurement | Clinical Observation | LOINC |
| 104 | Fliuds I/O | INTAKE | INTAKE : Blood | R SITE ANE OR CELL SAVER | n/a | equal | equal | n/a | 1620887 | LA32164-8 | Cell saver | S | Meas Value | Answer | LOINC |
| 105 | Fliuds I/O | OUTPUT | OUTPUT : Other Output | R SITE ANE OR OTHER OUTPUT | n/a | wider | equal | missing specificity of fluid output type | 4090192 | 251840008 | Fluid output | S | Measurement | Observable Entity | SNOMED |
| 106 | Fliuds I/O | INTAKE | INTAKE : Oral | ORAL | n/a | equal | equal | n/a | 3656010 | 870690008 | Oral intake | S | Observation | Observable Entity | SNOMED |
| 107 | Fliuds I/O | OUTPUT | OUTPUT : Urine | URINE (VOID) | n/a | equal | equal | n/a | 3014315 | 9187-6 | Urine output | S | Measurement | Clinical Observation | LOINC |
| 108 | Fliuds I/O | OUTPUT | OUTPUT : Urine | REPORTED URINE | n/a | equal | equal | n/a | 3014315 | 9187-6 | Urine output | S | Measurement | Clinical Observation | LOINC |
| 109 | Fliuds I/O | OUTPUT | OUTPUT : Estimated Blood Loss | REPORTED/OBSERVED EST. BLOOD LOSS NON C-SECTION | n/a | wider | equal | missing non c-section | 4254486 | 409084000 | Estimated blood loss | S | Observation | Observable Entity | SNOMED |
| 110 | Fliuds I/O | INTAKE | INTAKE : Crystalloid | REPORTED CRYSTALLOID INTAKE | n/a | wider | equal | missing crystalloid specificity | 45884925 | LA11258-3 | Infusion, intravenous | S | Meas Value | Answer | LOINC |
| 111 | Fliuds I/O | INTAKE | INTAKE : Blood | REPORTED BLOOD PRODUCT INTAKE | n/a | equal | equal | n/a | 4024636 | 116781005 | Blood product volume transfused | S | Observation | Observable Entity | SNOMED |
| 112 | Fliuds I/O | OUTPUT | OUTPUT : Other | FLUID LOSS | n/a | equal | equal | n/a | 4090192 | 251840008 | Fluid output | S | Measurement | Observable Entity | SNOMED |
| 113 | Fliuds I/O | INTAKE | INTAKE : Blood | (RETIRED) AMOUNT (ML) BLOOD | n/a | equal | equal | n/a | 4024636 | 116781005 | Blood product volume transfused | S | Observation | Observable Entity | SNOMED |
| 114 | Fliuds I/O | OUTPUT | OUTPUT : Urine | REPORTED VOID | n/a | equal | equal | n/a | 3014315 | 9187-6 | Urine output | S | Measurement | Clinical Observation | LOINC |
| 115 | Fliuds I/O | INTAKE | INTAKE : Colloid | REPORTED COLLOID | n/a | wider | wider | missing colloid specificity | 45884925 | LA11258-3 | Infusion, intravenous | S | Meas Value | Answer | LOINC |
| 116 | Fliuds I/O | INTAKE | INTAKE : Crystalloid | (RETIRED) NS AMOUNT (ML) | n/a | wider | equal | missing crystalloid and NS specificity | 45884925 | LA11258-3 | Infusion, intravenous | S | Meas Value | Answer | LOINC |
| 117 | Fliuds I/O | OUTPUT | OUTPUT : Drains | R DRAIN OUTPUT | n/a | equal | equal | n/a | 4090194 | 251843005 | Fluid output from drain | S | Measurement | Observable Entity | SNOMED |
| 118 | Fliuds I/O | OUTPUT | OUTPUT : Chest tube | R CHEST TUBE OUTPUT | n/a | equal | equal | n/a | 3026556 | 9129-8 | Fluid output chest tube | S | Measurement | Clinical Observation | LOINC |
| 119 | Fliuds I/O | OUTPUT | OUTPUT : Chest tube | R Y CHEST TUBE OUTPUT 1 AND 2 | n/a | equal | equal | n/a | 3026556 | 9129-8 | Fluid output chest tube | S | Measurement | Clinical Observation | LOINC |
| 120 | Fliuds I/O | OUTPUT | OUTPUT : Chest tube | R Y CHEST TUBE OUTPUT 3 AND 4 | n/a | equal | equal | n/a | 3026556 | 9129-8 | Fluid output chest tube | S | Measurement | Clinical Observation | LOINC |
| 121 | Fliuds I/O | OUTPUT | OUTPUT : Drains | SITE R DRAIN OUTPUT (ML) | n/a | wider | equal | missing laterality | 4090194 | 251843005 | Fluid output from drain | S | Measurement | Observable Entity | SNOMED |
| 122 | Fliuds I/O | INTAKE | INTAKE : Blood | AMOUNT (ML) BLOOD NEW | n/a | equal | equal | n/a | 4024636 | 116781005 | Blood product volume transfused | S | Observation | Observable Entity | SNOMED |
| 123 | Fliuds I/O | INTAKE | INTAKE : Crystalloid | NS AMOUNT (ML) NEW | n/a | wider | equal | missing crystalloid and NS specificity | 45884925 | LA11258-3 | Infusion, intravenous | S | Meas Value | Answer | LOINC |
| 124 | Fliuds I/O | OUTPUT | OUTPUT : Urine | SITE R INTERMITTENT CATH URINE OUTPUT (ML) | n/a | wider | equal | missing intermittent cath | 3014315 | 9187-6 | Urine output | S | Measurement | Clinical Observation | LOINC |
| 125 | Fliuds I/O | INTAKE | INTAKE : Crystalloid : Amiodarone | R AMIODARONE VOLUME | n/a | wider | equal | missing medication and crystalloid specificty; mapped to intake | 45884925 | LA11258-3 | Infusion, intravenous | S | Meas Value | Answer | LOINC |
| 126 | Fliuds I/O | INTAKE | INTAKE : Crystalloid : Dexmedetomidine | R DEXMEDETOMIDINE VOLUME | n/a | wider | equal | missing medication and crystalloid specificty; mapped to intake | 45884925 | LA11258-3 | Infusion, intravenous | S | Meas Value | Answer | LOINC |
| 127 | Fliuds I/O | INTAKE | INTAKE : Crystalloid : Diltiazem | R DILTIAZEM VOLUME | n/a | wider | equal | missing medication and crystalloid specificty; mapped to intake | 45884925 | LA11258-3 | Infusion, intravenous | S | Meas Value | Answer | LOINC |
| 128 | Fliuds I/O | INTAKE | INTAKE : Crystalloid : DOBUTamine | R DOBUTAMINE VOLUME | n/a | wider | equal | missing medication and crystalloid specificty; mapped to intake | 45884925 | LA11258-4 | Infusion, intravenous | S | Meas Value | Answer | LOINC |
| 129 | Fliuds I/O | INTAKE | INTAKE : Crystalloid : DOPamine | R DOPAMINE VOLUME | n/a | wider | equal | missing medication and crystalloid specificty; mapped to intake | 45884925 | LA11258-5 | Infusion, intravenous | S | Meas Value | Answer | LOINC |
| 130 | Fliuds I/O | INTAKE | INTAKE : Crystalloid : Epinephrine | R EPINEPHRINE VOLUME | n/a | wider | equal | missing medication and crystalloid specificty; mapped to intake | 45884925 | LA11258-6 | Infusion, intravenous | S | Meas Value | Answer | LOINC |
| 131 | Fliuds I/O | INTAKE | INTAKE : Crystalloid : Fentanyl | R FENTANYL VOLUME | n/a | wider | equal | missing medication and crystalloid specificty; mapped to intake | 45884925 | LA11258-7 | Infusion, intravenous | S | Meas Value | Answer | LOINC |
| 132 | Fliuds I/O | INTAKE | INTAKE : Crystalloid : Insulin | R INSULIN VOLUME | n/a | wider | equal | missing medication and crystalloid specificty; mapped to intake | 45884925 | LA11258-8 | Infusion, intravenous | S | Meas Value | Answer | LOINC |
| 133 | Fliuds I/O | INTAKE | INTAKE : Crystalloid : Lidocaine | R LIDOCAINE VOLUME | n/a | wider | equal | missing medication and crystalloid specificty; mapped to intake | 45884925 | LA11258-9 | Infusion, intravenous | S | Meas Value | Answer | LOINC |
| 134 | Fliuds I/O | INTAKE | INTAKE : Crystalloid : Nitroglycerin | R NITROGLYCERIN VOLUME | n/a | wider | equal | missing medication and crystalloid specificty; mapped to intake | 45884925 | LA11258-10 | Infusion, intravenous | S | Meas Value | Answer | LOINC |
| 135 | Fliuds I/O | INTAKE | INTAKE : Crystalloid : Nitroprusside | R NITROPRUSSIDE VOLUME | n/a | wider | equal | missing medication and crystalloid specificty; mapped to intake | 45884925 | LA11258-11 | Infusion, intravenous | S | Meas Value | Answer | LOINC |
| 136 | Fliuds I/O | INTAKE | INTAKE : Crystalloid : Norepinephrine | R NOREPINEPHRINE VOLUME | n/a | wider | equal | missing medication and crystalloid specificty; mapped to intake | 45884925 | LA11258-12 | Infusion, intravenous | S | Meas Value | Answer | LOINC |
| 137 | Fliuds I/O | INTAKE | INTAKE : Crystalloid : Phenylephrine | R PHENYLEPHRINE VOLUME | n/a | wider | equal | missing medication and crystalloid specificty; mapped to intake | 45884925 | LA11258-13 | Infusion, intravenous | S | Meas Value | Answer | LOINC |
| 138 | Fliuds I/O | INTAKE | INTAKE : Crystalloid : Propofol | R PROPOFOL VOLUME | n/a | wider | equal | missing medication and crystalloid specificty; mapped to intake | 45884925 | LA11258-14 | Infusion, intravenous | S | Meas Value | Answer | LOINC |
| 139 | Fliuds I/O | INTAKE | INTAKE : Crystalloid : Sodium Bicarbonate | R SODIUM BICARBONATE VOLUME | n/a | wider | equal | missing medication and crystalloid specificty; mapped to intake | 45884925 | LA11258-15 | Infusion, intravenous | S | Meas Value | Answer | LOINC |
| 140 | Fliuds I/O | INTAKE | INTAKE : Crystalloid : Vasopressin | R VASOPRESSIN VOLUME | n/a | wider | equal | missing medication and crystalloid specificty; mapped to intake | 45884925 | LA11258-16 | Infusion, intravenous | S | Meas Value | Answer | LOINC |
| 141 | Fliuds I/O | INTAKE | INTAKE : Crystalloid | R CRYSTALLOID IV FLUID AMOUNT | n/a | wider | equal | missing crystalloid specificity | 45884925 | LA11258-3 | Infusion, intravenous | S | Meas Value | Answer | LOINC |
| 142 | Fliuds I/O | INTAKE | INTAKE : Crystalloid : Clevidipine | CLEVIDIPINE VOLUME (ML) | n/a | wider | equal | missing medication and crystalloid specificty; mapped to intake | 45884925 | LA11258-3 | Infusion, intravenous | S | Meas Value | Answer | LOINC |
| 143 | Fliuds I/O | INTAKE | INTAKE : Crystalloid | R AMOUNT CRYSTALLOID INFUSION | n/a | wider | equal | missing crystalloid and NS specificity | 45884925 | LA11258-3 | Infusion, intravenous | S | Meas Value | Answer | LOINC |
| 144 | Fliuds I/O | INTAKE | INTAKE : Colloid | R AMOUNT COLLOID INFUSION | n/a | wider | wider | missing colloid specificity | 45884925 | LA11258-3 | Infusion, intravenous | S | Meas Value | Answer | LOINC |
| 145 | Fliuds I/O | INTAKE | INTAKE : Crystalloid | R PCA 1 MG/ML DILAUDID MG VOLUME INFUSED (ML) CALC | n/a | wider | equal | missing medication and crystalloid specificty; mapped to intake | 45884925 | LA11258-3 | Infusion, intravenous | S | Meas Value | Answer | LOINC |
| 146 | Fliuds I/O | INTAKE | INTAKE : Crystalloid | R AMOUNT IVPB CRYSTALLOID INFUSION | n/a | wider | equal | missing crystalloid specificity | 45884925 | LA11258-3 | Infusion, intravenous | S | Meas Value | Answer | LOINC |

| | | | | | | | | | | | | | | | |
|---|---|---|---|---|---|---|---|---|---|---|---|---|---|---|---|
| 147 | Fliuds I/O | INTAKE | INTAKE : Colloid | R AMOUNT IVPB COLLOID INFUSION | n/a | wider | wider | missing colloid specificity | 45884925 | LA11258-3 | Infusion, intravenous | S | Meas Value | Answer | LOINC |
| 148 | Fliuds I/O | INTAKE | INTAKE : Crystalloid | R IV FLUSH/PUSH AMOUNT (ML) | n/a | wider | equal | missing crystalloid specificity | 45884925 | LA11258-3 | Infusion, intravenous | S | Meas Value | Answer | LOINC |
| 149 | Fliuds I/O | OUTPUT | OUTPUT : Urine Output | URINE OUTPUT | n/a | equal | equal | n/a | 3014315 | 9187-6 | Urine output | S | Measurement | Clinical Observation | LOINC |
| 150 | Fliuds I/O | OUTPUT | OUTPUT : Blood | R OB QBL OR C-SECTION | n/a | wider | equal | missing c-section | 4254486 | 409084000 | Estimated blood loss | S | Observation | Observable Entity | SNOMED |
| 151 | Fliuds I/O | OUTPUT | OUTPUT : Urine | CPM F09 ROW URINE OUTPUT (ML) | n/a | equal | equal | n/a | 3014315 | 9187-6 | Urine output | S | Measurement | Clinical Observation | LOINC |
| 152 | Fliuds I/O | OUTPUT | OUTPUT : Stool | CPM F09 ROW STOOL OUTPUT (ML) | n/a | equal | equal | n/a | 3009704 | 17606-5 | Volume of stool | S | Measurement | Lab Test | LOINC |
| 153 | Fliuds I/O | OUTPUT | OUTPUT : Other | CPM F09 ROW IRRIGATION OUTPUT (ML) | n/a | equal | equal | n/a | 4194653 | 67889009 | Irrigation | S | Procedure | Procedure | SNOMED |
| 154 | Fliuds I/O | OUTPUT | OUTPUT : Other | CPM F09 ROW GENERAL OUTPUT (ML) | n/a | wider | equal | missing specificity of other as fluid output type | 4090192 | 251840008 | Fluid output | S | Measurement | Observable Entity | SNOMED |
| 155 | Fliuds I/O | OUTPUT | OUTPUT : Gastric | CPM ROW GAVAGE IRRIGANT OUTPUT | n/a | wider | equal | missing specificity of gavage as fluid output type | 4194653 | 67889009 | Irrigation | S | Procedure | Procedure | SNOMED |
| 156 | Fliuds I/O | INTAKE | INTAKE : Blood | SITE REPORTED BLOOD CELL SAVER | n/a | equal | equal | n/a | 1620887 | LA32164-8 | Cell saver | S | Meas Value | Answer | LOINC |
| 157 | Fliuds I/O | OUTPUT | OUTPUT : Blood | R QUANTITATIVE BLOOD LOSS | n/a | equal | equal | n/a | 4254486 | 409084000 | Estimated blood loss | S | Observation | Observable Entity | SNOMED |
| 158 | Fliuds I/O | INTAKE | INTAKE : Blood | R PERF AUTOLOGOUS WHOLE BLOOD VOLUME GIVEN | n/a | wider | equal | missing specificity of blood product type, mapped to closest concept representing the clinical concept intent | 4024636 | 116781005 | Blood product volume transfused | S | Observation | Observable Entity | SNOMED |
| 159 | Fliuds I/O | OUTPUT | OUTPUT : Blood | R PERF AUTOLOGOUS WHOLE BLOOD VOLUME REMOVED | n/a | equal | equal | n/a | 4176156 | 42736004 | Blood unit collection for autotransfusion | S | Observation | Procedure | SNOMED |
| 160 | Fliuds I/O | OUTPUT | OUTPUT : Urine | (RETIRED SPR19) R PERF ULTRAFILTRATE OUTPUT (RETIRED) | n/a | wider | equal | missing R Perf route | 1988590 | 99741-1 | Ultrafiltrate volume removed | S | Measurement | Clinical Observation | LOINC |
| 161 | Fliuds I/O | OUTPUT | OUTPUT : Urine | (RETIRED SPR19) R PERF MODIFIED ULTRAFILTRATE OUTPUT (RETIRED) | n/a | wider | equal | missing R Perf route | 1988590 | 99741-1 | Ultrafiltrate volume removed | S | Measurement | Clinical Observation | LOINC |
| 162 | Fliuds I/O | INTAKE | INTAKE : Blood | SITE R MTP REPORTED VOLUME - FFP BAG 1 | n/a | wider | equal | missing specificity of blood product type, mapped to closest concept representing the clinical concept intent | 4024636 | 116781005 | Blood product volume transfused | S | Observation | Observable Entity | SNOMED |
| 163 | Fliuds I/O | INTAKE | INTAKE : Blood | SITE R BPAM REPORTED VOLUME - RBC MTP BAG 1 | n/a | wider | equal | missing specificity of blood product type, mapped to closest concept representing the clinical concept intent | 4024636 | 116781005 | Blood product volume transfused | S | Observation | Observable Entity | SNOMED |
| 164 | Fliuds I/O | INTAKE | INTAKE : Blood | SITE R MTP REPORTED VOLUME - PLATELETPHERESIS | n/a | wider | equal | missing specificity of blood product type, mapped to closest concept representing the clinical concept intent | 4024636 | 116781005 | Blood product volume transfused | S | Observation | Observable Entity | SNOMED |
| 165 | Fliuds I/O | INTAKE | INTAKE : Blood | SITE R BPAM REPORTED VOLUME - CRYOPRECIPITATE | n/a | wider | equal | missing specificity of blood product type, mapped to closest concept representing the clinical concept intent | 4024636 | 116781005 | Blood product volume transfused | S | Observation | Observable Entity | SNOMED |
| 166 | Fliuds I/O | INTAKE | INTAKE : Blood | SITE R BPAM REPORTED VOLUME - CRYOPRECIPITATE POOLED | n/a | wider | equal | missing specificity of blood product type, mapped to closest concept representing the clinical concept intent | 4024636 | 116781005 | Blood product volume transfused | S | Observation | Observable Entity | SNOMED |
| 167 | Fliuds I/O | INTAKE | INTAKE : Blood | SITE R BPAM REPORTED VOLUME - FFP | n/a | wider | equal | missing specificity of blood product type, mapped to closest concept representing the clinical concept intent | 4024636 | 116781005 | Blood product volume transfused | S | Observation | Observable Entity | SNOMED |
| 168 | Fliuds I/O | INTAKE | INTAKE : Blood | SITE R BPAM REPORTED VOLUME - PLATELETPHERESIS | n/a | wider | equal | missing specificity of blood product type, mapped to closest concept representing the clinical concept intent | 4024636 | 116781005 | Blood product volume transfused | S | Observation | Observable Entity | SNOMED |
| 169 | Fliuds I/O | INTAKE | INTAKE : Blood | SITE R BPAM REPORTED VOLUME - RBC | n/a | wider | equal | missing specificity of blood product type, mapped to closest concept representing the clinical concept intent | 4024636 | 116781005 | Blood product volume transfused | S | Observation | Observable Entity | SNOMED |
| 170 | Fliuds I/O | INTAKE | INTAKE : Blood | SITE R BPAM REPORTED VOLUME - RBC MTP BAG 2 | n/a | wider | equal | missing specificity of blood product type, mapped to closest concept representing the clinical concept intent | 4024636 | 116781005 | Blood product volume transfused | S | Observation | Observable Entity | SNOMED |
| 171 | Fliuds I/O | INTAKE | INTAKE : Blood | SITE R BPAM REPORTED VOLUME - RBC MTP BAG 3 | n/a | wider | equal | missing specificity of blood product type, mapped to closest concept representing the clinical concept intent | 4024636 | 116781005 | Blood product volume transfused | S | Observation | Observable Entity | SNOMED |
| 172 | Fliuds I/O | INTAKE | INTAKE : Blood | SITE R BPAM REPORTED VOLUME - RBC MTP BAG 4 | n/a | wider | equal | missing specificity of blood product type, mapped to closest concept representing the clinical concept intent | 4024636 | 116781005 | Blood product volume transfused | S | Observation | Observable Entity | SNOMED |
| 173 | Fliuds I/O | INTAKE | INTAKE : Blood | SITE R BPAM REPORTED VOLUME - RBC MTP BAG 5 | n/a | wider | equal | missing specificity of blood product type, mapped to closest concept representing the clinical concept intent | 4024636 | 116781005 | Blood product volume transfused | S | Observation | Observable Entity | SNOMED |
| 174 | Fliuds I/O | INTAKE | INTAKE : Blood | SITE R MTP REPORTED VOLUME - FFP BAG 2 | n/a | wider | equal | missing specificity of blood product type, mapped to closest concept representing the clinical concept intent | 4024636 | 116781005 | Blood product volume transfused | S | Observation | Observable Entity | SNOMED |
| 175 | Fliuds I/O | INTAKE | INTAKE : Blood | SITE R MTP REPORTED VOLUME - FFP BAG 3 | n/a | wider | equal | missing specificity of blood product type, mapped to closest concept representing the clinical concept intent | 4024636 | 116781005 | Blood product volume transfused | S | Observation | Observable Entity | SNOMED |
| 176 | Fliuds I/O | INTAKE | INTAKE : Blood | SITE R MTP REPORTED VOLUME - FFP BAG 4 | n/a | wider | equal | missing specificity of blood product type, mapped to closest concept representing the clinical concept intent | 4024636 | 116781005 | Blood product volume transfused | S | Observation | Observable Entity | SNOMED |
| 177 | Fliuds I/O | INTAKE | INTAKE : Blood | SITE R MTP REPORTED VOLUME - FFP BAG 5 | n/a | wider | equal | missing specificity of blood product type, mapped to closest concept representing the clinical concept intent | 4024636 | 116781005 | Blood product volume transfused | S | Observation | Observable Entity | SNOMED |
| 178 | Fliuds I/O | OUTPUT | OUTPUT : Stool | STOOL (WEIGHT) | n/a | equal | equal | n/a | 37079480 | LA30063-4 | Stool | S | Meas Value | Answer | LOINC |
| 179 | Fliuds I/O | OUTPUT | OUTPUT : Emesis | EMESIS | n/a | equal | equal | n/a | 4113561 | 300368002 | Vomit volume | S | Measurement | Observable Entity | SNOMED |
| 180 | Fliuds I/O | OUTPUT | OUTPUT : Emesis | REPORTED EMESIS | n/a | equal | equal | n/a | 4113561 | 300368002 | Vomit volume | S | Measurement | Observable Entity | SNOMED |
| 181 | Fliuds I/O | INTAKE | INTAKE : Oral | REPORTED ORAL INTAKE | n/a | equal | equal | n/a | 3656010 | 870690008 | Oral intake | S | Observation | Observable Entity | SNOMED |
| 182 | Fliuds I/O | OUTPUT | OUTPUT : Stool | RECTAL TUBE AMOUNT | n/a | equal | equal | n/a | 37079480 | LA30063-4 | Stool | S | Meas Value | Answer | LOINC |
| 183 | Fliuds I/O | OUTPUT | OUTPUT : Other | SITE R UF REMOVED | n/a | wider | equal | missing R Perf route | 1988590 | 99741-1 | Ultrafiltrate volume removed | S | Measurement | Clinical Observation | LOINC |
| 184 | Fliuds I/O | OUTPUT | OUTPUT: Urine and Stool | URINE AND STOOL WT | n/a | wider | equal | missing stool component | 3014315 | 9187-6 | Urine output | S | Measurement | Clinical Observation | LOINC |
| 185 | Fliuds I/O | OUTPUT | OUTPUT : Dialysis | R DIALYSIS FLUID BALANCE (HEMODIALYSIS) | n/a | wider | equal | missing dialysis source | 4090192 | 251840008 | Fluid output | S | Measurement | Observable Entity | SNOMED |
| 186 | Fliuds I/O | INTAKE | INTAKE : Crystalloid | R TIROFIBAN VOLUME | n/a | wider | equal | missing medication and crystalloid specificty; mapped to intake | 45884925 | LA11258-3 | Infusion, intravenous | S | Meas Value | Answer | LOINC |
| 187 | Fliuds I/O | OUTPUT | OUTPUT: Cerebrospinal Fluid | R CEREBROSPINAL FLUID OUTPUT | n/a | wider | equal | missing type of output | 4090192 | 251840008 | Fluid output | S | Measurement | Observable Entity | SNOMED |
| 188 | Fliuds I/O | INTAKE | INTAKE : Intravenous Intake | RETIRED CUMULATIVE INTAKE MORPHINE 1 MG/ML PCA | n/a | wider | equal | missing medication name, but is mapped here to I & O | 45884925 | LA11258-3 | Infusion, intravenous | S | Meas Value | Answer | LOINC |
| 189 | Fliuds I/O | INTAKE | INTAKE : Intravenous Intake | R SITE IP CUMULATIVE INTAKE HYDROMORPHONE 1 MG/ML PCA | n/a | wider | equal | missing medication name, but is mapped here to I & O | 45884925 | LA11258-3 | Infusion, intravenous | S | Meas Value | Answer | LOINC |
| 190 | Fliuds I/O | INTAKE | INTAKE : Intravenous Intake | R SITE IP CUMULATIVE INTAKE FENTANYL 50 MCG/ML PCA | n/a | wider | equal | missing medication name, but is mapped here to I & O | 45884925 | LA11258-3 | Infusion, intravenous | S | Meas Value | Answer | LOINC |
| 191 | Fliuds I/O | INTAKE | INTAKE : Intravenous Intake | R SITE IP CUMULATIVE INTAKE PED NON-WT BASED MORPHINE 1 MG/ML PCA | n/a | wider | equal | missing medication name, but is mapped here to I & O | 45884925 | LA11258-3 | Infusion, intravenous | S | Meas Value | Answer | LOINC |
| 192 | Fliuds I/O | OUTPUT | OUTPUT : Urine | SITE R CBI NET OUTPUT | n/a | wider | equal | missing type of output | 4090192 | 251840008 | Fluid output | S | Measurement | Observable Entity | SNOMED |
| 193 | Fliuds I/O | OUTPUT | OUTPUT : Blood | SITE R OB GENERAL OUTPUT (ML) | n/a | wider | equal | missing type of output | 4090192 | 251840008 | Fluid output | S | Measurement | Observable Entity | SNOMED |
| 194 | Fliuds I/O | INTAKE | INTAKE : Irrigant | SITE R COLOSTOMY IRRIGANT INTAKE | n/a | wider | equal | missing colostomy as source | 37164797 | 1234789006 | Measurement of irrigating solution | S | Measurement | Procedure | SNOMED |
| 195 | Fliuds I/O | INTAKE | INTAKE : Crystalloid | R MEDICATION IV FLUID AMOUNT NEW | n/a | wider | equal | missing medication and crystalloid specificty; mapped to intake | 45884925 | LA11258-3 | Infusion, intravenous | S | Meas Value | Answer | LOINC |
| 196 | Fliuds I/O | INTAKE | INTAKE : Crystalloid | R AMINOCAPROIC ACID VOLUME | n/a | wider | equal | missing medication and crystalloid specificty; mapped to intake | 45884925 | LA11258-3 | Infusion, intravenous | S | Meas Value | Answer | LOINC |
| 197 | Fliuds I/O | INTAKE | INTAKE : Crystalloid | R CISATRACURIUM VOLUME | n/a | wider | equal | missing medication and crystalloid specificty; mapped to intake | 45884925 | LA11258-3 | Infusion, intravenous | S | Meas Value | Answer | LOINC |
| 198 | Fliuds I/O | INTAKE | INTAKE : Crystalloid | R ESMOLOL VOLUME | n/a | wider | equal | missing medication and crystalloid specificty; mapped to intake | 45884925 | LA11258-3 | Infusion, intravenous | S | Meas Value | Answer | LOINC |
| 199 | Fliuds I/O | INTAKE | INTAKE : Crystalloid | R FUROSEMIDE VOLUME | n/a | wider | equal | missing medication and crystalloid specificty; mapped to intake | 45884925 | LA11258-3 | Infusion, intravenous | S | Meas Value | Answer | LOINC |
| 200 | Fliuds I/O | INTAKE | INTAKE : Crystalloid | R HEPARIN VOLUME | n/a | wider | equal | missing medication and crystalloid specificty; mapped to intake | 45884925 | LA11258-3 | Infusion, intravenous | S | Meas Value | Answer | LOINC |
| 201 | Fliuds I/O | INTAKE | INTAKE : Crystalloid | R HYDROMORPHONE VOLUME | n/a | wider | equal | missing medication and crystalloid specificty; mapped to intake | 45884925 | LA11258-3 | Infusion, intravenous | S | Meas Value | Answer | LOINC |

| | | | | | | | | | | | | | | | |
|---|---|---|---|---|---|---|---|---|---|---|---|---|---|---|---|
| 202 | Fliuds I/O | INTAKE | INTAKE : Crystalloid | R ISOPROTERENOL VOLUME | n/a | wider | equal | missing medication and crystalloid specificty; mapped to intake | 45884925 | LA11258-3 | Infusion, intravenous | S | Meas Value | Answer | LOINC |
| 203 | Fliuds I/O | INTAKE | INTAKE : Crystalloid | R LABETALOL VOLUME | n/a | wider | equal | missing medication and crystalloid specificty; mapped to intake | 45884925 | LA11258-3 | Infusion, intravenous | S | Meas Value | Answer | LOINC |
| 204 | Fliuds I/O | INTAKE | INTAKE : Crystalloid | R MAGNESIUM SULFATE IN LR VOLUME | n/a | wider | equal | missing medication and crystalloid specificty; mapped to intake | 45884925 | LA11258-3 | Infusion, intravenous | S | Meas Value | Answer | LOINC |
| 205 | Fliuds I/O | INTAKE | INTAKE : Crystalloid | R METHYLPREDNISOLONE VOLUME | n/a | wider | equal | missing medication and crystalloid specificty; mapped to intake | 45884925 | LA11258-3 | Infusion, intravenous | S | Meas Value | Answer | LOINC |
| 206 | Fliuds I/O | INTAKE | INTAKE : Crystalloid | R MIDAZOLAM VOLUME | n/a | wider | equal | missing medication and crystalloid specificty; mapped to intake | 45884925 | LA11258-3 | Infusion, intravenous | S | Meas Value | Answer | LOINC |
| 207 | Fliuds I/O | INTAKE | INTAKE : Crystalloid | R MILRINONE VOLUME | n/a | wider | equal | missing medication and crystalloid specificty; mapped to intake | 45884925 | LA11258-3 | Infusion, intravenous | S | Meas Value | Answer | LOINC |
| 208 | Fliuds I/O | INTAKE | INTAKE : Crystalloid | R MORPHINE VOLUME | n/a | wider | equal | missing medication and crystalloid specificty; mapped to intake | 45884925 | LA11258-3 | Infusion, intravenous | S | Meas Value | Answer | LOINC |
| 209 | Fliuds I/O | INTAKE | INTAKE : Crystalloid | R NAFCILLIN VOLUME | n/a | wider | equal | missing medication and crystalloid specificty; mapped to intake | 45884925 | LA11258-3 | Infusion, intravenous | S | Meas Value | Answer | LOINC |
| 210 | Fliuds I/O | INTAKE | INTAKE : Crystalloid | R NALOXONE VOLUME | n/a | wider | equal | missing medication and crystalloid specificty; mapped to intake | 45884925 | LA11258-3 | Infusion, intravenous | S | Meas Value | Answer | LOINC |
| 211 | Fliuds I/O | INTAKE | INTAKE : Crystalloid | R NICARDIPINE VOLUME | n/a | wider | equal | missing medication and crystalloid specificty; mapped to intake | 45884925 | LA11258-3 | Infusion, intravenous | S | Meas Value | Answer | LOINC |
| 212 | Fliuds I/O | INTAKE | INTAKE : Crystalloid | R OCTREOTIDE VOLUME | n/a | wider | equal | missing medication and crystalloid specificty; mapped to intake | 45884925 | LA11258-3 | Infusion, intravenous | S | Meas Value | Answer | LOINC |
| 213 | Fliuds I/O | INTAKE | INTAKE : Crystalloid | R OXYTOCIN VOLUME | n/a | wider | equal | missing medication and crystalloid specificty; mapped to intake | 45884925 | LA11258-3 | Infusion, intravenous | S | Meas Value | Answer | LOINC |
| 214 | Fliuds I/O | INTAKE | INTAKE : Crystalloid | (RETIRED) R MEDICATION IV FLUID AMOUNT | n/a | wider | equal | missing medication and crystalloid specificty; mapped to intake | 45884925 | LA11258-3 | Infusion, intravenous | S | Meas Value | Answer | LOINC |
| 215 | Fliuds I/O | INTAKE | INTAKE : Crystalloid | R CALCIUM GLUCONATE VOLUME | n/a | wider | equal | missing medication and crystalloid specificty; mapped to intake | 45884925 | LA11258-3 | Infusion, intravenous | S | Meas Value | Answer | LOINC |
| 216 | Fliuds I/O | INTAKE | INTAKE : Crystalloid | R PANTOPRAZOLE VOLUME | n/a | wider | equal | missing medication and crystalloid specificty; mapped to intake | 45884925 | LA11258-3 | Infusion, intravenous | S | Meas Value | Answer | LOINC |
| 217 | Fliuds I/O | INTAKE | INTAKE : Crystalloid | R IMPELLA HEPARIN VOLUME | n/a | wider | equal | missing medication name and device; mapped here to intake | 45884925 | LA11258-3 | Infusion, intravenous | S | Meas Value | Answer | LOINC |
| 218 | Fliuds I/O | INTAKE | INTAKE : Colloid | R COLLOID IV FLUID AMOUNT | n/a | wider | wider | missing colloid specificity | 45884925 | LA11258-3 | Infusion, intravenous | S | Meas Value | Answer | LOINC |
| 219 | Fliuds I/O | INTAKE | INTAKE : Parenteral Nutrition | R TPN IV FLUID AMOUNT (MANUALLY ADD-REVISED) | n/a | wider | equal | missing TPN as type of intake | 45884925 | LA11258-3 | Infusion, intravenous | S | Meas Value | Answer | LOINC |
| 220 | Fliuds I/O | INTAKE | INTAKE : Other | R EPIDURAL TOTAL VOL (ML) | n/a | wider | equal | missing route | 3037106 | 9103-3 | Fluid intake total measured | S | Measurement | Clinical Observation | LOINC |
| 221 | Fliuds I/O | INTAKE | INTAKE : Intravenous Intake | R SITE IP CUMULATIVE INTAKE MORPHINE 1MG/ML PCA NEW | n/a | wider | equal | missing PCA as type of intake and missing medication name, but mapped to I&O | 45884925 | LA11258-3 | Infusion, intravenous | S | Meas Value | Answer | LOINC |
| 222 | Fliuds I/O | INTAKE | INTAKE : Urinary Irrigant | R SITE URINARY IRRIGANT INTAKE | n/a | equal | equal | n/a | 36308083 | LA9421-4 | GU irrigant | S | Meas Value | Answer | LOINC |
| 223 | Fliuds I/O | OUTPUT | OUTPUT : Urinary Irrigant | R SITE IP URINARY IRRGANT OUTPUT | n/a | equal | equal | n/a | 45884906 | LA11424-1 | Irrigation, urinary bladder | S | Measurement | Procedure | LOINC |
| 224 | Fliuds I/O | INTAKE | INTAKE : Epidural | R EPIDURAL BOLUS DOSE BY MD (EPIDURAL TYPE) (ML) | n/a | wider | equal | missing route | 3037106 | 9103-3 | Fluid intake total measured | S | Measurement | Clinical Observation | LOINC |
| 225 | Fliuds I/O | INTAKE | INTAKE : Epidural | R EPIDURAL PRIME TUBING AMOUNT (EPIDURAL TYPE) (ML) VIA PUMP | n/a | wider | equal | missing route | 3037106 | 9103-3 | Fluid intake total measured | S | Measurement | Clinical Observation | LOINC |
| 226 | Fliuds I/O | INTAKE | INTAKE : Epidural | EPIDURAL INFUSED (ML) (EPIDURAL TYPE) * | n/a | wider | equal | missing route | 3037106 | 9103-3 | Fluid intake total measured | S | Measurement | Clinical Observation | LOINC |
| 227 | Fliuds I/O | INTAKE | INTAKE : Crystalloid | R IMPELLA SODIUM BICARBONATE VOLUME | n/a | wider | equal | missing medication name and device; mapped here to intake | 45884925 | LA11258-3 | Infusion, intravenous | S | Meas Value | Answer | LOINC |
| 228 | Fliuds I/O | INTAKE | INTAKE : Irrigant | R RECTAL IRRIGANT / FLUSH AMOUNT (ML) DUPLICABLE | n/a | wider | equal | missing route | 37164797 | 1234789006 | Measurement of irrigating solution | S | Measurement | Procedure | SNOMED |
| 229 | Fliuds I/O | INTAKE | INTAKE : Other | R PCEA VOLUME INFUSED ML CALC | n/a | wider | equal | missing route | 3037106 | 9103-3 | Fluid intake total measured | S | Measurement | Clinical Observation | LOINC |
| 230 | Fliuds I/O | INTAKE | INTAKE : Other | (RETIRED) R AMOUNT ML (NERVE BLOCK) | n/a | wider | equal | missing route | 3037106 | 9103-3 | Fluid intake total measured | S | Measurement | Clinical Observation | LOINC |
| 231 | Fliuds I/O | INTAKE | INTAKE : Nerve Block | R AMOUNT ML (NERVE BLOCK) (NERVE BLOCK TYPE) | n/a | wider | equal | missing route | 3037106 | 9103-3 | Fluid intake total measured | S | Measurement | Clinical Observation | LOINC |
| 232 | Fliuds I/O | INTAKE | INTAKE : Other | (RETIRED) CLINICIAN BOLUS MD (ML) VIA PUMP | n/a | wider | equal | missing route | 45884925 | LA11258-3 | Infusion, intravenous | S | Meas Value | Answer | LOINC |
| 233 | Fliuds I/O | INTAKE | INTAKE : Nerve Block | R CLINICIAN BOLUS VOL (ML) NERVE BLOCK (NERVE BLOCK TYPE) | n/a | wider | equal | missing route | 3037106 | 9103-3 | Fluid intake total measured | S | Measurement | Clinical Observation | LOINC |
| 234 | Fliuds I/O | INTAKE | INTAKE : Other | R EPIDURAL BOLUS DOSE BY MD (ML) | n/a | wider | equal | missing route | 3037106 | 9103-3 | Fluid intake total measured | S | Measurement | Clinical Observation | LOINC |
| 235 | Fliuds I/O | INTAKE | INTAKE : Other | (RETIRED) EPIDURAL INFUSED (ML) * | n/a | wider | equal | missing route | 3037106 | 9103-3 | Fluid intake total measured | S | Measurement | Clinical Observation | LOINC |
| 236 | Fliuds I/O | INTAKE | INTAKE : Crystalloid | R VANCOMYCIN VOLUME | n/a | wider | equal | missing medication name; mapped to I&O | 45884925 | LA11258-3 | Infusion, intravenous | S | Meas Value | Answer | LOINC |
| 237 | Fliuds I/O | INTAKE | INTAKE : Crystalloid | R PCA 10 MCG/ML VOLUME INFUSED (ML) CALC | n/a | wider | equal | missing medication name and PCA route; mapped here to intake | 45884925 | LA11258-3 | Infusion, intravenous | S | Meas Value | Answer | LOINC |
| 238 | Fliuds I/O | INTAKE | INTAKE : Crystalloid | R PCA MS 1 MG/ML VOLUME INFUSED (ML) CALC | n/a | wider | equal | missing medication name and PCA route; mapped here to intake | 45884925 | LA11258-3 | Infusion, intravenous | S | Meas Value | Answer | LOINC |
| 239 | Fliuds I/O | INTAKE | INTAKE : Crystalloid | R PCA 50 MCG/ML VOLUME INFUSED (ML) CALC | n/a | wider | equal | missing medication name and PCA route; mapped here to intake | 45884925 | LA11258-3 | Infusion, intravenous | S | Meas Value | Answer | LOINC |
| 240 | Fliuds I/O | INTAKE | INTAKE : Crystalloid | R PCA 0.2 MG/ML DILAUDID VOLUME INFUSED (ML) CALC | n/a | wider | equal | missing medication name and PCA route; mapped here to intake | 45884925 | LA11258-3 | Infusion, intravenous | S | Meas Value | Answer | LOINC |
| 241 | Fliuds I/O | INTAKE | INTAKE : Crystalloid | R PCA MS 5 MG/ML VOLUME INFUSED (ML) CALC | n/a | wider | equal | missing medication name and PCA route; mapped here to intake | 45884925 | LA11258-3 | Infusion, intravenous | S | Meas Value | Answer | LOINC |
| 242 | Fliuds I/O | INTAKE | INTAKE : Crystalloid | R PCA 5 MG/ML DILAUDID VOLUME INFUSED (ML) CALC | n/a | wider | equal | missing medication name and PCA route; mapped here to intake | 45884925 | LA11258-3 | Infusion, intravenous | S | Meas Value | Answer | LOINC |
| 243 | Fliuds I/O | INTAKE | INTAKE : Crystalloid | R KETAMINE VOLUME | n/a | wider | equal | missing medication name; mapped here to intake | 45884925 | LA11258-3 | Infusion, intravenous | S | Meas Value | Answer | LOINC |
| 244 | Fliuds I/O | INTAKE | INTAKE : Crystalloid | R IV FLUSH/PUSH AMOUNT (ML) | n/a | wider | equal | missing fluid type | 45884925 | LA11258-3 | Infusion, intravenous | S | Meas Value | Answer | LOINC |
| 245 | Fliuds I/O | INTAKE | INTAKE : Lipid | R AMOUNT LIPID SHORT TERM INFUSION | n/a | wider | equal | missing fluid type | 45884925 | LA11258-3 | Infusion, intravenous | S | Meas Value | Answer | LOINC |
| 246 | Fliuds I/O | INTAKE | INTAKE : Crystalloid | R ROCURONIUM VOLUME | n/a | wider | equal | missing medication name; mapped here to intake | 45884925 | LA11258-3 | Infusion, intravenous | S | Meas Value | Answer | LOINC |
| 247 | Fliuds I/O | INTAKE | INTAKE : Parenteral Nutrition | R TPN VOLUME | n/a | wider | equal | missing fluid type | 45884925 | LA11258-3 | Infusion, intravenous | S | Meas Value | Answer | LOINC |
| 248 | Fliuds I/O | INTAKE | INTAKE : Lipid | R FAT VOLUME | n/a | wider | equal | missing fluid type | 45884925 | LA11258-3 | Infusion, intravenous | S | Meas Value | Answer | LOINC |
| 249 | Fliuds I/O | INTAKE | INTAKE : Crystalloid | R SITE IP PCA FENTANYL 50 MCG/ML TOTAL VOLUME INFUSED (ML) | n/a | wider | equal | missing medication name and PCA route; mapped here to intake | 45884925 | LA11258-3 | Infusion, intravenous | S | Meas Value | Answer | LOINC |
| 250 | Fliuds I/O | INTAKE | INTAKE : Crystalloid | R SITE IP PCA HYDROMORPHONE 1 MG/ML TOTAL VOLUME INFUSED (ML) | n/a | wider | equal | missing medication name and PCA route; mapped here to intake | 45884925 | LA11258-3 | Infusion, intravenous | S | Meas Value | Answer | LOINC |
| 251 | Fliuds I/O | INTAKE | INTAKE : Crystalloid | R SITE IP PCA MORPHINE 5 MG/ML TOTAL VOLUME INFUSED (ML) | n/a | wider | equal | missing medication name and PCA route; mapped here to intake | 45884925 | LA11258-3 | Infusion, intravenous | S | Meas Value | Answer | LOINC |
| 252 | Fliuds I/O | INTAKE | INTAKE : Crystalloid | R SITE IP PCA HYDROMORPHONE 5 MG/ML TOTAL VOLUME INFUSED (ML) | n/a | wider | equal | missing medication name and PCA route; mapped here to intake | 45884925 | LA11258-3 | Infusion, intravenous | S | Meas Value | Answer | LOINC |
| 253 | Fliuds I/O | INTAKE | INTAKE: Oral Intake | ORAL INTAKE | n/a | equal | equal | n/a | 3656010 | 870690008 | Oral intake | S | Observation | Observable Entity | SNOMED |
| 254 | Fliuds I/O | INTAKE | INTAKE : Intravenous Intake | INTRAVENOUS INTAKE | n/a | equal | equal | n/a | 45884925 | LA11258-3 | Infusion, intravenous | S | Meas Value | Answer | LOINC |
| 255 | Fliuds I/O | OUTPUT | OUTPUT: Emeisis | EMESIS | n/a | equal | equal | n/a | 4113561 | 300368002 | Vomit volume | S | Measurement | Observable Entity | SNOMED |
| 256 | Fliuds I/O | OUTPUT | OUTPUT : Other Drains | DRAINS | n/a | equal | equal | n/a | 4182282 | 366368001 | Fluid output from drain | S | Observation | Clinical Finding | SNOMED |
| 257 | Fliuds I/O | INTAKE | INTAKE : Enteral | R SITE NASO ORAL TUBE ENTERAL INTAKE (ML) FORMULA | n/a | wider | equal | missing specificity of enteral as tube intake type | 3656010 | 870690008 | Oral intake | S | Observation | Observable Entity | SNOMED |
| 258 | Fliuds I/O | INTAKE | INTAKE : Enteral | R SITE ENT TUBE ENTERAL INTAKE (ML) FORMULA | n/a | wider | equal | missing specificity of enteral as tube intake type | 3656010 | 870690008 | Oral intake | S | Observation | Observable Entity | SNOMED |
| 259 | Fliuds I/O | OUTPUT | OUTPUT : Blood | R SITE OB QUANTIFIED BLOOD LOSS | n/a | equal | equal | n/a | 4254486 | 409084000 | Estimated blood loss | S | Observation | Observable Entity | SNOMED |
| 260 | Fliuds I/O | INTAKE | INTAKE : Crystalloid | R SITE IP CIRCUIT FLUSH (ML) | n/a | wider | equal | missing fluid type or IP circuit as route | 45884925 | LA11258-3 | Infusion, intravenous | S | Meas Value | Answer | LOINC |
| 261 | Fliuds I/O | INTAKE | INTAKE : Enteral | CPM F09 ROW TUBE FEEDING INTAKE (ML) | n/a | wider | equal | missing specificty of tube intake type and feeding purpose | 3656010 | 870690008 | Oral intake | S | Observation | Observable Entity | SNOMED |
| 262 | Fliuds I/O | INTAKE | INTAKE : Crystalloid | R CANGRELOR VOLUME | n/a | wider | equal | missing medication name; mapped here to intake | 45884925 | LA11258-3 | Infusion, intravenous | S | Meas Value | Answer | LOINC |
| 263 | Fliuds I/O | INTAKE | INTAKE : Irrigant | CPM ROW DRAIN IRRIGANT | n/a | wider | equal | missing drain as source | 37164797 | 1234789006 | Measurement of irrigating solution | S | Measurement | Procedure | SNOMED |

| | | | | | | | | | | | | | | | |
|---|---|---|---|---|---|---|---|---|---|---|---|---|---|---|---|
| 264 | Fliuds I/O | INTAKE | INTAKE : Other | CPM DIALYSATE FLUID INTAKE | n/a | wider | equal | missing fluid type | 3037106 | 9103-3 | Fluid intake total measured | S | Measurement | Clinical Observation | LOINC |
| 265 | Fliuds I/O | OUTPUT | OUTPUT : Dialysis | CPM DIALYSATE FLUID OUTPUT | n/a | equal | equal | n/a | 3023665 | 12457-8 | Volume of dialysate fluid | S | Measurement | Lab Test | LOINC |
| 266 | Fliuds I/O | INTAKE | INTAKE : Irrigant | (RETIRED) CPM ROW URINARY IRRIGANT INTAKE | n/a | equal | equal | n/a | 3035875 | 9025-8 | Fluid intake urinary bladder irrigation estimated | S | Measurement | Clinical Observation | LOINC |
| 267 | Fliuds I/O | INTAKE | INTAKE : Irrigant | CPM ROW URETERAL CATHETER INTAKE | n/a | equal | equal | n/a | 3035875 | 9025-8 | Fluid intake urinary bladder irrigation estimated | S | Measurement | Clinical Observation | LOINC |
| 268 | Fliuds I/O | INTAKE | INTAKE : Irrigant | CPM ROW NEPHROSTOMY IRRIGANT INTAKE | n/a | wider | equal | missing specificty of nephrostomy | 36308083 | LA9421-4 | GU irrigant | S | Meas Value | Answer | LOINC |
| 269 | Fliuds I/O | INTAKE | INTAKE : Irrigant | CPM ROW UROSTOMY IRRIGANT INTAKE | n/a | wider | equal | missing specificty of urostomy | 36308083 | LA9421-4 | GU irrigant | S | Meas Value | Answer | LOINC |
| 270 | Fliuds I/O | INTAKE | INTAKE : Irrigant | CPM ROW SUPRAPUBIC CATHETER IRRIGANT INTAKE | n/a | wider | equal | missing specificity of suprapubic catheter as intake source | 36308083 | LA9421-4 | GU irrigant | S | Meas Value | Answer | LOINC |
| 271 | Fliuds I/O | INTAKE | INTAKE : Irrigant | CPM ROW WOUND IRRIGANT INTAKE | n/a | wider | equal | missing specificity of wound | 37164797 | 1234789006 | Measurement of irrigating solution | S | Measurement | Procedure | SNOMED |
| 272 | Fliuds I/O | INTAKE | INTAKE : Other | CPM S11 ROW GENERAL INTAKE (ML) (ADULT, NICU, NEWBORN, OB, PEDIATRIC) | n/a | equal | equal | n/a | 3037106 | 9103-3 | Fluid intake total measured | S | Measurement | Clinical Observation | LOINC |
| 273 | Fliuds I/O | INTAKE | INTAKE : Other | R SITE IP APHERESIS POSITIVE BALANCE | n/a | wider | equal | missing specificity of apheresis as source | 45884925 | LA11258-3 | Infusion, intravenous | S | Meas Value | Answer | LOINC |
| 274 | Fliuds I/O | OUTPUT | OUTPUT : Other | R SITE IP APHERESIS NEGATIVE BALANCE | n/a | wider | equal | missing specificity of apheresis as source | 4090192 | 251840008 | Fluid output | S | Measurement | Observable Entity | SNOMED |
| 275 | Fliuds I/O | INTAKE | INTAKE : Irrigant | R SITE IP GI TUBE IRRIGANT (ML) -FLOWS TO I/O | n/a | wider | equal | missing specificity of GI tube as source | 37164797 | 1234789006 | Measurement of irrigating solution | S | Measurement | Procedure | SNOMED |
| 276 | Fliuds I/O | INTAKE | INTAKE : Crystalloid | R MAGNESIUM SULFATE IN SW VOLUME | n/a | wider | equal | missing medication name; mapped here to intake | 45884925 | LA11258-3 | Infusion, intravenous | S | Meas Value | Answer | LOINC |
| 277 | Fliuds I/O | INTAKE | INTAKE : Crystalloid | SITE R ANGIOTENSIN II VOLUME | n/a | wider | equal | missing medication name; mapped here to intake | 45884925 | LA11258-3 | Infusion, intravenous | S | Meas Value | Answer | LOINC |
| 278 | Flowsheet | Flowsheet Vitals | n/a | R SITE ANE EKG HEART RATE | n/a | equal | equal | n/a | 3027018 | 8867-4 | Heart rate | S | Measurement | Clinical Observation | LOINC |
| 279 | Flowsheet | Flowsheet Vitals | n/a | R SITE ANE SET FRACTIONAL INSPIRED OXYGEN | n/a | equal | equal | n/a | 4353936 | 250774007 | Inspired oxygen concentration | S | Measurement | Observable Entity | SNOMED |
| 280 | Flowsheet | Flowsheet Vitals | n/a | R SITE ANE SET TIDAL VOLUME | n/a | wider | equal | missing specificty of set tidal volume | 4029625 | 13621006 | Tidal volume | S | Measurement | Observable Entity | SNOMED |
| 281 | Flowsheet | Flowsheet Vitals | n/a | R SITE ANE END TIDAL CO2 | n/a | equal | equal | n/a | 4353938 | 250784008 | End tidal carbon dioxide concentration | S | Measurement | Observable Entity | SNOMED |
| 282 | Flowsheet | measurement | n/a | R CRD BMI | n/a | equal | equal | n/a | 3038553 | 39156-5 | Body mass index (BMI) [Ratio] | S | Measurement | Clinical Observation | LOINC |
| 283 | Flowsheet | measurement | n/a | BMI | n/a | equal | equal | n/a | 3038553 | 39156-5 | Body mass index (BMI) [Ratio] | S | Measurement | Clinical Observation | LOINC |
| 284 | Flowsheet | measurement | n/a | SITE IP G STROKE NAV BMI | n/a | equal | equal | n/a | 3038553 | 39156-5 | Body mass index (BMI) [Ratio] | S | Measurement | Clinical Observation | LOINC |
| 285 | Flowsheet | measurement | n/a | R RX TPN PHARM BMI | n/a | equal | equal | n/a | 3038553 | 39156-5 | Body mass index (BMI) [Ratio] | S | Measurement | Clinical Observation | LOINC |
| 286 | Flowsheet | measurement | n/a | PGHD PATIENT ENTERED - BMI | n/a | equal | equal | n/a | 3038553 | 39156-5 | Body mass index (BMI) [Ratio] | S | Measurement | Clinical Observation | LOINC |
| 287 | Flowsheet | measurement | n/a | CPM S11 ROW AS BMI (KG/M2).BODY MEASUREMENTS (ADULT, OB, PEDIATRIC) | n/a | equal | equal | n/a | 3038553 | 39156-5 | Body mass index (BMI) [Ratio] | S | Measurement | Clinical Observation | LOINC |
| 288 | Flowsheet | measurement | n/a | CPM S11 GRP RD BMI (ADULT) | n/a | equal | equal | n/a | 3038553 | 39156-5 | Body mass index (BMI) [Ratio] | S | Measurement | Clinical Observation | LOINC |
| 289 | Flowsheet | measurement | n/a | CPM F14 ROW SGRP AS BODY MASS INDEX (BMI) (ADULT) | n/a | equal | equal | n/a | 3038553 | 39156-5 | Body mass index (BMI) [Ratio] | S | Measurement | Clinical Observation | LOINC |
| 290 | Flowsheet | measurement | n/a | CPM F14 ROW AS BMI (KG/M2) | n/a | equal | equal | n/a | 3038553 | 39156-5 | Body mass index (BMI) [Ratio] | S | Measurement | Clinical Observation | LOINC |
| 291 | Medication | n/a | n/a | EPINEPHRINE DRIP - ADULT 64 MCG/ML | n/a | wider | equal | missing drug concentration, adult, and infusion | 1343916 | 3992 | epinephrine | S | Drug | Ingredient | RxNorm |
| 292 | Medication | n/a | n/a | MILRINONE BOLUS (200 MCG/ML) | n/a | wider | equal | missing drug concentration | 1368671 | 51769 | milrinone | S | Drug | Ingredient | RxNorm |
| 293 | Medication | n/a | n/a | DOPAMINE DRIP - PEDS *HIGH* CONCENTRATION | n/a | equal | equal | n/a | 42799685 | 1292887 | 250 ML dopamine hydrochloride 3.2 MG/ML Injection | S | Drug | Quant Clinical Drug | RxNorm |
| 294 | Medication | n/a | n/a | NOREPINEPHRINE BITARTRATE 32 MCG/ML DRIP WEIGHT-BASED | n/a | equal | equal | n/a | 40167236 | 866824 | Norepinephrine 0.032 MG/ML Injectable Solution | S | Drug | Clinical Drug | RxNorm |
| 295 | Medication | n/a | n/a | NOREPINEPHRINE 64 MCG/ML IV DRIP WEIGHT-BASED | n/a | wider | equal | missing drug concentration and infusion | 1321341 | 7512 | norepinephrine | S | Drug | Ingredient | RxNorm |
| 296 | Medication | n/a | n/a | PHENYLEPHRINE DRIP | n/a | equal | equal | n/a | 42707286 | 1232651, 1666372 | 1 ML phenylephrine hydrochloride 10 MG/ML Injection, phenylephrine hydrochloride 10 MG/ML Injectable Solution | S | Drug | Clinical Drug | RxNorm |
| 297 | Medication | n/a | n/a | VASOPRESSIN DRIP | n/a | narrower | equal | adds concentration | 1507838 | 313578 | vasopressin (USP) 20 UNT/ML Injectable Solution | S | Drug | Clinical Drug | RxNorm |
| 298 | Medication | n/a | n/a | PHENYLEPHRINE DRIP - ADULT STANDARD CONCENTRATION | n/a | equal | equal | n/a | 42707286 | 1232651, 1666372 | 1 ML phenylephrine hydrochloride 10 MG/ML Injection, phenylephrine hydrochloride 10 MG/ML Injectable Solution | S | Drug | Clinical Drug | RxNorm |
| 299 | Medication | n/a | n/a | PHENYLEPHRINE DRIP - 80 MCG/ML STRAIGHT DOSE | n/a | equal | equal | n/a | 46287545 | 1666372 | 1 ML phenylephrine hydrochloride 10 MG/ML Injection | S | Drug | Clinical Drug | RxNorm |
| 300 | Medication | n/a | n/a | EPINEPHRINE 16 MCG/ML COMPOUNDED DRIP WEIGHT-BASED | n/a | wider | equal | missing concentration | 1343916 | 3992 | epinephrine | S | Drug | Ingredient | RxNorm |
| 301 | Medication | n/a | n/a | EPINEPHRINE DRIP - ADULT 64 MCG/ML WEIGHT-BASED | n/a | wider | equal | missing drug concentration, adult, and infusion | 1343916 | 3992 | epinephrine | S | Drug | Ingredient | RxNorm |
| 302 | Medication | n/a | n/a | MILRINONE BOLUS (200 MCG/ML) | n/a | equal | equal | n/a | 42628989 | 1791840 | 100 ML milrinone lactate 0.2 MG/ML Injection | S | Drug | Clinical Drug | RxNorm |
| 303 | Medication | n/a | n/a | MILRINONE DRIP - PEDS STANDARD CONCENTRATION | n/a | equal | equal | n/a | 42628989 | 1791840 | 100 ML milrinone lactate 0.2 MG/ML Injection | S | Drug | Clinical Drug | RxNorm |
| 304 | Medication | n/a | n/a | MILRINONE DRIP - PEDS *HIGH* CONCENTRATION | n/a | equal | equal | n/a | 42628995 | 1791859 | 20 ML milrinone lactate 1 MG/ML Injection | S | Drug | Clinical Drug | RxNorm |
| 305 | Medication | n/a | n/a | MILRINONE BOLUS (1 MG/ML) - PEDS | n/a | equal | equal | n/a | 42628995 | 1791859 | 20 ML milrinone lactate 1 MG/ML Injection | S | Drug | Clinical Drug | RxNorm |
| 306 | Medication | n/a | n/a | EPINEPHRINE DRIP - PED 64 MCG/ML | n/a | wider | equal | missing drug concentration, peds, and infusion | 1343916 | 3992 | epinephrine | S | Drug | Ingredient | RxNorm |
| 307 | Medication | n/a | n/a | INTRAOP VASOPRESSIN + BASE MIXTURE | n/a | equal | equal | n/a | 35202042 | 2103182, 313578 | 1 ML vasopressin (USP) 20 UNT/ML Injection, vasopressin (USP) 20 UNT/ML Injectable Solution | S | Drug | Clinical Drug | RxNorm |
| 308 | Medication | n/a | n/a | INTRAOP VASOPRESSIN + LIDOCAINE MIXTURE | n/a | equal | equal | n/a | 35202042 | 2103182, 313578 | 1 ML vasopressin (USP) 20 UNT/ML Injection, vasopressin (USP) 20 UNT/ML Injectable Solution | S | Drug | Clinical Drug | RxNorm |
| 309 | Medication | n/a | n/a | INTRAOP EPINEPHRINE + BSS PLUS MIXTURE | n/a | wider | equal | missing location intraop and mixture | 1343916 | 3992 | epinephrine | S | Drug | Ingredient | RxNorm |
| 310 | Medication | n/a | n/a | INTRAOP DEXTROSE 50% + EPINEPHRINE + BSS PLUS MIXTUREULAR | n/a | wider | equal | missing location intraop, D50, and mixture | 1343916 | 3992 | epinephrine | S | Drug | Ingredient | RxNorm |
| 311 | Medication | n/a | n/a | INTRAOP PHENYLEPHRINE IV INFUSION | n/a | equal | equal | n/a | 46287545 | 1666372 | 1 ML phenylephrine hydrochloride 10 MG/ML Injection | S | Drug | Clinical Drug | RxNorm |
| 312 | Medication | n/a | n/a | INTRAOP DOBUTAMINE 4000 MCG/ML IV INFUSION | n/a | wider | equal | missing location intraop and concentration | 1337720 | 3616 | dobutamine | S | Drug | Ingredient | RxNorm |
| 313 | Medication | n/a | n/a | INTRAOP DOPAMINE 3200 MCG/ML IV INFUSION | n/a | equal | equal | n/a | 35606567 | 1743938 | 10 ML dopamine hydrochloride 40 MG/ML Injection | S | Drug | Clinical Drug | RxNorm |
| 314 | Medication | n/a | n/a | INTRAOP EPINEPHRINE 64 MCG/ML IV INFUSION | n/a | wider | equal | missing drug concentration and infusion | 1343916 | 3992 | epinephrine | S | Drug | Ingredient | RxNorm |
| 315 | Medication | n/a | n/a | INTRAOP NOREPINEPHRINE 64 MCG/ML IV INFUSION | n/a | wider | equal | missing location intraop and concentration | 1321341 | 7512 | norepinephrine | S | Drug | Ingredient | RxNorm |
| 316 | Medication | n/a | n/a | INTRAOP EPINEPHRINE + BASE MIXTURE | n/a | wider | equal | missing location intraop and mixture | 1343916 | 3992 | epinephrine | S | Drug | Ingredient | RxNorm |
| 317 | Medication | n/a | n/a | INTRAOP VASOPRESSIN 40 UNIT IN NACL 250 ML IV INFUSION | n/a | equal | equal | n/a | 35202042 | 2103182, 313578 | 1 ML vasopressin (USP) 20 UNT/ML Injection, vasopressin (USP) 20 UNT/ML Injectable Solution | S | Drug | Clinical Drug | RxNorm |
| 318 | Medication | n/a | n/a | INTRAOP EPINEPHRINE + BSS REGULAR MIXTURE | n/a | wider | equal | missing location intraop and mixture | 1343916 | 3992 | epinephrine | S | Drug | Ingredient | RxNorm |
| 319 | Medication | n/a | n/a | INTRAOP EPINEPHRINE + GELFOAM SPONGE MIXTURE | n/a | wider | equal | missing location intraop, mixture, and route | 1343916 | 3992 | epinephrine | S | Drug | Ingredient | RxNorm |
| 320 | Medication | n/a | n/a | INTRAOP PHENYLEPHRINE + BASE MIXTURE | n/a | equal | equal | n/a | 46287545 | 1666372 | 1 ML phenylephrine hydrochloride 10 MG/ML Injection | S | Drug | Clinical Drug | RxNorm |
| 321 | Medication | n/a | n/a | INTRAOP EPINEPHRINE + LIDOCAINE MIXTURE | n/a | wider | equal | missing location intraop and mixture | 1343916 | 3992 | epinephrine | S | Drug | Ingredient | RxNorm |
| 322 | Medication | n/a | n/a | INTRAOP EPINEPHRINE 1 MG/ML INJ MULTIPLE SIZE | n/a | wider | equal | missing location intraop | 19076899 | 310132 | epinephrine 1 MG/ML Injectable Solution | S | Drug | Clinical Drug | RxNorm |
| 323 | Medication | n/a | n/a | INTRAOP VASOPRESSIN + BUPIVACAINE MIXTURE | n/a | equal | equal | n/a | 35202042 | 2103182, 313578 | 1 ML vasopressin (USP) 20 UNT/ML Injection, vasopressin (USP) 20 UNT/ML Injectable Solution | S | Drug | Clinical Drug | RxNorm |
| 324 | Medication | n/a | n/a | INTRAOP EPINEPHRINE + BUPIVACAINE MIXTURE | n/a | wider | equal | missing location intraop and mixture | 1343916 | 3992 | epinephrine | S | Drug | Ingredient | RxNorm |

| | | | | | | | | | | | | | | | |
|---|---|---|---|---|---|---|---|---|---|---|---|---|---|---|---|
| 325 | Medication | n/a | n/a | INTRAOP VASOPRESSIN 50 UNITS IN NACL 250 ML IV INFUSION | n/a | equal | equal | n/a | 35202042 | 2103182, 313578 | 1 ML vasopressin (USP) 20 UNT/ML Injection, vasopressin (USP) 20 UNT/ML Injectable Solution | S | Drug | Clinical Drug | RxNorm |
| 326 | Medication | n/a | n/a | INTRAOP EPINEPHRINE + LACTATED RINGERS MIXTURE | n/a | wider | equal | missing location intraop and LR | 1343916 | 3992 | epinephrine | S | Drug | Ingredient | RxNorm |
| 327 | Medication | n/a | n/a | INTRAOP VASOPRESSIN + BUPIVACAINE + BASE MIXTURE | n/a | equal | equal | n/a | 35202042 | 2103182, 313578 | 1 ML vasopressin (USP) 20 UNT/ML Injection, vasopressin (USP) 20 UNT/ML Injectable Solution | S | Drug | Clinical Drug | RxNorm |
| 328 | Medication | n/a | n/a | EPINEPHRINE HCL (PF) 1 MG/ML (1 ML) INJECTION SOLUTION | n/a | equal | equal | n/a | 19076899 | 310132 | epinephrine 1 MG/ML Injectable Solution | S | Drug | Clinical Drug | RxNorm |
| 329 | Medication | n/a | n/a | PHENYLEPHRINE (PF) 1 MG/10 ML (100 MCG/ML) IN 0.9 % NACL IV SYRINGE | n/a | equal | equal | n/a | 42902898 | 1360011 | phenylephrine hydrochloride 0.1 MG/ML Prefilled Syringe | S | Drug | Clinical Drug | RxNorm |
| 330 | Medication | n/a | n/a | MILRINONE 20 MG/100 ML(200 MCG/ML) IN 5 % DEXTROSE INTRAVENOUS PIGGYBK | n/a | equal | equal | n/a | 42628989 | 1791840 | 100 ML milrinone lactate 0.2 MG/ML Injection | S | Drug | Clinical Drug | RxNorm |
| 331 | Medication | n/a | n/a | DOPAMINE 800 MG/250 ML (3,200 MCG/ML) IN 5 % DEXTROSE INTRAVENOUS SOLN | n/a | equal | equal | n/a | 42799685 | 1292887 | 250 ML dopamine hydrochloride 3.2 MG/ML Injection | S | Drug | Clinical Drug | RxNorm |
| 332 | Medication | n/a | n/a | DOPAMINE 400 MG/250 ML (1,600 MCG/ML) IN 5 % DEXTROSE INTRAVENOUS SOLN | n/a | equal | equal | n/a | 35606564 | 1743877 | 250 ML dopamine hydrochloride 1.6 MG/ML Injection | S | Drug | Clinical Drug | RxNorm |
| 333 | Medication | n/a | n/a | PHENYLEPHRINE 0.5 MG/5 ML (100 MCG/ML)IN 0.9 % SOD.CHLORIDE IV SYRINGE | n/a | equal | equal | n/a | 42902898 | 1360011 | phenylephrine hydrochloride 0.1 MG/ML Prefilled Syringe | S | Drug | Clinical Drug | RxNorm |
| 334 | Medication | n/a | n/a | NOREPINEPHRINE BITARTRATE 8 MG/250 ML (32 MCG/ML) IN 0.9 % NACL IV | n/a | wider | equal | missing drug concentration and infusion | 1321341 | 7512 | norepinephrine | S | Drug | Ingredient | RxNorm |
| 335 | Medication | n/a | n/a | NOREPINEPHRINE BITARTRATE 8 MG/250 ML (32 MCG/ML) IN DEXTROSE 5 % IV | n/a | wider | equal | missing drug concentration and infusion | 1321341 | 7512 | norepinephrine | S | Drug | Ingredient | RxNorm |
| 336 | Medication | n/a | n/a | PHENYLEPHRINE 1 MG/10 ML (100 MCG/ML) IN 0.9 % SOD.CHLORIDE IV SYRINGE | n/a | wider | equal | missing drug concentration | 1135766 | 8163 | phenylephrine | S | Drug | Ingredient | RxNorm |
| 337 | Medication | n/a | n/a | PHENYLEPHRINE 20 MG/250 ML (80 MCG/ML) IN 0.9 % SODIUM CHLORIDE IV | n/a | wider | equal | missing drug concentration | 1135766 | 8163 | phenylephrine | S | Drug | Ingredient | RxNorm |
| 338 | Medication | n/a | n/a | EPINEPHRINE HCL 4 MG/250 ML (16 MCG/ML) IN 0.9 % SODIUM CHLORIDE IV | n/a | wider | equal | missing drug concentration | 1343916 | 3992 | epinephrine | S | Drug | Ingredient | RxNorm |
| 339 | Medication | n/a | n/a | EPINEPHRINE HCL 4 MG/250 ML (16 MCG/ML) IN 5 % DEXTROSE INTRAVENOUS | n/a | wider | equal | missing drug concentration and D5 | 1343916 | 3992 | epinephrine | S | Drug | Ingredient | RxNorm |
| 340 | Medication | n/a | n/a | VASOPRESSIN 20 UNIT/ML INTRAVENOUS SOLUTION | n/a | equal | equal | n/a | 1507838 | 313578 | vasopressin (USP) 20 UNT/ML Injectable Solution | S | Drug | Clinical Drug | RxNorm |
| 341 | Medication | n/a | n/a | CYCLOPENTOLATE 1 %-TROPICAM 1 %-PHENYLEPHRINE 2.5 % IN WATER EYE DROPS | n/a | wider | equal | missing drug concentration | 1135766 | 8163 | phenylephrine | S | Drug | Ingredient | RxNorm |
| 342 | Medication | n/a | n/a | DOBUTAMINE 500 MG/250 ML (2,000 MCG/ML) IN 5 % DEXTROSE IV | n/a | equal | equal | n/a | 19076659 | 309986 | 250 ML dobutamine 2 MG/ML Injection | S | Drug | Clinical Drug | RxNorm |
| 343 | Medication | n/a | n/a | INTRAOP EPINEPHRINE 1 MG/ML PF INJ 1 ML AMPULE | n/a | wider | equal | missing location OR | 19076899 | 310132 | epinephrine 1 MG/ML Injectable Solution | S | Drug | Clinical Drug | RxNorm |
| 344 | Medication | n/a | n/a | INTRAOP MUCOSAL ANESTHETIC SOLUTION (LIDOCAINE 4% + PHENYLEPHRINE 1%) (NO CHARGE) | n/a | equal | equal | n/a | 19076899 | 310132 | epinephrine 1 MG/ML Injectable Solution | S | Drug | Clinical Drug | RxNorm |
| 345 | Medication | n/a | n/a | INTRAOP NOREPINEPHRINE BOLUS FROM INFUSION BAG | n/a | wider | equal | missing drug concentration and infusion bolus | 1321341 | 7512 | norepinephrine | S | Drug | Ingredient | RxNorm |
| 346 | Medication | n/a | n/a | MILRINONE 1 MG/ML INTRAVENOUS SOLUTION | n/a | equal | equal | n/a | 42053301 | 1791861 | 50 ML milrinone lactate 1 MG/ML Injection | S | Drug | Clinical Drug | RxNorm |
| 347 | Medication | n/a | n/a | EPINEPHRINE 0.1 MG/ML INJECTION SYRINGE | n/a | equal | equal | n/a | 1344245 | 727373 | 10 ML epinephrine 0.1 MG/ML Prefilled Syringe | S | Drug | Quant Clinical Drug | RxNorm |
| 348 | Medication | n/a | n/a | PHENYLEPHRINE 10 MG/ML INJECTION SOLUTION | n/a | equal | equal | n/a | 42707286 | 1232651 | phenylephrine hydrochloride 10 MG/ML Injectable Solution | S | Drug | Clinical Drug | RxNorm |
| 349 | Medication | n/a | n/a | PHENYLEPHRINE 10 % EYE DROPS | n/a | equal | equal | n/a | 42707558 | 1234571 | phenylephrine hydrochloride 100 MG/ML Ophthalmic Solution | S | Drug | Clinical Drug | RxNorm |
| 350 | Medication | n/a | n/a | PHENYLEPHRINE 2.5 % EYE DROPS | n/a | equal | equal | n/a | 42707562 | 1234579 | phenylephrine hydrochloride 25 MG/ML Ophthalmic Solution | S | Drug | Clinical Drug | RxNorm |
| 351 | Medication | n/a | n/a | PHENYLEPHRINE 0.25 % NASAL SPRAY | n/a | equal | equal | n/a | 42707560 | 1234576 | phenylephrine hydrochloride 2.5 MG/ML Nasal Spray | S | Drug | Clinical Drug | RxNorm |
| 352 | Medication | n/a | n/a | VASOPRESSIN 20 UNIT/ML INJECTION SOLUTION | n/a | equal | equal | n/a | 1507838 | 313578 | vasopressin (USP) 20 UNT/ML Injectable Solution | S | Drug | Clinical Drug | RxNorm |
| 353 | Medication | n/a | n/a | INTRAOP ADV PREP EPINEPHRINE 30 MG IN NACL 0.9% 1 L IRRIGATION | n/a | wider | equal | missing location intraop, NS, and route | 1343916 | 3992 | epinephrine | S | Drug | Ingredient | RxNorm |
| 354 | Medication | n/a | n/a | PHENYLEPHRINE 1 % NASAL SPRAY | n/a | equal | equal | n/a | 40233714 | 1087043 | phenylephrine hydrochloride 10 MG/ML Nasal Spray | S | Drug | Clinical Drug | RxNorm |
| 355 | Medication | n/a | n/a | INTRAOP ADV PREP EPINEPHRINE 2 MG IN LACTATED RINGERS 1000 ML IVPB | n/a | wider | equal | missing location intraop, LR, and IVPB route | 1343916 | 3992 | epinephrine | S | Drug | Ingredient | RxNorm |
| 356 | Medication | n/a | n/a | INTRAOP MILRINONE + HEPARIN + BASE MIXTURE | n/a | equal | equal | n/a | 42628995 | 1791859 | 20 ML milrinone lactate 1 MG/ML Injection | S | Drug | Clinical Drug | RxNorm |
| 357 | Other | n/a | n/a | n/a | n/a | narrower | equal | does not allow for volume - do not use | 44806352 | 801351000000000 | Continuous intravenous infusion of therapeutic substance | S | Procedure | Procedure | SNOMED |
| 358 | Other | n/a | n/a | CONTINUOUS INFUSION | n/a | equal | equal | n/a | 45879918 | LA11260-9 | Infusion, intravenous catheter, continuous | S | Meas Value | Answer | LOINC |
| 359 | Other | n/a | n/a | IV push | n/a | equal | equal | n/a | 45879998 | LA18623-1 | intravenous push | S | Meas Value | Answer | LOINC |
| 360 | Oxygen Device | MECHANICAL VENTILATOR | MECHANICAL VENTILATOR | Mode | %;AC/Pressure control;% | equal | equal | n/a | 37152410 | 1186746005 | Assist control ventilation pressure control inflation-type | S | Procedure | Procedure | SNOMED |
| 361 | Oxygen Device | MECHANICAL VENTILATOR | MECHANICAL VENTILATOR | Mode | %;AC/PRVC;% | equal | equal | n/a | 37152411 | 1186747001 | Assist control ventilation volume-targeted pressure-control inflation-type | S | Procedure | Procedure | SNOMED |
| 362 | Oxygen Device | MECHANICAL VENTILATOR | MECHANICAL VENTILATOR | Mode | %;AC/Volume control;% | equal | equal | n/a | 37152409 | 1186745009 | Assist control ventilation volume-control inflation-type | S | Procedure | Procedure | SNOMED |
| 363 | Oxygen Device | AEROSOL MASK | SUPPLEMENTAL OXYGEN | Device (Oxygen Therapy) | %;aerosol mask;% | equal | equal | n/a | 4145694 | 426851007 | Aerosol oxygen mask | S | Device | Physical Object | SNOMED |
| 364 | Oxygen Device | MECHANICAL VENTILATOR | MECHANICAL VENTILATOR | Mode | %;APRV/Bilevel;% | wider | equal | missing bilevel component | 4072515 | 243143008 | Airway pressure release ventilation | S | Procedure | Procedure | SNOMED |
| 365 | Oxygen Device | MECHANICAL VENTILATOR | MECHANICAL VENTILATOR | Mode | %;ASV;% | wider | equal | missing precise vent mode | 37158404 | 1258985005 | Invasive mechanical ventilation | S | Procedure | Procedure | SNOMED |
| 366 | Oxygen Device | MECHANICAL VENTILATOR | MECHANICAL VENTILATOR | Mode | %;Automode;% | wider | equal | missing precise vent mode | 37158404 | 1258985005 | Invasive mechanical ventilation | S | Procedure | Procedure | SNOMED |
| 367 | Oxygen Device | BAG VALVE MASK | SUPPLEMENTAL OXYGEN: BAG VALVE | Device (Oxygen Therapy) | %;bag valve mask;% | equal | equal | n/a | 4143247 | 425696007 | Respiratory assist, manual, using bag-valve-mask | S | Procedure | Procedure | SNOMED |
| 368 | Oxygen Device | BAG VALVE OETT | SUPPLEMENTAL OXYGEN: BAG VALVE | Device (Oxygen Therapy) | %;bag valve OETT;% | wider | equal | missing Oral ETT route | 4006318 | 11140008 | Respiratory assist, manual | S | Procedure | Procedure | SNOMED |
| 369 | Oxygen Device | MECHANICAL VENTILATOR | MECHANICAL VENTILATOR | Mode | %;Bilevel;% | equal | equal | n/a | 37152361 | 1186618000 | Bilevel artificial ventilation | S | Procedure | Procedure | SNOMED |
| 370 | Oxygen Device | BIPAP | NON-INVASIVE POSITIVE PRESSURE VENTILATION (NIPPV) | Device (Oxygen Therapy) | %;BIPAP full face;% | equal | equal | n/a | 605914 | 1141760009 | Continuous positive airway pressure bilevel positive airway pressure face mask (physical object) | S | Device | Physical Object | SNOMED |
| 371 | Oxygen Device | BIPAP | NON-INVASIVE POSITIVE PRESSURE VENTILATION (NIPPV) | Device (Oxygen Therapy) | %;BIPAP- full face;% | equal | equal | n/a | 605914 | 1141760009 | Continuous positive airway pressure bilevel positive airway pressure face mask (physical object) | S | Device | Physical Object | SNOMED |
| 372 | Oxygen Device | BIPAP | NON-INVASIVE POSITIVE PRESSURE VENTILATION (NIPPV) | Device (Oxygen Therapy) | %;BIPAP nasal;% | equal | equal | n/a | 4138614 | 425826004 | BiPAP oxygen nasal cannula | S | Device | Physical Object | SNOMED |
| 373 | Oxygen Device | BIPAP | NON-INVASIVE POSITIVE PRESSURE VENTILATION (NIPPV) | Device (Oxygen Therapy) | %;BIPAP- nasal;% | equal | equal | n/a | 4138614 | 425826004 | BiPAP oxygen nasal cannula | S | Device | Physical Object | SNOMED |
| 374 | Oxygen Device | BIPAP | NON-INVASIVE POSITIVE PRESSURE VENTILATION (NIPPV) | Mode | %;BIPAP;% | equal | equal | n/a | 4327886 | 429091008 | Dependence on biphasic positive airway pressure (BIPAP) ventilation | S | Condition | Clinical Finding | SNOMED |
| 375 | Oxygen Device | BIPAP | NON-INVASIVE POSITIVE PRESSURE VENTILATION (NIPPV) | Non-invasive | %;BIPAP;% | equal | equal | n/a | 4327886 | 429091008 | Dependence on biphasic positive airway pressure (BIPAP) ventilation | S | Condition | Clinical Finding | SNOMED |
| 376 | Oxygen Device | NON-INVASIVE | SUPPLEMENTAL OXYGEN | Device (Oxygen Therapy) | %;blender system;% | wider | equal | missing precise O2 delivery mode | 42873170 | 931000000000 | Dependence on supplemental oxygen (finding) | S | Condition | Clinical Finding | SNOMED |

| | | | | | | | | | | | | | | | |
|---|---|---|---|---|---|---|---|---|---|---|---|---|---|---|---|
| 377 | Oxygen Device | BLOW BY | SPONTANEOUS BREATHING TRIAL: GENERAL | Device (Oxygen Therapy) | %;blow by;% | wider | equal | missing qualifier blow by | 4308797 | 424139009 | Trial of spontaneous breathing | S | Procedure | Procedure | SNOMED |
| 378 | Oxygen Device | BLOW BY | SPONTANEOUS BREATHING TRIAL: GENERAL | Device (Oxygen Therapy) | %;blow-by;% | wider | equal | missing qualifier blow by | 4308797 | 424139009 | Trial of spontaneous breathing | S | Procedure | Procedure | SNOMED |
| 379 | Oxygen Device | CPAP | SPONTANEOUS BREATHING TRIAL: CPAP | Device (Oxygen Therapy) | %;CPAP full face;% | equal | equal | n/a | 605914 | 1141760009 | Continuous positive airway pressure bilevel positive airway pressure face mask (physical object) | S | Device | Physical Object | SNOMED |
| 380 | Oxygen Device | CPAP | SPONTANEOUS BREATHING TRIAL: CPAP | Device (Oxygen Therapy) | %;CPAP- full face;% | equal | equal | n/a | 605914 | 1141760009 | Continuous positive airway pressure bilevel positive airway pressure face mask (physical object) | S | Device | Physical Object | SNOMED |
| 381 | Oxygen Device | CPAP | SPONTANEOUS BREATHING TRIAL: CPAP | Device (Oxygen Therapy) | %;CPAP nasal;% | wider | equal | missing spontaneous breathing trial | 45761494 | 467645007 | CPAP nasal oxygen cannula | S | Device | Physical Object | SNOMED |
| 382 | Oxygen Device | CPAP | SPONTANEOUS BREATHING TRIAL: CPAP | Device (Oxygen Therapy) | %;CPAP- nasal;% | wider | equal | missing spontaneous breathing trial, nasal | 45761494 | 467645007 | CPAP nasal oxygen cannula | S | Device | Physical Object | SNOMED |
| 383 | Oxygen Device | CPAP | SPONTANEOUS BREATHING TRIAL: CPAP | Mode | %;CPAP;% | wider | equal | missing spontaneous breathing trial | 45761494 | 467645007 | CPAP nasal oxygen cannula | S | Device | Physical Object | SNOMED |
| 384 | Oxygen Device | CPAP | SPONTANEOUS BREATHING TRIAL: CPAP | Device (Oxygen Therapy) | %;CPAP;% | wider | equal | missing spontaneous breathing trial | 45761494 | 467645007 | CPAP nasal oxygen cannula | S | Device | Physical Object | SNOMED |
| 385 | Oxygen Device | CPAP | NON-INVASIVE POSITIVE PRESSURE VENTILATION (NIPPV) | Non-invasive | %;CPAP;% | equal | equal | n/a | 40488354 | 446573003 | CPAP (continuous positive airway pressure) titration | S | Procedure | Procedure | SNOMED |
| 386 | Oxygen Device | NON-INVASIVE | SUPPLEMENTAL OXYGEN | Device (Oxygen Therapy) | %;enclosed incubator;% | wider | equal | missing precise O2 delivery mode | 42873170 | 931000000000 | Dependence on supplemental oxygen (finding) | S | Condition | Clinical Finding | SNOMED |
| 387 | Oxygen Device | SIMPLE FACE MASK | SUPPLEMENTAL OXYGEN | Device (Oxygen Therapy) | %;face tent;% | equal | equal | n/a | 45759373 | 464578003 | Oxygen administration face tent | S | Device | Physical Object | SNOMED |
| 388 | Oxygen Device | BIPAP | NON-INVASIVE POSITIVE PRESSURE VENTILATION (NIPPV) | Device (Oxygen Therapy) | %;Full Face BIPAP;% | equal | equal | n/a | 605914 | 1141760009 | Continuous positive airway pressure bilevel positive airway pressure face mask (physical object) | S | Device | Physical Object | SNOMED |
| 389 | Oxygen Device | BIPAP | NON-INVASIVE POSITIVE PRESSURE VENTILATION (NIPPV) | Device (Oxygen Therapy) | %;Full Face BIPAP;% | equal | equal | n/a | 605914 | 1141760009 | Continuous positive airway pressure bilevel positive airway pressure face mask (physical object) | S | Device | Physical Object | SNOMED |
| 390 | Oxygen Device | BIPAP | NON-INVASIVE POSITIVE PRESSURE VENTILATION (NIPPV) | Device (Oxygen Therapy) | %;Full Face BIPAP;% | equal | equal | n/a | 605914 | 1141760009 | Continuous positive airway pressure bilevel positive airway pressure face mask (physical object) | S | Device | Physical Object | SNOMED |
| 391 | Oxygen Device | CPAP | SPONTANEOUS BREATHING TRIAL: CPAP | Device (Oxygen Therapy) | %;full face CPAP;% | equal | equal | n/a | 605914 | 1141760009 | Continuous positive airway pressure bilevel positive airway pressure face mask (physical object) | S | Device | Physical Object | SNOMED |
| 392 | Oxygen Device | HIGH FLOW NASAL CANNULA | HIGH FLOW NASAL CANNULA | Device (Oxygen Therapy) | %;heated HFNC system;% | equal | equal | n/a | 37158406 | 1259025002 | Heated and humidified high flow oxygen therapy using nasal cannula | S | Procedure | Procedure | SNOMED |
| 393 | Oxygen Device | n/a | n/a | Device (Oxygen Therapy) | %;heated;% | narrower | equal | adds humidified | 3655950 | 870533002 | Heated and humidified high flow oxygen therapy (procedure) | S | Procedure | Procedure | SNOMED |
| 394 | Oxygen Device | MECHANICAL VENTILATOR | MECHANICAL VENTILATOR | Mode | %;HFJV;% | equal | equal | n/a | 4055377 | 243154003 | High frequency jet ventilation | S | Procedure | Procedure | SNOMED |
| 395 | Oxygen Device | HIGH FLOW NASAL CANNULA | HIGH FLOW NASAL CANNULA | Mode | %;High Flow Nasal Cannula;% | equal | equal | n/a | 4139525 | 426854004 | High flow oxygen nasal cannula (physical object) | S | Device | Physical Object | SNOMED |
| 396 | Oxygen Device | NON REBREATHER MASK | SUPPLEMENTAL OXYGEN | Device (Oxygen Therapy) | %;high-flow mask;% | wider | equal | missing type of mask | 4162736 | 371908008 | Oxygen administration by mask (procedure) | S | Procedure | Procedure | SNOMED |
| 397 | Oxygen Device | HIGH FLOW NASAL CANNULA | HIGH FLOW NASAL CANNULA | Device (Oxygen Therapy) | %;high-flow nasal cannula;% | equal | equal | n/a | 4139525 | 426854004 | High flow oxygen nasal cannula (physical object) | S | Device | Procedure | SNOMED |
| 398 | Oxygen Device | n/a | n/a | Device (Oxygen Therapy) | %;humidified;% | equal | equal | n/a | 4119964 | 304577004 | Humidified oxygen | S | Procedure | Procedure | SNOMED |
| 399 | Oxygen Device | BAG VALVE MASK | SUPPLEMENTAL OXYGEN: BAG VALVE | Device (Oxygen Therapy) | %;manual resuscitator;% | equal | equal | n/a | 4143247 | 425696007 | Respiratory assist, manual, using bag-valve-mask | S | Procedure | Procedure | SNOMED |
| 400 | Oxygen Device | MECHANICAL VENTILATOR | MECHANICAL VENTILATOR | Device (Oxygen Therapy) | %;mechanical ventilator;% | equal | equal | n/a | 37158404 | 1258985005 | Invasive mechanical ventilation | S | Procedure | Procedure | SNOMED |
| 401 | Oxygen Device | NON-INVASIVE | SUPPLEMENTAL OXYGEN | Device (Oxygen Therapy) | %;mist tent;% | equal | equal | n/a | 4138190 | 425994007 | Oxygen mist tent (physical object) | S | Device | Physical Object | SNOMED |
| 402 | Oxygen Device | BIPAP | NON-INVASIVE POSITIVE PRESSURE VENTILATION (NIPPV) | Device (Oxygen Therapy) | %;Nasal BIPAP;% | equal | equal | n/a | 4138614 | 425826004 | BiPAP oxygen nasal cannula | S | Device | Physical Object | SNOMED |
| 403 | Oxygen Device | BIPAP | NON-INVASIVE POSITIVE PRESSURE VENTILATION (NIPPV) | Device (Oxygen Therapy) | %;Nasal BIPAP;% | equal | equal | n/a | 4138614 | 425826004 | BiPAP oxygen nasal cannula | S | Device | Physical Object | SNOMED |
| 404 | Oxygen Device | BIPAP | NON-INVASIVE POSITIVE PRESSURE VENTILATION (NIPPV) | Device (Oxygen Therapy) | %;Nasal BIPAP;% | equal | equal | n/a | 4138614 | 425826004 | BiPAP oxygen nasal cannula | S | Device | Physical Object | SNOMED |
| 405 | Oxygen Device | SIMPLE NASAL CANNULA | SUPPLEMENTAL OXYGEN | Device (Oxygen Therapy) | %;nasal cannula w/ humidification;% | wider | equal | missing humidification | 4155151 | 371907003 | Oxygen administration by nasal cannula (procedure) | S | Procedure | Procedure | SNOMED |
| 406 | Oxygen Device | SIMPLE NASAL CANNULA | SUPPLEMENTAL OXYGEN | Device (Oxygen Therapy) | %;nasal cannula w/ reservoir (i.e. oximizer);% | wider | equal | missing resevoir | 4155151 | 371907003 | Oxygen administration by nasal cannula (procedure) | S | Procedure | Procedure | SNOMED |
| 407 | Oxygen Device | SIMPLE NASAL CANNULA | SUPPLEMENTAL OXYGEN | Device (Oxygen Therapy) | %;nasal cannula with humidification;% | wider | equal | missing humidification | 4155151 | 371907003 | Oxygen administration by nasal cannula (procedure) | S | Procedure | Procedure | SNOMED |
| 408 | Oxygen Device | SIMPLE NASAL CANNULA | SUPPLEMENTAL OXYGEN | Device (Oxygen Therapy) | %;nasal cannula with reservoir (i.e. oximizer);% | wider | equal | missing resevoir | 4155151 | 371907003 | Oxygen administration by nasal cannula (procedure) | S | Procedure | Procedure | SNOMED |
| 409 | Oxygen Device | SIMPLE NASAL CANNULA | SUPPLEMENTAL OXYGEN | Device (Oxygen Therapy) | %;nasal cannula with reservoir (Oximizer);% | wider | equal | missing resevoir | 4155151 | 371907003 | Oxygen administration by nasal cannula (procedure) | S | Procedure | Procedure | SNOMED |
| 410 | Oxygen Device | SIMPLE NASAL CANNULA | SUPPLEMENTAL OXYGEN | Device (Oxygen Therapy) | %;nasal cannula;% | equal | equal | n/a | 4155151 | 371907003 | Oxygen administration by nasal cannula (procedure) | S | Procedure | Procedure | SNOMED |
| 411 | Oxygen Device | CPAP | SPONTANEOUS BREATHING TRIAL: CPAP | Device (Oxygen Therapy) | %;nasal CPAP;% | wider | equal | missing spontaneous breathing trial, nasal | 45761494 | 467645007 | CPAP nasal oxygen cannula | S | Device | Physical Object | SNOMED |
| 412 | Oxygen Device | NON-INVASIVE | NON-INVASIVE POSITIVE PRESSURE VENTILATION (NIPPV) | Device (Oxygen Therapy) | %;nasal prongs;% | wider | equal | missing nasal prongs as route | 40486624 | 447837008 | Non-invasive positive pressure ventilation | S | Procedure | Procedure | SNOMED |
| 413 | Oxygen Device | NIV | NON-INVASIVE POSITIVE PRESSURE VENTILATION (NIPPV) | Mode | %;NIPPV;% | equal | equal | n/a | 40486624 | 447837008 | Non-invasive positive pressure ventilation (regime/therapy) | S | Procedure | Procedure | SNOMED |
| 414 | Oxygen Device | NIV | NON-INVASIVE POSITIVE PRESSURE VENTILATION (NIPPV) | Mode | %;NIV/BIPAP;% | narrower | equal | adds nasal cannula | 4138614 | 425826004 | BiPAP oxygen nasal cannula | S | Device | Physical Object | SNOMED |
| 415 | Oxygen Device | NIV | NON-INVASIVE POSITIVE PRESSURE VENTILATION (NIPPV) | Mode | %;NIV/CPAP;% | wider | equal | missing CPAP | 40486624 | 447837008 | Non-invasive positive pressure ventilation (regime/therapy) | S | Procedure | Procedure | SNOMED |
| 416 | Oxygen Device | NIV | NON-INVASIVE POSITIVE PRESSURE VENTILATION (NIPPV) | Mode | %;NIV/PC;% | wider | equal | missing pressure control | 40486624 | 447837008 | Non-invasive positive pressure ventilation (regime/therapy) | S | Procedure | Procedure | SNOMED |
| 417 | Oxygen Device | NIV | NON-INVASIVE POSITIVE PRESSURE VENTILATION (NIPPV) | Mode | %;NIV/PS;% | wider | equal | missing NIV | 40486624 | 447837008 | Non-invasive positive pressure ventilation (regime/therapy) | S | Procedure | Procedure | SNOMED |
| 418 | Oxygen Device | NIV | NON-INVASIVE POSITIVE PRESSURE VENTILATION (NIPPV) | Mode | %;NIV/ST;% | narrower | equal | adds PPV | 40486624 | 447837008 | Non-invasive positive pressure ventilation (regime/therapy) | S | Procedure | Procedure | SNOMED |
| 419 | Oxygen Device | NIV | NON-INVASIVE POSITIVE PRESSURE VENTILATION (NIPPV) | Mode | %;NIV;% | narrower | equal | adds PPV | 40486624 | 447837008 | Non-invasive positive pressure ventilation (regime/therapy) | S | Procedure | Procedure | SNOMED |
| 420 | Oxygen Device | NON REBREATHER MASK | SUPPLEMENTAL OXYGEN | Device (Oxygen Therapy) | %;nonrebreather mask;% | equal | equal | n/a | 44790731 | 240051000000000 | Oxygen administration by non rebreather mask (procedure) | S | Procedure | Procedure | SNOMED |
| 421 | Oxygen Device | BIPAP | NON-INVASIVE POSITIVE PRESSURE VENTILATION (NIPPV) | Device (Oxygen Therapy) | %;NPPV/NIV;% | equal | equal | n/a | 40486624 | 447837008 | Non-invasive positive pressure ventilation (regime/therapy) | S | Procedure | Procedure | SNOMED |
| 422 | Oxygen Device | NON-INVASIVE | SUPPLEMENTAL OXYGEN | Device (Oxygen Therapy) | %;O2 ring;% | wider | equal | missing ring | 42873170 | 931000000000 | Dependence on supplemental oxygen (finding) | S | Condition | Clinical Finding | SNOMED |
| 423 | Oxygen Device | SIMPLE FACE MASK | SUPPLEMENTAL OXYGEN | Device (Oxygen Therapy) | %;open oxygen mask;% | wider | equal | missing type of mask | 4162736 | 371908008 | Oxygen administration by mask (procedure) | S | Procedure | Procedure | SNOMED |
| 424 | Oxygen Device | MECHANICAL VENTILATOR | MECHANICAL VENTILATOR | Mode | %;Oscillator;% | narrower | equal | adds high frequency | 4074666 | 243155002 | High frequency oscillatory ventilation | S | Procedure | Procedure | SNOMED |
| 425 | Oxygen Device | OTHER | SPONTANEOUS BREATHING TRIAL: GENERAL | Mode | %;Other (non-invasive) Mode;% | equal | equal | n/a | 40486624 | 447837008 | Non-invasive positive pressure ventilation (regime/therapy) | S | Procedure | Procedure | SNOMED |
| 426 | Oxygen Device | MECHANICAL VENTILATOR OR OTHER | SPONTANEOUS BREATHING TRIAL: GENERAL | Device (Oxygen Therapy) | %;other (see comments);% | wider | equal | missing precise vent mode, spontaneous breathing trial | 37158404 | 1258985005 | Invasive mechanical ventilation | S | Procedure | Procedure | SNOMED |
| 427 | Oxygen Device | MECHANICAL VENTILATOR | MECHANICAL VENTILATOR | Mode | %;Other Invasive Vent Mode;% | wider | equal | missing precise vent mode | 37158404 | 1258985005 | Invasive mechanical ventilation | S | Procedure | Procedure | SNOMED |
| 428 | Oxygen Device | MECHANICAL VENTILATOR OR OTHER | SPONTANEOUS BREATHING TRIAL: GENERAL | Mode | %;Other;% | wider | equal | missing precise vent mode, spontaneous breathing trial | 37158404 | 1258985005 | Invasive mechanical ventilation | S | Procedure | Procedure | SNOMED |
| 429 | Oxygen Device | NON-INVASIVE | SUPPLEMENTAL OXYGEN | Device (Oxygen Therapy) | %;oxygen hood;% | equal | equal | n/a | 45759376 | 464581008 | Oxygen administration hood, adult | S | Device | Physical Object | SNOMED |
| 430 | Oxygen Device | NON-INVASIVE | SUPPLEMENTAL OXYGEN | Device (Oxygen Therapy) | %;oxygen tent;% | equal | equal | n/a | 4070683 | 20428008 | Oxygen tent | S | Device | Physical Object | SNOMED |
| 431 | Oxygen Device | NON-INVASIVE | SUPPLEMENTAL OXYGEN | Device (Oxygen Therapy) | %;oxyhood;% | equal | equal | n/a | 45759376 | 464581008 | Oxygen administration hood, adult | S | Device | Physical Object | SNOMED |
| 432 | Oxygen Device | NON REBREATHER MASK | SUPPLEMENTAL OXYGEN | Device (Oxygen Therapy) | %;partial rebreather mask;% | wider | equal | missing type of mask | 4162736 | 371908008 | Oxygen administration by mask (procedure) | S | Procedure | Procedure | SNOMED |
| 433 | Oxygen Device | MECHANICAL VENTILATOR | MECHANICAL VENTILATOR | Mode | %;PAV;% | wider | equal | missing precise vent mode | 37158404 | 1258985005 | Invasive mechanical ventilation | S | Procedure | Procedure | SNOMED |
| 434 | Oxygen Device | MECHANICAL VENTILATOR | MECHANICAL VENTILATOR | Mode | %;Pressure Support/CPAP;% | wider | equal | missing pressure support/CPAP | 37154096 | 1259864003 | Pressure support ventilation | S | Procedure | Procedure | SNOMED |
| 435 | Oxygen Device | MECHANICAL VENTILATOR | MECHANICAL VENTILATOR | Mode | %;Pressure Support;% | equal | equal | n/a | 37154096 | 1259864003 | Pressure support ventilation | S | Procedure | Procedure | SNOMED |
| 436 | Oxygen Device | ROOM AIR | ROOM AIR | Device (Oxygen Therapy) | %;room air;% | equal | equal | n/a | 36716647 | 722742002 | Breathing room air (finding) | S | Condition | Clinical Finding | SNOMED |
| 437 | Oxygen Device | SIMPLE FACE MASK | SUPPLEMENTAL OXYGEN | Device (Oxygen Therapy) | %;simple face mask;% | wider | equal | missing type of mask | 4162736 | 371908008 | Oxygen administration by mask (procedure) | S | Procedure | Procedure | SNOMED |
| 438 | Oxygen Device | MECHANICAL VENTILATOR | MECHANICAL VENTILATOR | Mode | %;SIMV Pressure control/pressure support;% | equal | equal | n/a | 37152364 | 1186622005 | Synchronized intermittent mandatory ventilation - pressure-control pressure-support inflation | S | Procedure | Procedure | SNOMED |

| | | | | | | | | | | | | | | | |
|---|---|---|---|---|---|---|---|---|---|---|---|---|---|---|---|
| 439 | Oxygen Device | MECHANICAL VENTILATOR | MECHANICAL VENTILATOR | Mode | %;SIMV/Pressure control;% | narrower | equal | adds detail | 37152388 | 1186683003 | Synchronized intermittent mandatory ventilation with pressure-control, an assured constant airway pressure adjunct and pressure-support at the higher pressure level only | S | Procedure | Procedure | SNOMED |
| 440 | Oxygen Device | MECHANICAL VENTILATOR | MECHANICAL VENTILATOR | Mode | %;SIMV/PRVC/Pressure support;% | equal | equal | n/a | 37152384 | 1186679003 | Synchronized intermittent mandatory ventilation - volume targeted pressure-control and pressure-support inflation | S | Procedure | Procedure | SNOMED |
| 441 | Oxygen Device | MECHANICAL VENTILATOR | MECHANICAL VENTILATOR | Mode | %;SIMV/PRVC;% | equal | equal | n/a | 37152384 | 1186679003 | Synchronized intermittent mandatory ventilation - volume targeted pressure-control and pressure-support inflation | S | Procedure | Procedure | SNOMED |
| 442 | Oxygen Device | MECHANICAL VENTILATOR | MECHANICAL VENTILATOR | Mode | %;SIMV/Volume control/Pressure support;% | equal | equal | n/a | 37152363 | 1186621003 | Synchronized intermittent mandatory ventilation - volume control pressure-support inflation | S | Procedure | Procedure | SNOMED |
| 443 | Oxygen Device | MECHANICAL VENTILATOR | MECHANICAL VENTILATOR | Mode | %;SIMV/Volume control;% | narrower | equal | adds detail | 37152363 | 1186621003 | Synchronized intermittent mandatory ventilation - volume control pressure-support inflation | S | Procedure | Procedure | SNOMED |
| 444 | Oxygen Device | MECHANICAL VENTILATOR | MECHANICAL VENTILATOR | Mode | %;Spontaneous;% | wider | equal | missing vent mode spontaneous | 37158404 | 1258985005 | Invasive mechanical ventilation | S | Procedure | Procedure | SNOMED |
| 445 | Oxygen Device | TRACHEOSTOMY COLLAR/TRACH MIST/T-PIECE | SPONTANEOUS BREATHING TRIAL: GENERAL | Device (Oxygen Therapy) | %;T - piece;% | wider | equal | missing spontaneus breathing trial | 4188570 | 415700003 | T piece without bag | S | Device | Physical Object | SNOMED |
| 446 | Oxygen Device | TRACHEOSTOMY COLLAR/TRACH MIST/T-PIECE | SPONTANEOUS BREATHING TRIAL: GENERAL | Device (Oxygen Therapy) | %;T - piece;% | wider | equal | missing spontaneous breathing trial | 4188569 | 415698007 | T piece with bag | S | Device | Physical Object | SNOMED |
| 447 | Oxygen Device | TRACHEOSTOMY COLLAR/TRACH MIST/T-PIECE | SPONTANEOUS BREATHING TRIAL: GENERAL | Device (Oxygen Therapy) | %;T - piece;% | wider | equal | missing spontaneous breathing trial | 4188569 | 415698007 | T piece with bag | S | Device | Physical Object | SNOMED |
| 448 | Oxygen Device | TRACHEOSTOMY COLLAR/TRACH MIST/T-PIECE | SPONTANEOUS BREATHING TRIAL: GENERAL | Device (Oxygen Therapy) | %;trach mist;% | wider | equal | missing trach or mist, spontaneous breathing trial | 42873170 | 931000000000 | Dependence on supplemental oxygen (finding) | S | Condition | Clinical Finding | SNOMED |
| 449 | Oxygen Device | TRACHEOSTOMY COLLAR/TRACH MIST/T-PIECE | SPONTANEOUS BREATHING TRIAL: GENERAL | Device (Oxygen Therapy) | %;tracheostomy collar;% | wider | equal | missing trach or collar, spontaneous breathing trial | 42873170 | 931000000000 | Dependence on supplemental oxygen (finding) | S | Condition | Clinical Finding | SNOMED |
| 450 | Oxygen Device | OTHER | SUPPLEMENTAL OXYGEN | Device (Oxygen Therapy) | %;transtracheal catheter;% | wider | wider | missing transtracheal catheter | 42873170 | 931000000000 | Dependence on supplemental oxygen (finding) | S | Condition | Clinical Finding | SNOMED |
| 451 | Oxygen Device | CONDITIONAL MECHANICAL VENTILATOR: VAPS | CONDITIONAL MECHANICAL VENTILATOR: VAPS | Mode | %;VAPS;% | wider | wider | missing precise mode | 42873170 | 931000000000 | Dependence on supplemental oxygen (finding) | S | Condition | Clinical Finding | SNOMED |
| 452 | Oxygen Device | MECHANICAL VENTILATOR | MECHANICAL VENTILATOR | Device (Oxygen Therapy) | %;ventilator;% | equal | equal | n/a | 37158404 | 1258985005 | Invasive mechanical ventilation | S | Procedure | Procedure | SNOMED |
| 453 | Oxygen Device | VENTURI MASK | SUPPLEMENTAL OXYGEN | Device (Oxygen Therapy) | %;Venturi mask system;% | equal | equal | n/a | 4323763 | 429253002 | Oxygen administration by Venturi mask (procedure) | S | Procedure | Procedure | SNOMED |
| 454 | Oxygen Device | VENTURI MASK | SUPPLEMENTAL OXYGEN | Device (Oxygen Therapy) | %;Venturi mask;% | equal | equal | n/a | 4323763 | 429253002 | Oxygen administration by Venturi mask (procedure) | S | Procedure | Procedure | SNOMED |
| 455 | Oxygen Device | MECHANICAL VENTILATOR | MECHANICAL VENTILATOR | Mode | %;Volume Guarantee/SIMV + PS;% | narrower | equal | adds detail | 37152384 | 1186679003 | Synchronized intermittent mandatory ventilation - volume targeted pressure-control and pressure-support inflation | S | Procedure | Procedure | SNOMED |
| 456 | Oxygen Device | MECHANICAL VENTILATOR | MECHANICAL VENTILATOR | Mode | %;Volume Guarantee/VC;% | wider | equal | missing precise vent mode | 37158404 | 1258985005 | Invasive mechanical ventilation | S | Procedure | Procedure | SNOMED |
| 457 | Oxygen Device | MECHANICAL VENTILATOR | MECHANICAL VENTILATOR | Mode | %;Volume Support;% | wider | equal | missing precise vent mode | 37158404 | 1258985005 | Invasive mechanical ventilation | S | Procedure | Procedure | SNOMED |
| 458 | Oxygen Device | MECHANICAL VENTILATOR | MECHANICAL VENTILATOR | Invasive Ventilation Started | %;Yes;% | wider | equal | missing precise vent mode | 37158404 | 1258985005 | Invasive mechanical ventilation | S | Procedure | Procedure | SNOMED |
| 459 | Oxygen Device | NON-INVASIVE | SUPPLEMENTAL OXYGEN | Invasive Ventilation Stopped | %;Yes;% | equal | equal | n/a | 42873170 | 931000000000 | Dependence on supplemental oxygen (finding) | S | Condition | Clinical Finding | SNOMED |
| 460 | Race | n/a | n/a | American Indian or Alaska Native | n/a | equal | equal | n/a | 8657 | 1 | American Indian or Alaska Native | S | Race | Race | Race |
| 461 | Race | n/a | n/a | Asian | n/a | equal | equal | n/a | 8515 | 2 | Asian | S | Race | Race | Race |
| 462 | Race | n/a | n/a | Other | n/a | equal | equal | n/a | 8522 | 9 | Other Race | S | Race | Race | Race |
| 463 | Race | n/a | n/a | Unavailable or unknown | n/a | wider | equal | missing precision with race sub category | 8552 | 9 | Other Race | S | Race | Race | Race |
| 464 | Race | n/a | n/a | Haitian (Inactive) | n/a | wider | equal | missing precision with race sub category | 8522 | 9 | Other Race | S | Race | Race | Race |
| 465 | Race | n/a | n/a | Filipino (Inactive) | n/a | wider | equal | missing precision with race sub category | 8515 | 2 | Asian | S | Race | Race | Race |
| 466 | Race | n/a | n/a | Cuban (Inactive) | n/a | wider | equal | missing precision with race sub category | 8522 | 9 | Other Race | S | Race | Race | Race |
| 467 | Race | n/a | n/a | Chinese (Inactive) | n/a | wider | equal | missing precision with race sub category | 8515 | 2 | Asian | S | Race | Race | Race |
| 468 | Race | n/a | n/a | Mexican American (Inactive) | n/a | wider | equal | missing precision with race sub category | 8522 | 9 | Other Race | S | Race | Race | Race |
| 469 | Race | n/a | n/a | Japanese (Inactive) | n/a | wider | equal | missing precision with race sub category | 8515 | 2 | Asian | S | Race | Race | Race |
| 470 | Race | n/a | n/a | Korean (Inactive) | n/a | wider | equal | missing precision with race sub category | 8515 | 2 | Asian | S | Race | Race | Race |
| 471 | Race | n/a | n/a | African American or Black | n/a | equal | equal | n/a | 8516 | 3 | Black or African American | S | Race | Race | Race |
| 472 | Race | n/a | n/a | Laotian/Hmong (Inactive) | n/a | wider | equal | missing precision with race sub category | 8515 | 2 | Asian | S | Race | Race | Race |
| 473 | Race | n/a | n/a | N. African Non-Specific (Inactive) | n/a | wider | equal | missing precision with race sub category | 8516 | 3 | Black or African American | S | Race | Race | Race |
| 474 | Race | n/a | n/a | Pakistani (Inactive) | n/a | wider | equal | missing precision with race sub category | 8515 | 2 | Asian | S | Race | Race | Race |
| 475 | Race | n/a | n/a | OTHER HISPANIC (INACTIVE) | n/a | wider | equal | missing precision with race sub category | 8522 | 9 | Other Race | S | Race | Race | Race |
| 476 | Race | n/a | n/a | Thai (Inactive) | n/a | wider | equal | missing precision with race sub category | 8515 | 2 | Asian | S | Race | Race | Race |
| 477 | Race | n/a | n/a | Vietnamese (Inactive) | n/a | wider | equal | missing precision with race sub category | 8515 | 2 | Asian | S | Race | Race | Race |
| 478 | Race | n/a | n/a | Iranian (Inactive) | n/a | wider | equal | missing precision with race sub category | 8527 | 5 | White | S | Race | Race | Race |
| 479 | Race | n/a | n/a | Cambodian (Inactive) | n/a | wider | equal | missing precision with race sub category | 8515 | 2 | Asian | S | Race | Race | Race |
| 480 | Race | n/a | n/a | Afgan (Inactive) | n/a | wider | equal | missing precision with race sub category | 8527 | 5 | White | S | Race | Race | Race |
| 481 | Race | n/a | n/a | Indo-Chinese Non-Specific (Inactive) | n/a | wider | equal | missing precision with race sub category | 8515 | 2 | Asian | S | Race | Race | Race |
| 482 | Race | n/a | n/a | Unable to respond | n/a | wider | equal | missing precision with race sub category | 8552 | 9 | Unknown | S | Race | Race | Race |
| 483 | Race | n/a | n/a | Native Hawaiian or other Pacific Islander | n/a | equal | equal | n/a | 8557 | 4 | Native Hawaiian or Other Pacific Islander | S | Race | Race | Race |
| 484 | Race | n/a | n/a | White | n/a | equal | equal | n/a | 8527 | 5 | White | S | Race | Race | Race |
| 485 | Race | n/a | n/a | Decline to state | n/a | wider | equal | missing precision with race sub category | 8552 | 9 | Unknown | S | Race | Race | Race |
| 486 | Surgical Specialty | n/a | n/a | Surgical Oncology | n/a | wider | equal | does not map to specific site service line name | 1074156 | 1335967008 | Oncology Service (Oncology) | S | Observation | Qualifier Value | SNOMED |
| 487 | Surgical Specialty | n/a | n/a | Ophthalmology | n/a | equal | equal | n/a | 4125853 | 305895002 | Seen by Opthamology Service (HEENT) | S | Observation | Clinical Finding | SNOMED |
| 488 | Surgical Specialty | n/a | n/a | Ortho Trauma | n/a | wider | equal | does not map to specific site service line name | 4150872 | 310161006 | Orthopedic Service (Ortho) | S | Observation | Qualifier Value | SNOMED |
| 489 | Surgical Specialty | n/a | n/a | Orthopedics | n/a | equal | equal | n/a | 4150872 | 310161006 | Orthopedic Service (Ortho) | S | Observation | Qualifier Value | SNOMED |
| 490 | Surgical Specialty | n/a | n/a | Peds Surgery | n/a | wider | equal | does not map to specific site service line name | 4139904 | 305891006 | Seen by General Surgery Service (General Surgery) | S | Observation | Clinical Finding | SNOMED |
| 491 | Surgical Specialty | n/a | n/a | Peds CVT | n/a | wider | equal | does not map to specific site service line name | 37397651 | 718348005 | Cardiovascular Care (Cardiovascular) | S | Observation | Procedure | SNOMED |
| 492 | Surgical Specialty | n/a | n/a | Peds GI | n/a | wider | equal | does not map to specific site service line name | 4139904 | 305891006 | Seen by General Surgery Service (General Surgery) | S | Observation | Clinical Finding | SNOMED |
| 493 | Surgical Specialty | n/a | n/a | Peds Hem Onc | n/a | wider | equal | does not map to specific site service line name | 1074156 | 1335967008 | Oncology Service (Oncology) | S | Observation | Qualifier Value | SNOMED |
| 494 | Surgical Specialty | n/a | n/a | Peds Ophth | n/a | wider | equal | does not map to specific site service line name | 4125853 | 305895002 | Seen by Opthamology Service (HEENT) | S | Observation | Clinical Finding | SNOMED |
| 495 | Surgical Specialty | n/a | n/a | Peds Urology | n/a | wider | equal | does not map to specific site service line name | 4215134 | 394612005 | Urology (Urology and Gynecology) | S | Observation | Qualifier Value | SNOMED |
| 496 | Surgical Specialty | n/a | n/a | Pain Management | n/a | equal | equal | n/a | 4192791 | 394882004 | Pain Management (specialty) | S | Observation | Qualifier Value | SNOMED |
| 497 | Surgical Specialty | n/a | n/a | Plastics | n/a | wider | equal | does not map to specific site service line name | 4139904 | 305891006 | Seen by General Surgery Service (General Surgery) | S | Observation | Clinical Finding | SNOMED |
| 498 | Surgical Specialty | n/a | n/a | Transplant | n/a | wider | equal | does not map to specific site service line name | 4139904 | 305891006 | Seen by General Surgery Service (General Surgery) | S | Observation | Clinical Finding | SNOMED |
| 499 | Surgical Specialty | n/a | n/a | ACS | n/a | wider | equal | does not map to specific site service line name | 4139904 | 305891006 | Seen by General Surgery Service (General Surgery) | S | Observation | Clinical Finding | SNOMED |
| 500 | Surgical Specialty | n/a | n/a | Urology | n/a | equal | equal | n/a | 4215134 | 394612005 | Urology (Urology and Gynecology) | S | Observation | Qualifier Value | SNOMED |
| 501 | Surgical Specialty | n/a | n/a | Vascular | n/a | wider | equal | does not map to specific site service line name | 37397651 | 718348005 | Cardiovascular Care (Cardiovascular) | S | Observation | Procedure | SNOMED |
| 502 | Surgical Specialty | n/a | n/a | Radiology | n/a | equal | equal | n/a | 4150863 | 310125001 | Radiology Service (Other) | S | Observation | Qualifier Value | SNOMED |
| 503 | Surgical Specialty | n/a | n/a | Neurology | n/a | equal | equal | n/a | 4138361 | 305894003 | Neurology (Neurosurgery) | S | Observation | Qualifier Value | SNOMED |
| 504 | Surgical Specialty | n/a | n/a | Cardiology | n/a | wider | equal | does not map to specific site service line name | 37397651 | 718348005 | Cardiovascular Care (Cardiovascular) | S | Observation | Procedure | SNOMED |
| 505 | Surgical Specialty | n/a | n/a | Dental | n/a | equal | equal | n/a | 4148674 | 310143002 | Dental Surgery Service (HEENT) | S | Observation | Qualifier Value | SNOMED |
| 506 | Surgical Specialty | n/a | n/a | Burn | n/a | wider | equal | does not map to specific site service line name | 4139904 | 305891006 | Seen by General Surgery Service (General Surgery) | S | Observation | Clinical Finding | SNOMED |
| 507 | Surgical Specialty | n/a | n/a | Obstetrics/GYN | n/a | equal | equal | n/a | 4215122 | 394585009 | Obstetrics and Gynecology (Urology and Gynecology) | S | Observation | Qualifier Value | SNOMED |
| 508 | Surgical Specialty | n/a | n/a | Peds Nephrology | n/a | wider | equal | does not map to specific site service line name | 4139904 | 305891006 | Seen by General Surgery Service (General Surgery) | S | Observation | Clinical Finding | SNOMED |
| 509 | Surgical Specialty | n/a | n/a | Family Practice | n/a | equal | equal | n/a | 4304711 | 419772000 | Family Practice (Other) | S | Observation | Qualifier Value | SNOMED |
| 510 | Surgical Specialty | n/a | n/a | Peds Pulmonary | n/a | wider | equal | does not map to specific site service line name | 4139904 | 305891006 | Seen by General Surgery Service (General Surgery) | S | Observation | Clinical Finding | SNOMED |
| 511 | Surgical Specialty | n/a | n/a | Peds Cardiology | n/a | wider | equal | does not map to specific site service line name | 37397651 | 718348005 | Cardiovascular Care (Cardiovascular) | S | Observation | Procedure | SNOMED |
| 512 | Surgical Specialty | n/a | n/a | Podiatry | n/a | equal | equal | n/a | 4149143 | 210087009 | Podiatry Service (Other) | S | Observation | Qualifier Value | SNOMED |
| 513 | Surgical Specialty | n/a | n/a | Peds Derm | n/a | equal | equal | n/a | 37207439 | 128001000 | Pediatric Dermatology Service (Other) | S | Observation | Qualifier Value | SNOMED |
| 514 | Surgical Specialty | n/a | n/a | Gastroenterology | n/a | wider | equal | does not map to specific site service line name | 4139904 | 305891006 | Seen by General Surgery Service (General Surgery) | S | Observation | Clinical Finding | SNOMED |
| 515 | Surgical Specialty | n/a | n/a | CVT | n/a | wider | equal | does not map to specific site service line name | 37397651 | 718348005 | Cardiovascular Care (Cardiovascular) | S | Observation | Procedure | SNOMED |
| 516 | Surgical Specialty | n/a | n/a | Colo-Rectal | n/a | wider | equal | does not map to specific site service line name | 4139904 | 305891006 | Seen by General Surgery Service (General Surgery) | S | Observation | Clinical Finding | SNOMED |
| 517 | Surgical Specialty | n/a | n/a | Hem Onc/BMT | n/a | wider | equal | does not map to specific site service line name | 1074156 | 1335967008 | Oncology Service (Oncology) | S | Observation | Qualifier Value | SNOMED |
| 518 | Surgical Specialty | n/a | n/a | FEMS (Bari/FG) | n/a | wider | equal | does not map to specific site service line name | 4139904 | 305891006 | Seen by General Surgery Service (General Surgery) | S | Observation | Clinical Finding | SNOMED |
| 519 | Surgical Specialty | n/a | n/a | Pulmonary | n/a | wider | equal | does not map to specific site service line name | 4139904 | 305891006 | Seen by General Surgery Service (General Surgery) | S | Observation | Clinical Finding | SNOMED |
| 520 | Surgical Specialty | n/a | n/a | Ortho Foot & Ankle | n/a | wider | equal | does not map to specific site service line name | 4150872 | 310161006 | Orthopedic Service (Ortho) | S | Observation | Qualifier Value | SNOMED |
| 521 | Surgical Specialty | n/a | n/a | Gynecologic Oncology | n/a | wider | equal | does not map to specific site service line name | 1074156 | 1335967008 | Oncology Service (Oncology) | S | Observation | Qualifier Value | SNOMED |
| 522 | Surgical Specialty | n/a | n/a | Ortho Hand | n/a | wider | equal | does not map to specific site service line name | 4150872 | 310161006 | Orthopedic Service (Ortho) | S | Observation | Qualifier Value | SNOMED |

| | | | | | | | | | | | | | | | | |
|---|---|---|---|---|---|---|---|---|---|---|---|---|---|---|---|---|
| 523 | Surgical Specialty | n/a | n/a | Ortho Joint | n/a | wider | equal | does not map to specific site service line name | 4150872 | 310161006 | Orthopedic Service (Ortho) | S | | Observation | Qualifier Value | SNOMED |
| 524 | Surgical Specialty | n/a | n/a | Neuro Spine | n/a | wider | equal | does not map to specific site service line name | 4138361 | 305894003 | Seen by Neurosurgery Service (Neurosurgery) | S | | Observation | Clinical Finding | SNOMED |
| 525 | Surgical Specialty | n/a | n/a | ENT | n/a | equal | equal | n/a | 4149155 | 310149003 | Ear, nose and throat service (HEENT) | S | | Observation | Qualifier Value | SNOMED |
| 526 | Surgical Specialty | n/a | n/a | Family Planning | n/a | wider | equal | does not map to specific site service line name | 4215122 | 394585009 | Obstetrics and Gynecology (Urology and Gynecology) | S | | Observation | Qualifier Value | SNOMED |
| 527 | Surgical Specialty | n/a | n/a | Ortho Oncology | n/a | wider | equal | does not map to specific site service line name | 1074156 | 1335967008 | Oncology Service (Oncology) | S | | Observation | Qualifier Value | SNOMED |
| 528 | Surgical Specialty | n/a | n/a | Ortho Spine | n/a | wider | equal | does not map to specific site service line name | 4150872 | 310161006 | Orthopedic Service (Ortho) | S | | Observation | Qualifier Value | SNOMED |
| 529 | Surgical Specialty | n/a | n/a | Peds Neurosurgery | n/a | wider | equal | does not map to specific site service line name | 4138361 | 305894003 | Seen by Neurosurgery Service (Neurosurgery) | S | | Observation | Clinical Finding | SNOMED |
| 530 | Surgical Specialty | n/a | n/a | Peds Orthopedics | n/a | wider | equal | does not map to specific site service line name | 4150872 | 310161006 | Orthopedic Service (Ortho) | S | | Observation | Qualifier Value | SNOMED |
| 531 | Surgical Specialty | n/a | n/a | Ortho Sports | n/a | wider | equal | does not map to specific site service line name | 4150872 | 310161006 | Orthopedic Service (Ortho) | S | | Observation | Qualifier Value | SNOMED |
| 532 | Surgical Specialty | n/a | n/a | Thoracic Surgery | n/a | wider | equal | does not map to specific site service line name | 4139904 | 305891006 | Seen by General Surgery Service (General Surgery) | S | | Observation | Clinical Finding | SNOMED |
| 533 | Surgical Specialty | n/a | n/a | Urogynecology | n/a | wider | equal | does not map to specific site service line name | 4215134 | 394612005 | Urology (Urology and Gynecology) | S | | Observation | Qualifier Value | SNOMED |
| 534 | Surgical Specialty | n/a | n/a | Cardiac Surgery | n/a | wider | equal | does not map to specific site service line name | 37397651 | 718348005 | Cardiovascular Care (Cardiovascular) | S | | Observation | Procedure | SNOMED |
| 535 | Surgical Specialty | n/a | n/a | Community Orthopedics | n/a | wider | equal | does not map to specific site service line name | 4150872 | 310161006 | Orthopedic Service (Ortho) | S | | Observation | Qualifier Value | SNOMED |
| 536 | Surgical Specialty | n/a | n/a | GEN SURG | n/a | equal | equal | n/a | 4139904 | 305891006 | Seen by General Surgery Service (General Surgery) | S | | Observation | Clinical Finding | SNOMED |
| 537 | Surgical Specialty | n/a | n/a | Neurosurgery | n/a | equal | equal | n/a | 4138361 | 305894003 | Seen by Neurosurgery Service (Neurosurgery) | S | | Observation | Clinical Finding | SNOMED |
| 538 | Surgical Specialty | n/a | n/a | NA | n/a | wider | equal | does not map to specific site service line name | 43273960 | 711385006 | Surgery inpatient care management (Other) | S | | Procedure | Procedure | SNOMED |

**Supplementary Table S4. Domain-Specific Mapping Performance**

**Purpose:** Detailed mapping statistics summarized by analytic domain.

| **STRICT** | Equal # | Equal % | Wider # | Wider % | Narrower # | Narrower % | Total |
|---|---|---|---|---|---|---|---|
| Admission Source | 6 | 30.0% | 11 | 55.0% | 3 | 15.0% | 20 |
| Admission Type | 6 | 100.0% | 0 | 0.0% | 0 | 0.0% | 6 |
| Anesthesia Class | 7 | 100.0% | 0 | 0.0% | 0 | 0.0% | 7 |
| Anesthesia Type | 1 | 100.0% | 0 | 0.0% | 0 | 0.0% | 1 |
| Drug Frequency | 0 | 0.0% | 2 | 100.0% | 0 | 0.0% | 2 |
| Drug Route | 3 | 100.0% | 0 | 0.0% | 0 | 0.0% | 3 |
| Ethnicity | 4 | 80.0% | 1 | 20.0% | 0 | 0.0% | 5 |
| Financial Class | 32 | 71.1% | 13 | 28.9% | 0 | 0.0% | 45 |
| Fluids I/O | 38 | 20.5% | 147 | 79.5% | 0 | 0.0% | 185 |
| Flowsheet | 12 | 92.3% | 1 | 7.7% | 0 | 0.0% | 13 |
| Medication | 36 | 54.5% | 29 | 43.9% | 1 | 1.5% | 66 |
| Other (drip v push) | 2 | 66.7% | 0 | 0.0% | 1 | 33.3% | 3 |
| Oxygen Device | 49 | 49.0% | 43 | 43.0% | 8 | 8.0% | 100 |
| Patient Type | 1 | 100.0% | 0 | 0.0% | 0 | 0.0% | 1 |
| Race | 6 | 23.1% | 20 | 76.9% | 0 | 0.0% | 26 |
| Surgery Type | 1 | 100.0% | 0 | 0.0% | 0 | 0.0% | 1 |
| Surgical Specialty | 14 | 26.4% | 39 | 73.6% | 0 | 0.0% | 53 |
| Tracheostomy | 1 | 100.0% | 0 | 0.0% | 0 | 0.0% | 1 |
| TOTAL | 219 | 40.7% | 306 | 58.0% | 13 | 2.0% | 538 |

| **LENIENT** | Equal # | Equal % | Wider # | Wider % | Narrower # | Narrower % | Total |
|---|---|---|---|---|---|---|---|
| Admission Source | 14 | 70.0% | 3 | 15.0% | 3 | 15.0% | 20 |
| Admission Type | 6 | 100.0% | 0 | 0.0% | 0 | 0.0% | 6 |
| Anesthesia Class | 7 | 100.0% | 0 | 0.0% | 0 | 0.0% | 7 |
| Anesthesia Type | 1 | 100.0% | 0 | 0.0% | 0 | 0.0% | 1 |
| Drug Frequency | 0 | 0.0% | 2 | 100.0% | 0 | 0.0% | 2 |
| Drug Route | 3 | 100.0% | 0 | 0.0% | 0 | 0.0% | 3 |
| Ethnicity | 5 | 100.0% | 0 | 0.0% | 0 | 0.0% | 5 |
| Financial Class | 45 | 100.0% | 0 | 0.0% | 0 | 0.0% | 45 |
| Fluids I/O | 181 | 97.8% | 4 | 2.2% | 0 | 0.0% | 185 |
| Flowsheet | 13 | 100.0% | 0 | 0.0% | 0 | 0.0% | 13 |
| Medication | 66 | 100.0% | 0 | 0.0% | 0 | 0.0% | 66 |
| Other (drip v push) | 3 | 100.0% | 0 | 0.0% | 0 | 0.0% | 3 |
| Oxygen Device | 98 | 98.0% | 2 | 2.0% | 0 | 0.0% | 100 |
| Patient Type | 1 | 100.0% | 0 | 0.0% | 0 | 0.0% | 1 |

| | | | | | | | |
|---|---|---|---|---|---|---|---|
| Race | 26 | 100.0% | 0 | 0.0% | 0 | 0.0% | 26 |
| Surgery Type | 1 | 100.0% | 0 | 0.0% | 0 | 0.0% | 1 |
| Surgical Specialty | 53 | 100.0% | 0 | 0.0% | 0 | 0.0% | 53 |
| Tracheostomy | 1 | 100.0% | 0 | 0.0% | 0 | 0.0% | 1 |
| TOTAL | 524 | 97.4% | 11 | 1.6% | 3 | 0.0% | 538 |

**Supplementary Table S5. Encounter-Level Validation of Analytic Concept Reconstruction Between Native Epic and OMOP Implementations**

| Analytic variable | Total, n (%) | PRF cases, n (%) | Controls, n (%) | Source/magnitude of difference | Interpretation for analytic fidelity |
|---|---|---|---|---|---|
| **Perioperative timeline** | | | | | |
| Hospital admission time | 0 (0.00%) | 0 (0.00%) | 0 (0.00%) | None | Complete agreement |
| Hospital discharge time | 0 (0.00%) | 0 (0.00%) | 0 (0.00%) | None | Complete agreement |
| Surgery date | 7 (0.02%) | 0 (0.00%) | 7 (0.02%) | Existing institutional OMOP ETL uses a different Epic source field. | No effect on cohort inclusion or derived analytic variables |
| OR wheels in time | 0 (0.00%) | 0 (0.00%) | 0 (0.00%) | None | Complete agreement |
| OR wheels out time | 0 (0.00%) | 0 (0.00%) | 0 (0.00%) | None | Complete agreement |
| Anesthesia start time | 0 (0.00%) | 0 (0.00%) | 0 (0.00%) | None | Complete agreement |
| Anesthesia end time | 0 (0.00%) | 0 (0.00%) | 0 (0.00%) | None | Complete agreement |
| Derived surgery start time | 0 (0.00%) | 0 (0.00%) | 0 (0.00%) | None | Complete agreement |
| Derived surgery end time | 0 (0.00%) | 0 (0.00%) | 0 (0.00%) | None | Complete agreement |
| Surgery start time | 0 (0.00%) | 0 (0.00%) | 0 (0.00%) | None | Complete agreement |
| Surgery end time | 0 (0.00%) | 0 (0.00%) | 0 (0.00%) | None | Complete agreement |
| **Outcome variables** | | | | | |
| Postoperative respiratory failure (case/control) | 0 (0.00%) | 0 (0.00%) | 0 (0.00%) | None | Complete agreement |
| Mechanical ventilation duration | 3 (0.01%) | 1 (0.38%) | 2 (0.01%) | SQL date-time precision; maximum difference 0.02 hours. | No meaningful effect on analytic value |
| **Perioperative fluid variables** | | | | | |

| Analytic variable | Total, n (%) | PRF cases, n (%) | Controls, n (%) | Source/magnitude of difference | Interpretation for analytic fidelity |
|---|---|---|---|---|---|
| OR fluid intake | 84 (0.27%) | 0 (0.00%) | 84 (0.27%) | Floating-point precision; maximum difference 0.001 L. | No meaningful effect on analytic value |
| OR fluid output | 1 (0.00%) | 0 (0.00%) | 1 (0.00%) | Floating-point precision; difference 0.001 L. | No meaningful effect on analytic value |
| OR net fluid balance | 79 (0.25%) | 0 (0.00%) | 79 (0.26%) | Propagated floating-point precision; maximum difference 0.001 L. | No meaningful effect on analytic value |
| Postoperative fluid intake | 452 (1.45%) | 1 (0.38%) | 451 (1.46%) | Floating-point precision; maximum difference 0.001 L. | No meaningful effect on analytic value |
| Postoperative fluid output | 23 (0.07%) | 0 (0.00%) | 23 (0.07%) | Floating-point precision; maximum difference 0.001 L. | No meaningful effect on analytic value |
| Postoperative net fluid balance | 501 (1.61%) | 1 (0.38%) | 500 (1.62%) | Propagated floating-point precision; maximum difference 0.001 L. | No meaningful effect on analytic value |
| **Medication exposure variables** | | | | | |
| Any vasopressor administration | 4 (0.01%) | 0 (0.00%) | 4 (0.01%) | Slightly different Epic and OMOP source data used to identify intravenous continuous infusion. | Difference in analytic classification; discrepant encounters reviewed |
| Continuous vasopressor infusion | 4 (0.01%) | 0 (0.00%) | 4 (0.01%) | Slightly different Epic and OMOP source data used to identify intravenous continuous infusion. | Structural/source-data representation difference affecting analytic classification |
| **Operative variables** | | | | | |
| Anesthesia duration | 0 (0.00%) | 0 (0.00%) | 0 (0.00%) | None | Complete agreement |
| Surgery duration | 0 (0.00%) | 0 (0.00%) | 0 (0.00%) | None | Complete agreement |
| ASA physical status | 0 (0.00%) | 0 (0.00%) | 0 (0.00%) | None | Complete agreement |
| Surgical specialty | 0 (0.00%) | 0 (0.00%) | 0 (0.00%) | Direct mapping according to predefined mapping specification. | Complete agreement |
| **Physiologic measurements** | | | | | |
| Median heart rate | 0 (0.00%) | 0 (0.00%) | 0 (0.00%) | None | Complete agreement |
| Median FiO2 | 0 (0.00%) | 0 (0.00%) | 0 (0.00%) | None | Complete agreement |

| Analytic variable | Total, n (%) | PRF cases, n (%) | Controls, n (%) | Source/magnitude of difference | Interpretation for analytic fidelity |
|---|---|---|---|---|---|
| Median tidal volume | 0 (0.00%) | 0 (0.00%) | 0 (0.00%) | None | Complete agreement |
| Median end-tidal CO2 | 654 (2.10%) | 8 (3.03%) | 646 (2.10%) | Floating-point precision; maximum difference 0.1%. | No meaningful effect on analytic value |
| **Demographic variables** | | | | | |
| Body mass index (continuous) | 1,555 (5.00%) | 20 (7.58%) | 1,535 (4.98%) | Different qualifying BMI, height, or weight measurements selected when multiple observations were available. | Measurement-selection difference; potential effect on continuous analytic value |
| BMI category | 81 (0.26%) | 6 (2.27%) | 75 (0.24%) | Differences propagated from selection of qualifying BMI, height, or weight measurements. | Difference in analytic classification; most differed by one BMI category |
| Medicare primary payer | 142 (0.46%) | 0 (0.00%) | 142 (0.46%) | Different payer selected when multiple overlapping payer records existed during hospitalization. | Structural difference in payer-selection hierarchy affecting analytic classification |
| Age | 0 (0.00%) | 0 (0.00%) | 0 (0.00%) | None | Complete agreement |
| Sex | 0 (0.00%) | 0 (0.00%) | 0 (0.00%) | Direct mapping according to predefined mapping specification. | Complete agreement |
| Race | 0 (0.00%) | 0 (0.00%) | 0 (0.00%) | Direct mapping according to predefined mapping specification. | Complete agreement |
| Ethnicity | 0 (0.00%) | 0 (0.00%) | 0 (0.00%) | Direct mapping according to predefined mapping specification. | Complete agreement |
| **Hospital utilization variables** | | | | | |
| Hospital length of stay | 1,289 (4.15%) | 11 (4.17%) | 1,278 (4.15%) | SQL date-time precision; maximum difference 0.01 days (~15 minutes). | No meaningful effect on analytic value |
| Postoperative day of diagnosis (POD_DX) | 2 (0.01%) | 2 (0.76%) | 0 (0.00%) | Differences in Oracle and Microsoft SQL Server date precision and rounding. | No meaningful effect on analytic classification |
| ICU length of stay | 250 (0.80%) | 73 (27.65%) | 177 (0.57%) | Mostly SQL date-time precision differences; maximum difference 0.04 days (~1 hour); 15 encounters | No meaningful effect on analytic value |

| Analytic variable | Total, n (%) | PRF cases, n (%) | Controls, n (%) | Source/magnitude of difference | Interpretation for analytic fidelity |
|---|---|---|---|---|---|
| | | | | differed by >0.01 days (~15 minutes). | |

Note: Variable-level concordance was evaluated among the 31,083 surgical encounters represented in both the native Epic and OMOP analytic cohorts, including 264 postoperative respiratory failure cases and 30,819 controls. Percentages are calculated using the corresponding population as the denominator (overall, N=31,083; cases, n=264; controls, n=30,819). A discrepancy was defined as any difference in the reconstructed analytic variable between implementations, regardless of magnitude. Discrepancy counts therefore include minor differences attributable to numerical precision, date-time precision or rounding, or selection among multiple qualifying source observations, even when these differences did not meaningfully alter the resulting analytic variable or classification. Sources of discrepancies and their potential effects on analytic representation are shown for each variable. Additional details regarding known sources of representation differences are provided in Supplementary Appendix B.

**Abbreviations:** ASA, American Society of Anesthesiologists; BMI, body mass index; FiO2, fraction of inspired oxygen; ICU, intensive care unit; LOS, length of stay; OMOP, Observational Medical Outcomes Partnership; OR, operating room; POD_DX, postoperative day of respiratory failure diagnosis; PRF, postoperative respiratory failure.

**Supplementary Figure S1**

**Operational ETL Architecture for Analytic Concept Reconstruction**

**Figure legend**
**Supplementary Figure S1. Operational ETL architecture used to transform native Epic EHR data into an analysis-ready OMOP dataset.** Unlike the conceptual framework presented in Figure 1, this operational workflow illustrates the functional implementation modules used during data processing. Structured source data extracted from Epic Clarity undergo vocabulary standardization before higher-order clinical representations are reconstructed using temporal logic, aggregation rules, and clinical decision rules. Clinical representations are subsequently transformed into derived analytic concepts that are integrated into the final analysis-ready dataset. Individual ETL modules were validated independently prior to final dataset assembly.

Clinical representation rules applied within this workflow are summarized in Supplementary Table S3.

**ALT TEXT**: Workflow showing transformation of native Epic Clarity EHR data through source extraction, OMOP vocabulary standardization, clinical representation, and analytic concept derivation to produce an analysis-ready OMOP dataset.

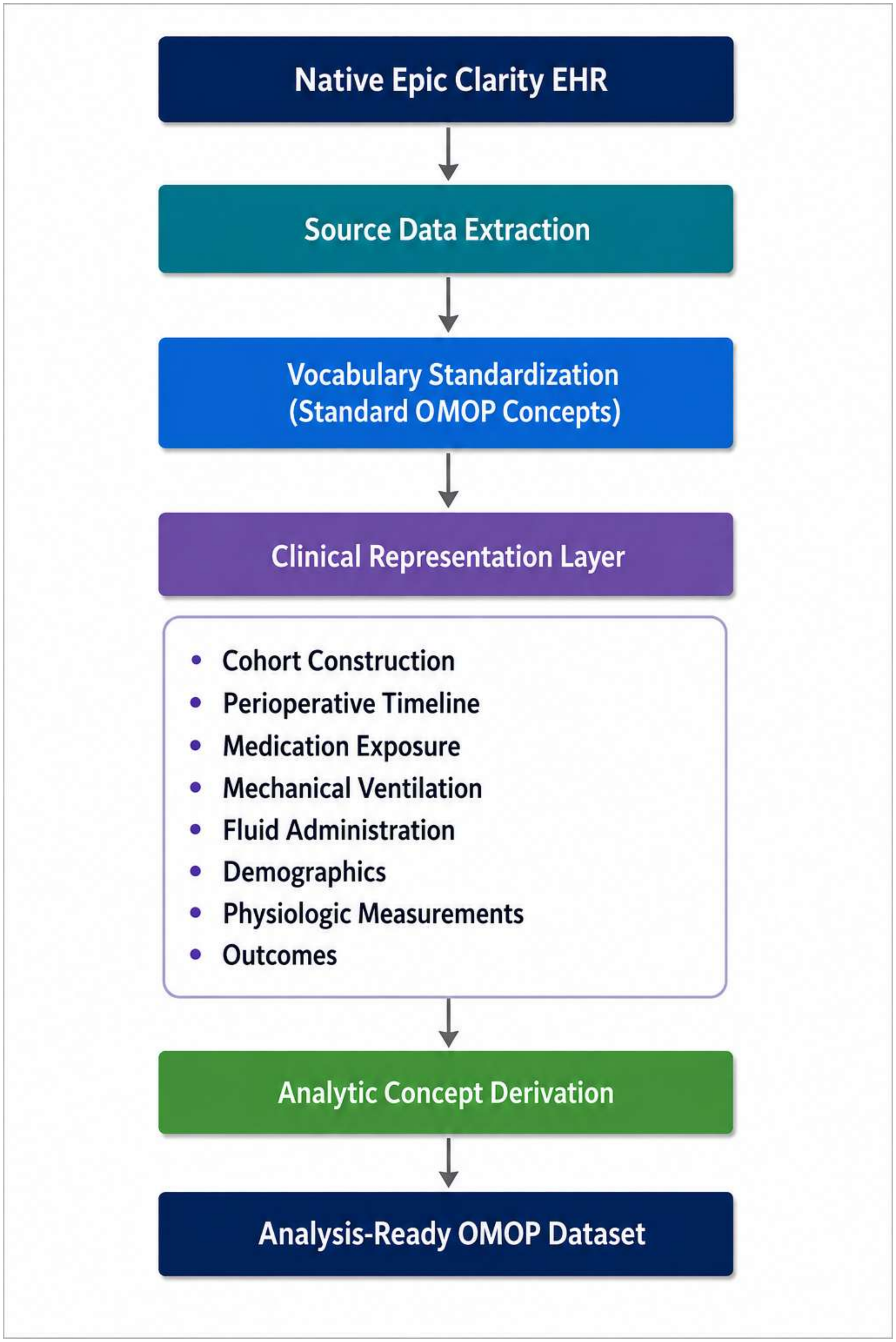
Native Epic Clarity EHR
Source Data Extraction
Vocabulary Standardization
(Standard OMOP Concepts)
Clinical Representation Layer
Cohort Construction
Perioperative Timeline
Medication Exposure
Mechanical Ventilation
Fluid Administration
Demographics
Physiologic Measurements
Outcomes
Analytic Concept Derivation
Analysis-Ready OMOP Dataset

**Supplementary Figure S2**

**Sequential Cohort Derivation from the OMOP Database**

**Figure legend**

**Supplementary Figure S2. Sequential derivation of the postoperative respiratory failure OMOP cohort.** Inclusion and exclusion criteria were applied sequentially to eligible surgical encounters. We excluded patients whose index operation involved a tracheostomy as part of a planned ENT procedure, which was evaluated at hospital admission and operating room exit. Upon record review, these patients had prolonged mechanical ventilation for airway protection, not for postoperative respiratory failure. Numbers within boxes indicate the remaining surgical encounters; numbers beside arrows indicate encounters excluded at each step.

Overall cohort concordance is presented in Figure 2 of the main manuscript.

**ALT TEXT:** Flowchart showing sequential application of eligibility criteria to 289,630 surgical encounters in the OMOP database, resulting in a final postoperative respiratory failure analysis cohort of 31,169 encounters.

### Flowchart of Cohort Derivation

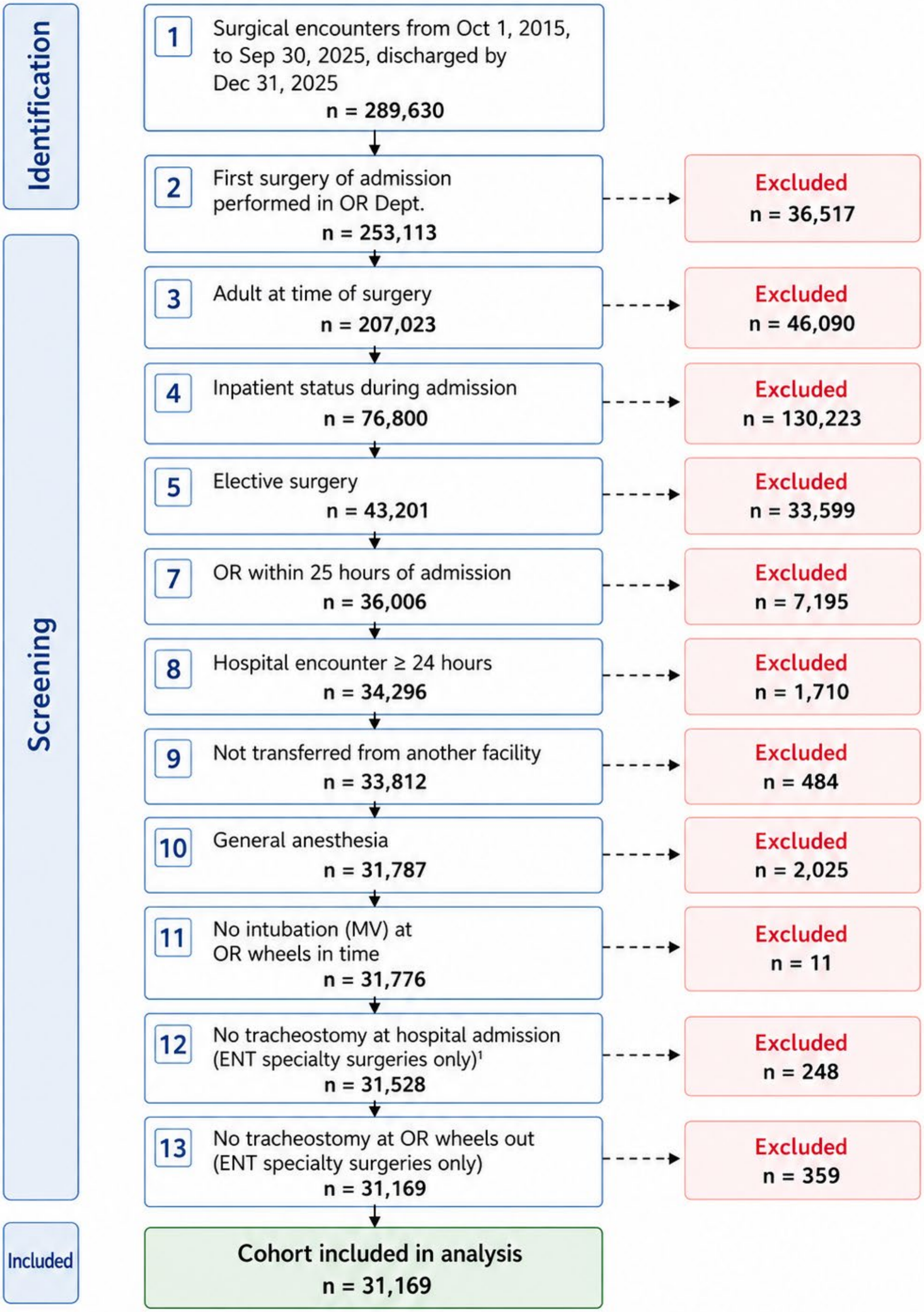


[1] ENT specialty surgeries only: ear, nose, and throat surgeries.

**Supplementary Figure S3**

**Admission Source Mapping and Sources of Cohort Discordance**

**Figure legend**

**Supplementary Figure S3. Conceptual representation of admission source mapping from Epic to OMOP.** Several granular Epic admission categories map to broader OMOP admission concepts. Loss of source granularity prevented differentiation of external versus internal emergency department transfers and accounted for most cohort discordance.

Known limitations of admission source representation are described in Supplementary Appendix B1.

**ALT TEXT**: Diagram showing granular Epic admission categories mapped to broader OMOP concepts, causing loss of source granularity, inability to distinguish internal versus external emergency transfers, and cohort discordance.

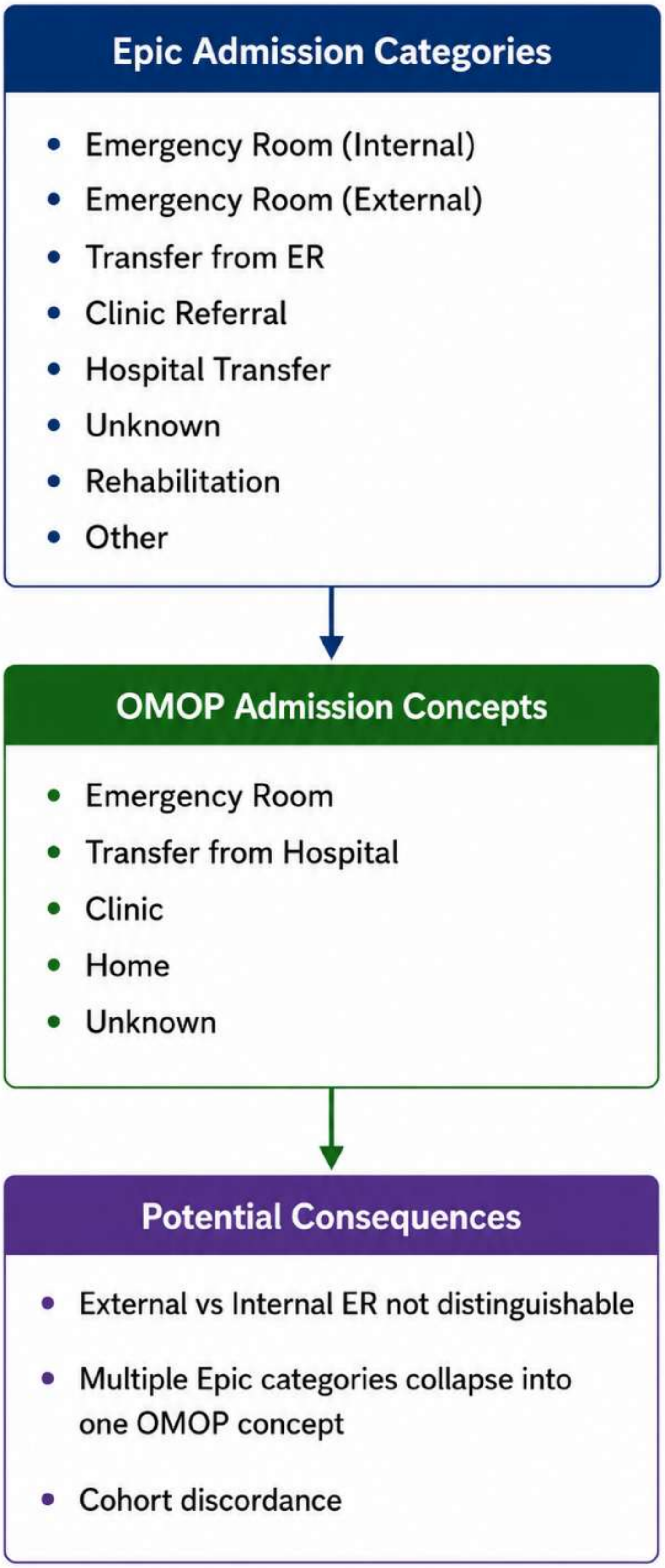
Epic Admission Categories
Emergency Room (Internal)
Emergency Room (External)
Transfer from ER
Clinic Referral
Hospital Transfer
Unknown
Rehabilitation
Other
OMOP Admission Concepts
Emergency Room
Transfer from Hospital
Clinic
Home
Unknown
Potential Consequences
External vs Internal ER not distinguishable
Multiple Epic categories collapse into one OMOP concept
Cohort discordance

**Supplementary Figure S4**

**Reconstruction of Continuous Vasopressor Infusion Episodes from Standard OMOP Concepts**

**Figure legend**

**Supplementary Figure S4. Reconstruction of continuous vasopressor infusion from standardized OMOP concepts.** Multiple standardized ingredient, route, and frequency concepts representing vasopressor infusion were combined using temporal logic to reconstruct clinically meaningful infusion episodes. These were subsequently aggregated to generate the analytic outcome used for continuous vasopressor infusion duration, however our model variable was binary (yes/no).

Detailed derivation rules are provided in Supplementary Appendix A1.

**ALT TEXT**: Flowchart showing standardized OMOP vasopressor concepts combined using classification and temporal logic to reconstruct continuous infusion episodes, calculate total infusion duration, and derive a binary continuous vasopressor variable.

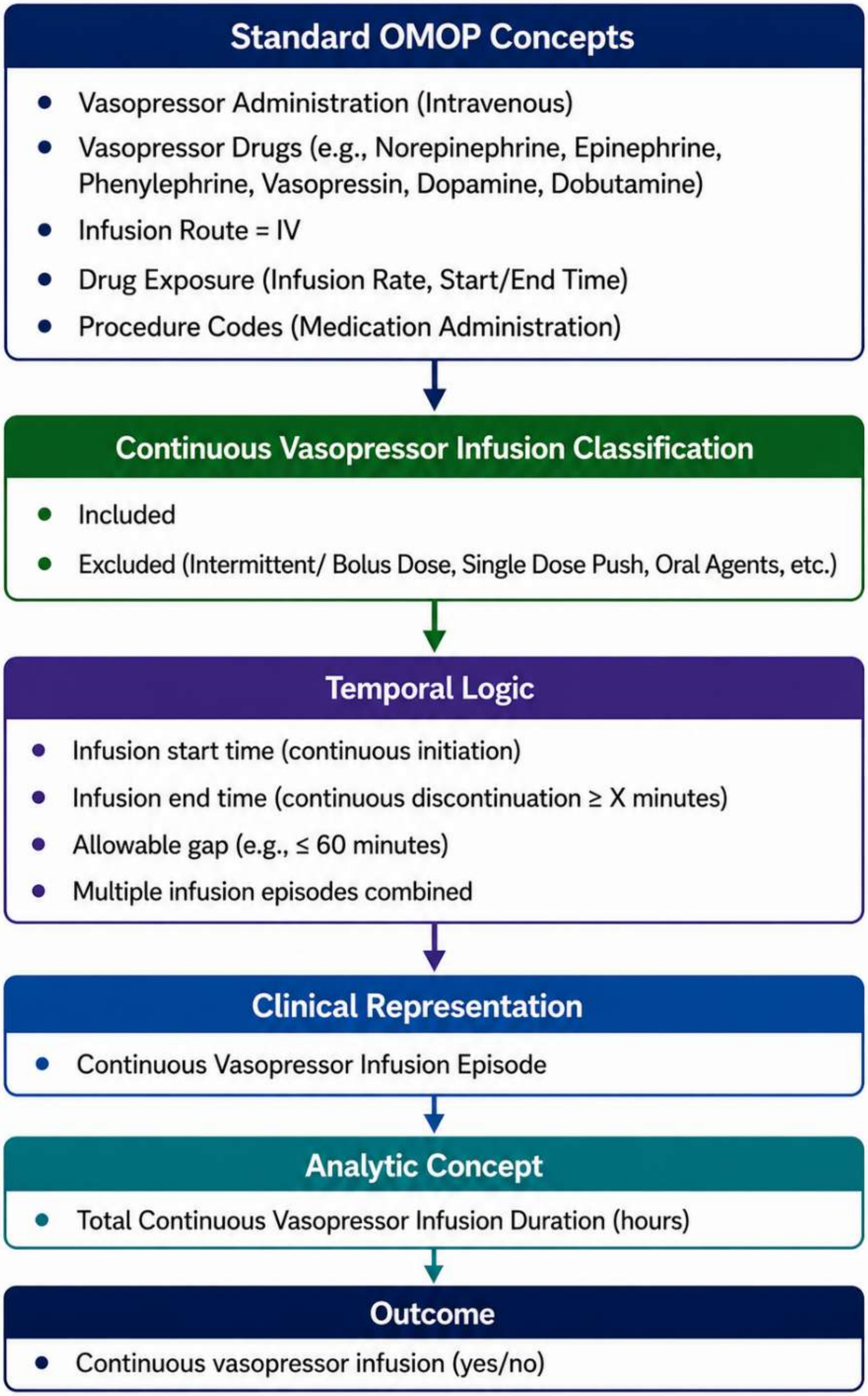
Standard OMOP Concepts
Vasopressor Administration (Intravenous)
Vasopressor Drugs (e.g., Norepinephrine, Epinephrine, Phenylephrine, Vasopressin, Dopamine, Dobutamine)
Infusion Route = IV
Drug Exposure (Infusion Rate, Start/End Time)
Procedure Codes (Medication Administration)
Continuous Vasopressor Infusion Classification
Included
Excluded (Intermittent/ Bolus Dose, Single Dose Push, Oral Agents, etc.)
Temporal Logic
Infusion start time (continuous initiation)
Infusion end time (continuous discontinuation ≥ X minutes)
Allowable gap (e.g., ≤ 60 minutes)
Multiple infusion episodes combined
Clinical Representation
Continuous Vasopressor Infusion Episode
Analytic Concept
Total Continuous Vasopressor Infusion Duration (hours)
Outcome
Continuous vasopressor infusion (yes/no)

**Supplementary Figure S5**

**Reconstruction of Mechanical Ventilation Episodes from Standard OMOP Concepts**

**Figure legend**

**Supplementary Figure S5. Reconstruction of postoperative mechanical ventilation from standardized OMOP concepts.** Multiple standardized device and procedure concepts representing invasive ventilation were combined using temporal logic to reconstruct clinically meaningful ventilation episodes. Episode durations were subsequently aggregated to generate the analytic outcome used for postoperative respiratory failure classification.

The standardized concepts used to reconstruct invasive mechanical ventilation episodes are listed in Supplementary Appendix C. Detailed derivation rules are provided in Supplementary Appendix A2.

**ALT TEXT**: Flowchart showing standardized OMOP ventilation concepts classified as invasive mechanical ventilation, combined using temporal logic into episodes, aggregated into total ventilation duration, and classified as postoperative respiratory failure at ≥48 hours.

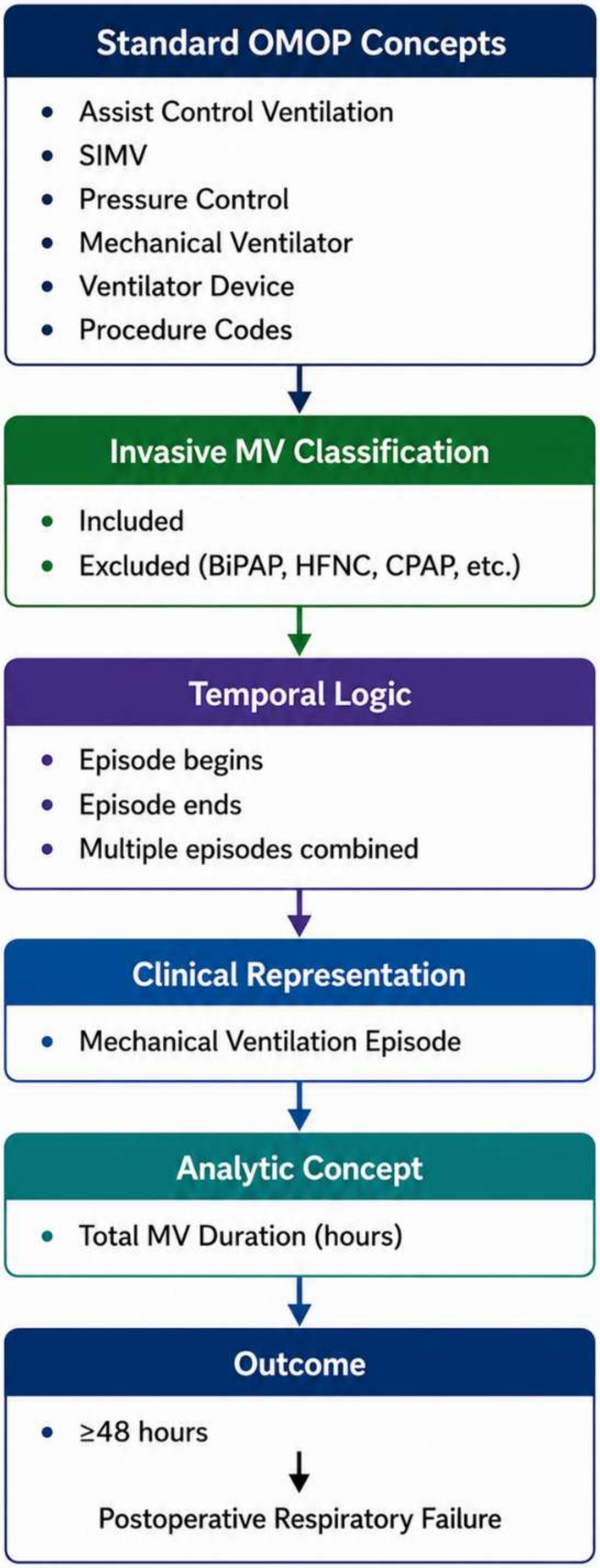
Standard OMOP Concepts
Assist Control Ventilation
SIMV
Pressure Control
Mechanical Ventilator
Ventilator Device
Procedure Codes
Invasive MV Classification
Included
Excluded (BiPAP, HFNC, CPAP, etc.)
Temporal Logic
Episode begins
Episode ends
Multiple episodes combined
Clinical Representation
Mechanical Ventilation Episode
Analytic Concept
Total MV Duration (hours)
Outcome
≥48 hours
Postoperative Respiratory Failure

**FIGURE LEGEND and ALT TEXT:**

**Supplementary Material**:

**Supplementary Figure S1**

**Operational ETL Architecture for Analytic Concept Reconstruction**

**Figure legend**
**Supplementary Figure S1. Operational ETL architecture used to transform native Epic EHR data into an analysis-ready OMOP dataset.** Unlike the conceptual framework presented in Figure 1, this operational workflow illustrates the functional implementation modules used during data processing. Structured source data extracted from Epic Clarity undergo vocabulary standardization before higher-order clinical representations are reconstructed using temporal logic, aggregation rules, and clinical decision rules. Clinical representations are subsequently transformed into derived analytic concepts that are integrated into the final analysis-ready dataset. Individual ETL modules were validated independently prior to final dataset assembly.

Clinical representation rules applied within this workflow are summarized in Supplementary Table S3.

**ALT TEXT**: Workflow showing transformation of native Epic Clarity EHR data through source extraction, OMOP vocabulary standardization, clinical representation, and analytic concept derivation to produce an analysis-ready OMOP dataset.

**Supplementary Figure S2**

**Sequential Cohort Derivation from the OMOP Database**

**Figure legend**
**Supplementary Figure S2. Sequential derivation of the postoperative respiratory failure OMOP cohort.** Inclusion and exclusion criteria were applied sequentially to eligible surgical encounters. We excluded patients whose index operation involved a tracheostomy as part of a planned ENT procedure, which was evaluated at hospital admission and operating room exit. Upon record review, these patients had prolonged mechanical ventilation for airway protection, not for postoperative respiratory failure. Numbers within boxes indicate the remaining surgical encounters; numbers beside arrows indicate encounters excluded at each step.

Overall cohort concordance is presented in Figure 2 of the main manuscript.

**ALT TEXT:** Flowchart showing sequential application of eligibility criteria to 289,630 surgical encounters in the OMOP database, resulting in a final postoperative respiratory failure analysis cohort of 31,169 encounters.

### Supplementary Figure S3

#### Admission Source Mapping and Sources of Cohort Discordance

**Figure legend**
**Supplementary Figure S3. Conceptual representation of admission source mapping from Epic to OMOP.** Several granular Epic admission categories map to broader OMOP admission concepts. Loss of source granularity prevented differentiation of external versus internal emergency department transfers and accounted for most cohort discordance.

Known limitations of admission source representation are described in Supplementary Appendix B1.
**ALT TEXT**: Diagram showing granular Epic admission categories mapped to broader OMOP concepts, causing loss of source granularity, inability to distinguish internal versus external emergency transfers, and cohort discordance.

### Supplementary Figure S4

#### Reconstruction of Continuous Vasopressor Infusion Episodes from Standard OMOP Concepts

**Figure legend**
**Supplementary Figure S4. Reconstruction of continuous vasopressor infusion from standardized OMOP concepts.** Multiple standardized ingredient, route, and frequency concepts representing vasopressor infusion were combined using temporal logic to reconstruct clinically meaningful infusion episodes. These were subsequently aggregated to generate the analytic outcome used for continuous vasopressor infusion duration, however our model variable was binary (yes/no).

Detailed derivation rules are provided in Supplementary Appendix A1.

**ALT TEXT**: Flowchart showing standardized OMOP vasopressor concepts combined using classification and temporal logic to reconstruct continuous infusion episodes, calculate total infusion duration, and derive a binary continuous vasopressor variable.

### Supplementary Figure S5

#### Reconstruction of Mechanical Ventilation Episodes from Standard OMOP Concepts

**Figure legend**
**Supplementary Figure S5. Reconstruction of postoperative mechanical ventilation from standardized OMOP concepts.** Multiple standardized device and procedure concepts representing invasive ventilation were combined using temporal logic to reconstruct clinically meaningful ventilation episodes. Episode durations were subsequently aggregated to generate the analytic outcome used for postoperative respiratory failure classification.

The standardized concepts used to reconstruct invasive mechanical ventilation episodes are listed in Supplementary Appendix C. Detailed derivation rules are provided in Supplementary Appendix A2.

**ALT TEXT**: Flowchart showing standardized OMOP ventilation concepts classified as invasive mechanical ventilation, combined using temporal logic into episodes, aggregated into total ventilation duration, and classified as postoperative respiratory failure at ≥48 hours.

**Supplementary Appendix A**

**Detailed Analytic Concept Definitions**

**Overview**
This appendix documents the analytic concept specifications used to reconstruct the postoperative respiratory failure (PRF) analytic dataset from standardized OMOP concepts. Unlike the source concept mappings presented in Supplementary Table S1, these specifications describe how standardized concepts were combined using reproducible clinical representation logic to generate the analysis-ready variables used in this study.

Each analytic concept is described using a standardized template consisting of:

- Clinical intent
- Source concepts
- Standard OMOP concepts
- Clinical representation logic
- Derived analytic concept
- Validation performed
- Known limitations

These specifications are implementation-independent and are intended to facilitate reproducible analytic concept development across institutions.

**A1. Continuous Vasopressor Infusion**

**Clinical Intent**
Identify patients who received continuous intravenous vasopressor therapy during the intraoperative period.

**Source Concepts**

- Medication administration records
- Medication orders
- Drug administration timestamps
- Administration route
- Dose units

**Standard OMOP Concepts**

- Standard RxNorm ingredient concepts representing vasopressor medications
- Administration route concepts
- Drug exposure timestamps

**Clinical Representation Logic**
Individual vasopressor drug exposures were first standardized to OMOP ingredient concepts. Continuous intravenous administration was then reconstructed using administration characteristics rather than medication formulation.
A qualifying exposure required:

- vasopressor ingredient,
- intravenous administration,
- evidence of continuous administration,
- administration during the anesthesia interval.

Continuous administration was inferred from dose-unit terminology containing a temporal component (for example mcg/min or mcg/kg/min) when explicit infusion frequency was unavailable.

**Derived Analytic Concept**
Binary indicator:
Continuous intraoperative vasopressor infusion (Yes/No)

**Validation Performed**
Compared with the native Epic implementation among all overlapping cohort encounters. Agreement exceeded 99%. Discordant records underwent manual review.

**Known Limitations**
The OMOP CDM does not explicitly represent medication administration frequency within the Drug Exposure table. Continuous infusion therefore required inference from dose-unit terminology.

**A2. Mechanical Ventilation Duration**

**Clinical Intent**

Determine cumulative postoperative invasive mechanical ventilation duration.

**Source Concepts**
- Oxygen device documentation
- Ventilator documentation
- Respiratory procedure records

**Standard OMOP Concepts**
Standard procedure concepts representing invasive ventilation
Standard device concepts representing ventilators

**Clinical Representation Logic**
Individual device and procedure concepts representing invasive mechanical ventilation were classified according to predefined inclusion criteria. Qualifying concepts were linked using temporal overlap to reconstruct ventilation episodes. Non-invasive respiratory support (BiPAP, CPAP, HFNC, oxygen mask, nasal cannula) was excluded. Multiple episodes were combined to calculate cumulative postoperative duration.

**Derived Analytic Concept**
Mechanical ventilation duration (hours)

**Validation Performed**
Compared against the Epic-derived duration. Maximum observed difference was clinically negligible.

**Known Limitations**
Dependent upon accurate documentation of ventilation start and stop times. Complete list of concepts appears in Appendix C.

### A3. Postoperative Respiratory Failure

**Clinical Intent**
Classify postoperative respiratory failure according to the study phenotype.

**Source Concepts**
Mechanical ventilation episodes

**Standard OMOP Concepts**
Derived mechanical ventilation episode

**Clinical Representation Logic**
Patients accumulating at least 48 hours of invasive postoperative mechanical ventilation were classified as cases. Remaining patients were classified as controls.

**Derived Analytic Concept**
Binary PRF outcome

**Validation**
Compared directly against the Epic implementation. Complete agreement.

### A4. Perioperative Fluid Balance

**Clinical Intent**
Estimate fluid administration during predefined perioperative periods.

**Source Concepts**
- Intake documentation
- Output documentation

**Standard OMOP Concepts**
Measurement concepts
Observation concepts

**Clinical Representation Logic**
Fluid intake and output were independently aggregated over predefined perioperative windows.

Net balance was calculated as: Fluid Intake − Fluid Output
Separate variables were generated for
- intraoperative interval
- postoperative 24-hour interval

**Derived Analytic Concepts**
- OR fluid intake
- OR fluid output
- OR net fluid balance
- Postoperative intake
- Postoperative output

- Postoperative net fluid balance

**Validation**
Compared numerically against Epic. Observed differences reflected floating-point precision only.

**A5. Perioperative Physiologic Measurements**

**Clinical Intent**
To summarize intraoperative physiologic status using standardized measures that characterize the patient's physiologic condition during general anesthesia. Rather than relying on individual observations, representative summary measures were generated for each physiologic variable over the intraoperative period.

**Source Concepts**
Source physiologic data were extracted from structured anesthesia documentation and perioperative flowsheet records within the native Epic EHR. Variables included heart rate, inspired oxygen fraction ($FiO_2$), tidal volume, and end-tidal carbon dioxide ($EtCO_2$).

**Standard OMOP Concepts**
Physiologic observations were mapped to standard OMOP Measurement concepts representing:

- Heart rate
- Fraction of inspired oxygen ($FiO_2$)
- Tidal volume
- End-tidal carbon dioxide ($EtCO_2$)

All measurements retained their original timestamps following standardization.

**Clinical Representation Logic**
Only physiologic measurements occurring during the intraoperative period were considered for analysis. The intraoperative interval was defined using the anesthesia start and anesthesia end times. Measurements outside this interval were excluded. Multiple observations (minute-by-minute) were available and all qualifying values were retained for summary calculation.

**Aggregation Method**
For each physiologic variable, the median value of all qualifying intraoperative measurements was calculated. Median values were selected because they provide a robust estimate of central tendency while minimizing the influence of transient physiologic artifacts, monitoring noise, and isolated outlier measurements that commonly occur during anesthesia.

**Temporal Window**
**Start:** Anesthesia start time

**End:** Anesthesia end time
Only observations whose timestamps occurred within this interval contributed to the analytic variable.

**Derived Analytic Variables**
The following analytic variables were generated:

- Median heart rate (beats/minute)
- Median $FiO_2$ (%)

- Median tidal volume (mL)
- Median end-tidal carbon dioxide (mmHg)

**Validation Performed**
Derived variables were compared with the corresponding native Epic implementation for all encounters represented in both datasets.
Complete agreement was observed for median heart rate, $FiO_2$, and tidal volume. Minor numeric differences were observed for end-tidal carbon dioxide and reflected floating-point precision rather than clinically meaningful differences.

**Known Limitations**
Because physiologic measurements were summarized rather than represented individually, transient physiologic events were not explicitly preserved within the analytic dataset. This approach was appropriate for the predictive modeling application but may not be suitable for investigations requiring high-frequency physiologic time-series analyses.

**A6. Body Mass Index**

**Clinical Intent**
To estimate preoperative body habitus using the most clinically appropriate body mass index (BMI) available before surgery.

**Source Concepts**
Source data included the following structured measurements recorded before surgery within the native EHR.

- Body mass index
- Height
- Weight

**Standard OMOP Concepts**
Standard OMOP **Measurement** concepts representing:

- Body mass index
- Body height
- Body weight

**Clinical Representation Logic**
BMI values were determined using the following hierarchical selection strategy.

**Selection Hierarchy**
**Step 1:** When a qualifying BMI measurement was available before anesthesia, the measurement recorded closest to anesthesia start was selected. **Step 2:** If no qualifying BMI measurement existed, the nearest qualifying height and weight measurements recorded before anesthesia were identified. **Step 3:** BMI was calculated using:

$$BMI = \frac{\text{Weight (kg)}}{\text{Height (m)}^2}$$

**Temporal Window**

Measurements were eligible only if recorded before anesthesia start time. When multiple qualifying observations were present, the measurement closest to anesthesia start was preferentially selected.

**Derived Analytic Variables**

- Continuous BMI ($kg/m^2$)
- BMI category

**Validation Performed**

BMI values were compared between the native Epic and reconstructed OMOP datasets. Most differences resulted from selection of different qualifying measurements when multiple height, weight, or BMI observations were available before surgery. Categorical BMI agreement remained high, with most discrepancies differing by only a single BMI category.

**Known Limitations**

The OMOP Common Data Model does not preserve the exact measurement selection logic used within the native Epic implementation. Consequently, encounters containing multiple qualifying height, weight, or BMI observations occasionally resulted in different measurement selection despite identical derivation rules. These differences reflected alternative—but clinically reasonable—measurement selection rather than failures of concept mapping or analytic reconstruction.

**A7. Medicare Primary Payer**

**Clinical Intent**

To determine whether Medicare represented the patient's primary payer during the operative hospitalization.

**Source Concepts**

Administrative payer records associated with the hospitalization.

**Standard OMOP Concepts**

OMOP Payer Plan Period records linked to the patient.

**Clinical Representation Logic**

Payer records whose coverage periods overlapped the operative hospitalization were identified. When multiple qualifying payer records existed, a single payer was selected according to a predefined hierarchy designed to approximate the payer associated with surgery.

**Selection Hierarchy**

1. Identify all payer plan periods overlapping the hospitalization.
2. When only one qualifying payer existed, assign that payer.
3. When multiple qualifying payer records existed, select the payer plan period whose start or end date occurred closest to hospital admission.
4. Determine whether the selected payer represented Medicare.

**Temporal Window**

Coverage periods overlapping the hospitalization.

**Derived Analytic Variable**

Medicare primary payer (Yes/No)

**Validation Performed**
The selected payer was compared with the native Epic implementation. Agreement exceeded 99%. Nearly all discordant records occurred among patients with multiple overlapping payer records.

**Known Limitations**
Neither the OMOP CDM nor the native Epic implementation provides a universally accepted method for selecting a primary payer when multiple concurrent payer records exist. Consequently, small differences reflected alternative payer selection rather than differences in standardized vocabulary mapping.

### A8. Cohort Construction

**Clinical Intent**
To reproduce the reference postoperative respiratory failure cohort within the OMOP Common Data Model while preserving the clinical intent of the original Epic implementation.

**Clinical Representation Logic**
Rather than identifying isolated encounters, cohort construction reconstructed the clinical eligibility criteria used in the original study by integrating standardized encounter, procedure, observation, condition, and demographic concepts. All inclusion and exclusion criteria were evaluated sequentially.

**Inclusion Criteria**
Eligible encounters satisfied all of the following conditions:

- Adult patients (≥18 years at surgery)
- Elective surgery
- First operative procedure during the hospitalization
- Surgery performed in an operating room
- General anesthesia
- Inpatient hospitalization
- Operating room entry within 25 hours of hospital admission
- Hospital length of stay of at least 24 hours
- Surgery performed during the study period
- Hospital discharge before the administrative study end date

**Exclusion Criteria**
Encounters were excluded if they met any of the following criteria:

- Transfer from another acute care facility
- Mechanical ventilation present at operating room entry
- Pre-existing (upon admission) tracheostomy among ENT specialty surgical encounters
- Tracheostomy present at operating room exit among ENT specialty surgical encounters

Tracheostomy variables were retained within the analytic dataset and were used to support flexible cohort definition.

**Derived Analytic Concept**
Binary cohort inclusion indicator: (Eligible / Not Eligible)

**Validation Performed**
The reconstructed OMOP cohort was compared directly with the native Epic cohort. Overall cohort concordance exceeded 99%. Most discordant encounters resulted from:

- institutional ETL requirements excluding surgical encounters lacking qualifying CPT codes,
- structural differences in admission source representation,
- isolated discrepancies in inpatient status documentation.

**Known Limitations**
Several cohort eligibility criteria required approximation because of limitations in the current OMOP Common Data Model. Most notably, standardized admission source concepts could not distinguish transfers originating from external versus internal emergency departments. In addition, institutional OMOP ETL processes excluded certain surgical encounters lacking qualifying CPT codes before cohort construction. Despite these limitations, reconstructed cohort membership demonstrated excellent agreement with the native Epic implementation, supporting the validity of the analytic interoperability framework.

### Supplementary Appendix B

### Known Representation Limitations

#### Overview
Despite high implementation fidelity, several analytic concepts required approximation because of limitations in either the current OMOP vocabulary or the underlying OMOP Common Data Model. These limitations did not substantially affect analytic fidelity but should be considered during implementation at other institutions.

#### B1. Admission Source

**Limitation**
OMOP admission source concepts do not distinguish transfers from internal and external emergency departments.

**Consequence**
Small cohort differences.

**Potential future solution**
Expanded admission source vocabulary.

#### B2. Continuous Infusion Frequency

**Limitation**
Drug Exposure contains: Drug, Dose, Route, but not explicit administration frequency.

**Consequence**
Continuous infusions inferred from dose-unit terminology.

**Future enhancement**
Dedicated infusion frequency attribute.

#### B3. CPT-dependent Surgical Encounter Identification

**Limitation**
Institutional OMOP ETL required valid CPT codes before surgical encounters were transferred.

**Consequence**
Some otherwise eligible surgical encounters absent from OMOP.

**Future enhancement**
Encounter-level surgical identifiers independent of CPT.

#### B4. Body Mass Index Selection

**Limitation**
Multiple qualifying height and weight observations may exist.

**Consequence**
Epic and OMOP occasionally selected different measurements.

**Effect**
Minimal. Mostly <1 BMI categorical unit.

**B5. Primary Payer Hierarchy**

**Limitation**
Multiple payer records frequently overlap.

**Consequence**
Epic and OMOP occasionally selected different primary payers.

**Future enhancement**
Standardized payer prioritization algorithm.

**Supplementary Appendix C**

**Appendix C. Standard OMOP Concepts Used for Mechanical Ventilation Reconstruction**

| Standard OMOP Concept ID | Standard OMOP Concept Name | Vocabulary | Included as Invasive Mechanical Ventilation | Notes |
|---|---|---|---|---|
| 4055377 | High frequency jet ventilation | SNOMED | ✓ | Included as invasive mechanical ventilation |
| 37152363 | Synchronized intermittent mandatory ventilation – volume control pressure-support inflation | SNOMED | ✓ | Included as invasive mechanical ventilation |
| 37152364 | Synchronized intermittent mandatory ventilation – pressure-control pressure-support inflation | SNOMED | ✓ | Included as invasive mechanical ventilation |
| 37152384 | Synchronized intermittent mandatory ventilation – volume targeted pressure-control and pressure-support inflation | SNOMED | ✓ | Included as invasive mechanical ventilation |
| 37152388 | Synchronized intermittent mandatory ventilation with pressure-control, an assured constant airway pressure adjunct and pressure-support at the higher pressure level only | SNOMED | ✓ | Included as invasive mechanical ventilation |
| 37152409 | Assist control ventilation volume-control inflation-type | SNOMED | ✓ | Included as invasive mechanical ventilation |
| 37152410 | Assist control ventilation pressure control inflation-type | SNOMED | ✓ | Included as invasive mechanical ventilation |
| 37152411 | Assist control ventilation volume-targeted pressure-control inflation-type | SNOMED | ✓ | Included as invasive mechanical ventilation |
| 37154096 | Pressure support ventilation (regime/therapy) | SNOMED | ✓ | Included as invasive mechanical ventilation |

| | | | | |
|---|---|---|---|---|
| 37158404 | Invasive mechanical ventilation | SNOMED | ✓ | Included as invasive mechanical ventilation |
| 40493026 | Mechanical ventilator | SNOMED | ✓ | Included as invasive mechanical ventilation |
| 45768197 | Ventilator | SNOMED | ✓ | Included as invasive mechanical ventilation |
| 605914 | Continuous positive airway pressure bilevel positive airway pressure face mask (physical object) | SNOMED | ✗ | Non-invasive respiratory support |
| 4070683 | Oxygen tent | SNOMED | ✗ | Supplemental oxygen only |
| 4138614 | BiPAP oxygen nasal cannula | SNOMED | ✗ | Non-invasive respiratory support |
| 4139525 | High flow oxygen nasal cannula (physical object) | SNOMED | ✗ | High-flow oxygen therapy; non-invasive |
| 4143247 | Bag valve mask ventilation | SNOMED | ✗ | Temporary manual ventilation; not considered invasive mechanical ventilation episode |
| 4145694 | Aerosol oxygen mask | SNOMED | ✗ | Supplemental oxygen only |
| 4155151 | Oxygen administration by nasal cannula (procedure) | SNOMED | ✗ | Supplemental oxygen only |
| 4162736 | Oxygen administration by mask (procedure) | SNOMED | ✗ | Supplemental oxygen only |
| 4165535 | Continuous positive airway pressure ventilation treatment | SNOMED | ✗ | Non-invasive respiratory support |
| 4188569 | T piece with bag | SNOMED | ✗ | Airway adjunct; not classified as invasive mechanical ventilation |
| 4188570 | T piece without bag | SNOMED | ✗ | Airway adjunct; not classified as invasive mechanical ventilation |
| 4323763 | Oxygen administration by Venturi mask (procedure) | SNOMED | ✗ | Supplemental oxygen only |
| 4327886 | Dependence on biphasic positive airway pressure (BiPAP) ventilation | SNOMED | ✗ | Non-invasive respiratory support |

| | | | | |
|---|---|---|---|---|
| 36305294 | LCDS v4.00 – Spontaneous breathing trial during assessment period [CMS Assessment] | SNOMED | ✗ | Assessment concept only |
| 36308581 | Supplemental oxygen | SNOMED | ✗ | Supplemental oxygen only |
| 36716647 | Breathing room air (finding) | SNOMED | ✗ | Indicates absence of oxygen therapy |
| 40486624 | Non-invasive positive pressure ventilation (regime/therapy) | SNOMED | ✗ | Non-invasive respiratory support |
| 40488354 | CPAP (continuous positive airway pressure) titration | SNOMED | ✗ | CPAP assessment/titration |
| 44790731 | Oxygen administration by non-rebreather mask (procedure) | SNOMED | ✗ | Supplemental oxygen only |
| 45759373 | Oxygen administration face tent | SNOMED | ✗ | Supplemental oxygen only |
| 45759376 | Oxygen administration hood, adult | SNOMED | ✗ | Supplemental oxygen only |
| 45761494 | CPAP nasal oxygen cannula | SNOMED | ✗ | Non-invasive respiratory support |

**Abbreviations:** BiPAP, bilevel positive airway pressure; CPAP, continuous positive airway pressure; SNOMED, Systematized Nomenclature of Medicine Clinical Terms.

**Footnote:** Standard OMOP concepts representing invasive mechanical ventilation were used to reconstruct postoperative mechanical ventilation episodes through temporal aggregation, as described in Supplementary Method 3. Concepts representing supplemental oxygen therapy, non-invasive ventilation, respiratory assessments, or airway adjuncts were intentionally excluded from episode reconstruction to preserve the clinical definition of invasive mechanical ventilation used for postoperative respiratory failure phenotyping.

**FIGURE LEGEND and ALT TEXT:**

**Supplementary Material**:

**Supplementary Figure S1**

**Operational ETL Architecture for Analytic Concept Reconstruction**

**Figure legend**
**Supplementary Figure S1. Operational ETL architecture used to transform native Epic EHR data into an analysis-ready OMOP dataset.** Unlike the conceptual framework presented in Figure 1, this operational workflow illustrates the functional implementation modules used during data processing. Structured source data extracted from Epic Clarity undergo vocabulary standardization before higher-order clinical representations are reconstructed using temporal logic, aggregation rules, and clinical decision rules. Clinical representations are subsequently transformed into derived analytic concepts that are integrated into the final analysis-ready dataset. Individual ETL modules were validated independently prior to final dataset assembly.

Clinical representation rules applied within this workflow are summarized in Supplementary Table S3.

**ALT TEXT**: Workflow showing transformation of native Epic Clarity EHR data through source extraction, OMOP vocabulary standardization, clinical representation, and analytic concept derivation to produce an analysis-ready OMOP dataset.

**Supplementary Figure S2**

**Sequential Cohort Derivation from the OMOP Database**

**Figure legend**
**Supplementary Figure S2. Sequential derivation of the postoperative respiratory failure OMOP cohort.** Inclusion and exclusion criteria were applied sequentially to eligible surgical encounters. We excluded patients whose index operation involved a tracheostomy as part of a planned ENT procedure, which was evaluated at hospital admission and operating room exit. Upon record review, these patients had prolonged mechanical ventilation for airway protection, not for postoperative respiratory failure. Numbers within boxes indicate the remaining surgical encounters; numbers beside arrows indicate encounters excluded at each step.

Overall cohort concordance is presented in Figure 2 of the main manuscript.

**ALT TEXT:** Flowchart showing sequential application of eligibility criteria to 289,630 surgical encounters in the OMOP database, resulting in a final postoperative respiratory failure analysis cohort of 31,169 encounters.

**Supplementary Figure S3**

**Admission Source Mapping and Sources of Cohort Discordance**

**Figure legend**
**Supplementary Figure S3. Conceptual representation of admission source mapping from Epic to OMOP.** Several granular Epic admission categories map to broader OMOP admission concepts. Loss of source granularity prevented differentiation of external versus internal emergency department transfers and accounted for most cohort discordance.

Known limitations of admission source representation are described in Supplementary Appendix B1.
**ALT TEXT**: Diagram showing granular Epic admission categories mapped to broader OMOP concepts, causing loss of source granularity, inability to distinguish internal versus external emergency transfers, and cohort discordance.

**Supplementary Figure S4**

**Reconstruction of Continuous Vasopressor Infusion Episodes from Standard OMOP Concepts**

**Figure legend**
**Supplementary Figure S4. Reconstruction of continuous vasopressor infusion from standardized OMOP concepts.** Multiple standardized ingredient, route, and frequency concepts representing vasopressor infusion were combined using temporal logic to reconstruct clinically meaningful infusion episodes. These were subsequently aggregated to generate the analytic outcome used for continuous vasopressor infusion duration, however our model variable was binary (yes/no).

Detailed derivation rules are provided in Supplementary Appendix A1.

**ALT TEXT**: Flowchart showing standardized OMOP vasopressor concepts combined using classification and temporal logic to reconstruct continuous infusion episodes, calculate total infusion duration, and derive a binary continuous vasopressor variable.

**Supplementary Figure S5**

**Reconstruction of Mechanical Ventilation Episodes from Standard OMOP Concepts**

**Figure legend**
**Supplementary Figure S5. Reconstruction of postoperative mechanical ventilation from standardized OMOP concepts.** Multiple standardized device and procedure concepts representing invasive ventilation were combined using temporal logic to reconstruct clinically meaningful ventilation episodes. Episode durations were subsequently aggregated to generate the analytic outcome used for postoperative respiratory failure classification.

The standardized concepts used to reconstruct invasive mechanical ventilation episodes are listed in Supplementary Appendix C. Detailed derivation rules are provided in Supplementary Appendix A2.

**ALT TEXT**: Flowchart showing standardized OMOP ventilation concepts classified as invasive mechanical ventilation, combined using temporal logic into episodes, aggregated into total ventilation duration, and classified as postoperative respiratory failure at ≥48 hours.

**FIGURE LEGEND and ALT TEXT:**

**Supplementary Material**:

**Supplementary Figure S1**

**Operational ETL Architecture for Analytic Concept Reconstruction**

**Figure legend**

**Supplementary Figure S1. Operational ETL architecture used to transform native Epic EHR data into an analysis-ready OMOP dataset.** Unlike the conceptual framework presented in Figure 1, this operational workflow illustrates the functional implementation modules used during data processing. Structured source data extracted from Epic Clarity undergo vocabulary standardization before higher-order clinical representations are reconstructed using temporal logic, aggregation rules, and clinical decision rules. Clinical representations are subsequently transformed into derived analytic concepts that are integrated into the final analysis-ready dataset. Individual ETL modules were validated independently prior to final dataset assembly.

Clinical representation rules applied within this workflow are summarized in Supplementary Table S3.

**ALT TEXT**: Workflow showing transformation of native Epic Clarity EHR data through source extraction, OMOP vocabulary standardization, clinical representation, and analytic concept derivation to produce an analysis-ready OMOP dataset.

**Supplementary Figure S2**

**Sequential Cohort Derivation from the OMOP Database**

**Figure legend**

**Supplementary Figure S2. Sequential derivation of the postoperative respiratory failure OMOP cohort.** Inclusion and exclusion criteria were applied sequentially to eligible surgical encounters. We excluded patients whose index operation involved a tracheostomy as part of a planned ENT procedure, which was evaluated at hospital admission and operating room exit. Upon record review, these patients had prolonged mechanical ventilation for airway protection, not for postoperative respiratory failure. Numbers within boxes indicate the remaining surgical encounters; numbers beside arrows indicate encounters excluded at each step.

Overall cohort concordance is presented in Figure 2 of the main manuscript.

**ALT TEXT:** Flowchart showing sequential application of eligibility criteria to 289,630 surgical encounters in the OMOP database, resulting in a final postoperative respiratory failure analysis cohort of 31,169 encounters.

**Supplementary Figure S3**

**Admission Source Mapping and Sources of Cohort Discordance**

**Figure legend**
**Supplementary Figure S3. Conceptual representation of admission source mapping from Epic to OMOP.** Several granular Epic admission categories map to broader OMOP admission concepts. Loss of source granularity prevented differentiation of external versus internal emergency department transfers and accounted for most cohort discordance.

Known limitations of admission source representation are described in Supplementary Appendix B1.
**ALT TEXT**: Diagram showing granular Epic admission categories mapped to broader OMOP concepts, causing loss of source granularity, inability to distinguish internal versus external emergency transfers, and cohort discordance.

**Supplementary Figure S4**

**Reconstruction of Continuous Vasopressor Infusion Episodes from Standard OMOP Concepts**

**Figure legend**
**Supplementary Figure S4. Reconstruction of continuous vasopressor infusion from standardized OMOP concepts.** Multiple standardized ingredient, route, and frequency concepts representing vasopressor infusion were combined using temporal logic to reconstruct clinically meaningful infusion episodes. These were subsequently aggregated to generate the analytic outcome used for continuous vasopressor infusion duration, however our model variable was binary (yes/no).

Detailed derivation rules are provided in Supplementary Appendix A1.

**ALT TEXT**: Flowchart showing standardized OMOP vasopressor concepts combined using classification and temporal logic to reconstruct continuous infusion episodes, calculate total infusion duration, and derive a binary continuous vasopressor variable.

**Supplementary Figure S5**

**Reconstruction of Mechanical Ventilation Episodes from Standard OMOP Concepts**

**Figure legend**
**Supplementary Figure S5. Reconstruction of postoperative mechanical ventilation from standardized OMOP concepts.** Multiple standardized device and procedure concepts representing invasive ventilation were combined using temporal logic to reconstruct clinically meaningful ventilation episodes. Episode durations were subsequently aggregated to generate the analytic outcome used for postoperative respiratory failure classification.

The standardized concepts used to reconstruct invasive mechanical ventilation episodes are listed in Supplementary Appendix C. Detailed derivation rules are provided in Supplementary Appendix A2.

**ALT TEXT**: Flowchart showing standardized OMOP ventilation concepts classified as invasive mechanical ventilation, combined using temporal logic into episodes, aggregated into total ventilation duration, and classified as postoperative respiratory failure at ≥48 hours.
